# Validity and reliability of the SSF Preschoolers’ Activity Questionnaire (SSF-PAQ) for assessing active play, screen time, and sleep in Japanese preschool children

**DOI:** 10.64898/2026.11.17.26368286

**Authors:** Keisuke Komura, Mayumi Nagano, Rie Takenaga, Akira Kyan, Shigeru Inoue, Mitsuya Yamakita

**Affiliations:** Department of Liberal Arts, Faculty of Agriculture, Meijo University, Aichi, Japan; Graduate School of Environmental and Human Sciences, Meijo University, Aichi, Japan; International College of Arts and Sciences, Fukuoka Women’s University, Fukuoka, Japan; Sasakawa Sports Foundation, Tokyo, Japan; Faculty of Medicine, University of the Ryukyus, Okinawa, Japan; Center for Social Common Capital Beyond 2050, Office of Institutional Advancement and Communications, Kyoto University, Kyoto, Japan; Department of Preventive Medicine and Public Health, Tokyo Medical University, Tokyo, Japan; Faculty of Nursing, Yamanashi Prefectural University, Yamanashi, Japan

**Keywords:** accelerometry, sedentary behaviour, exercise, reproducibility of results, Japan

## Abstract

**Background:** The parent-reported Sasakawa Sports Foundation Preschoolers’ Activity Questionnaire (SSF-PAQ) is from a national survey of Japanese preschoolers’ out-of-preschool exercise, active play, screen time, and sleep. We assessed the validity, reliability, measurement error, paper–electronic agreement, and practicality of the Original version and explored a 24-hour version (SSF-PAQ-24).

**Methods:** Children aged 3–6 years from 10 conveniently sampled preschools wore an accelerometer for seven days while parents kept a time-use diary and completed the SSF-PAQ on Days 7 and 10. Criteria were accelerometer-measured physical activity (PA), diary-recorded screen time, and diary- and accelerometer-derived night sleep.

**Results:** The concurrent-validity sample comprised 167 children (Original, 127; SSF-PAQ-24, 40). Original-version correlations were weak for organized PA and active play (ρ = 0.24) and moderate-to-vigorous PA (ρ = 0.29) but strong for screen time (ρ = 0.83) and night sleep (ρ = 0.78), which averaged 8.2 and 10.7 min/day above the criteria. Test–retest intraclass correlations were 0.78–0.96 for these measures and 0.27–0.89 for setting-specific items. Screen time was higher on paper than electronically. SSF-PAQ-24 activity correlations were not significant (ρ = 0.17 and 0.06). Completion took 6.5–9.7 minutes.

**Conclusion:** The Original SSF-PAQ’s main measures showed good-to-excellent test–retest reliability, and screen time and night sleep correlated strongly with the criteria. Activity-time validity was limited but comparable to that of existing parent-report questionnaires. SSF-PAQ-24 activity items need re-examination in a larger sample.

## INTRODUCTION

Physical activity (PA), screen time, and sleep in early childhood are critical to health and development, as outlined in the World Health Organization’s 24-hour movement guidelines for children under 5 years.^1–4^ A pooled analysis across 33 countries found that only 14.3% of children aged 3–4 years met all three recommendations,^5^ highlighting the need for continuous monitoring.^6^ However, daily durations and guideline adherence do not indicate where PA could be increased. To design effective family and community interventions, the settings in which children exercise and play must be identified.

Such assessments require scalable, accurate measurement tools.^7^ While accelerometers objectively measure body movement, their high cost^8^ and complex data processing^9^ limit their use in large-scale surveys.

Conversely, parent proxy-report questionnaires are easy to administer and can capture the types and settings of exercise and play. However, a systematic review of 37 proxy-report questionnaires for children aged 0–5 years concluded that none was sufficiently valid and/or reliable and that the quality of evidence was mostly low or very low; only six questionnaires covered all three behaviours, and measurement error had been evaluated for only one.^7^ Since then, the Movement Behaviour Questionnaire–Child (MBQ-C), which assesses active play, screen time, and sleep, was developed in Australia and evaluated in Australia and China.^10–12^

In Japan, the Sasakawa Sports Foundation (SSF) surveyed 3,144 parents of preschool children nationwide in 2023.^13^ The survey’s activity, screen-time, and sleep items are collectively referred to in this study as the SSF Preschoolers’ Activity Questionnaire (SSF-PAQ). Unlike the Japanese version of the MBQ-C,^14^ which only provides a 24-hour overview, the SSF-PAQ asks about out-of-preschool organized sports or exercise lessons and outdoor and indoor active play by setting and day of the week, screen time, and weekday and weekend sleep schedules.^13^ However, the measurement properties of the SSF-PAQ have not been evaluated; interpreting the survey data and designing future surveys require clarifying its correspondence with accelerometers and diaries, setting-specific reliability, measurement error, and response burden. Because the survey was conducted online^13^ and validation in the intended mode of administration is recommended,^8^ paper–electronic agreement should also be examined. Testing a 24-hour version (SSF-PAQ-24), which extends the activity and screen-time items of the Original version to preschool hours, also helps identify challenges in whole-day assessment.

This study aimed to evaluate the concurrent validity, test–retest reliability, measurement error, agreement between paper and electronic administration modes, and practicality of the SSF-PAQ Original version in Japanese preschoolers. A secondary, exploratory aim was to evaluate the concurrent validity and test–retest reliability of the SSF-PAQ-24.

## METHODS

### Study design

This cross-sectional validation study shared a sample and data collection period with a separate validation study of the MBQ-C.^14^ Data were collected in May–June and October–December 2025. Reporting followed the STROBE statement (eTable 1).^15^ The Meijo University Human Research Ethics Committee approved the study (no. 2024-43), and each child’s parents/guardians provided written informed consent.

### Participants and group allocation

Children aged 3–6 years were recruited through 10 conveniently sampled preschools in four Japanese cities; no exclusion criteria were applied at recruitment. The sample size was set to detect ρ = 0.25 between the Original-version activity items (exercise and active play) and accelerometer-measured PA, based on earlier correlations of 0.20–0.39 between parent-reported play and accelerometry;^11,16^ this required 123 children with valid accelerometer data (α = 0.05, 80% power). Allowing for non-consent and missing data^17^ (eMethods 1), study documents were distributed to the parents of 623 children, and written informed consent was obtained for 208 children (33%).

Consenting children were allocated to six groups in a fixed, non-random rotation through a roster ordered by preschool and grade, to balance groups across both (eTable 2, eMethods 1). Once sufficient data for the Original version had been collected, subsequent participants were allocated to the 24-hour version. Parents completed their allocated version on Days 7 and 10, either in the same mode (paper or electronic) both times to assess test–retest reliability or once in each mode to assess paper–electronic agreement (Original version).

### Study protocol

Children wore the accelerometer on Days 1–7 (including during sleep), while parents maintained a 7-day time-use diary and completed the SSF-PAQ on Days 7 and 10 (eMethods 1).

### The SSF-PAQ

In the Original version,^13^ parents recalled a usual week over the past month, reporting daily out-of-preschool time across three domains: organized sports/exercise, outdoor active play (by location), and indoor active play (by setting). For each domain, they also noted time spent in activities intense enough to make the child breathe harder. Additionally, parents reported screen time (excluding body movement-based screen use), and usual weekday and weekend bedtime/wake-up time. From these responses, four daily-duration measures (min/day) were derived: organized physical activity and active play (hereafter ‘organized PA and AP’; organized lessons plus outdoor and indoor active play), moderate-to-vigorous PA (MVPA; total ‘breathing harder’ time), total screen time, and night sleep (from bedtime to wake-up time).

The SSF-PAQ-24 adds at-preschool rows to the play items and removes the ‘outside of preschool’ restriction from play and screen-time items, tracking these behaviours across the entire day. Organized-lessons, bedtime, and wake-up time items remain unchanged. Both versions were administered via paper and electronic modes (eMaterials 1–8).

### Criterion measures

PA was measured using a right hip-worn triaxial accelerometer (ActiGraph wGT3X-BT; ActiGraph LLC, Pensacola, FL, USA). Raw acceleration signals were classified into 15-second epochs using a machine-learning classifier for preschoolers^18,19^ (Preschool Hip Random Forest free-living classifier^18^ within the R package actimetric^20^). This yielded accelerometer-derived total PA and MVPA after excluding non-wear time.^21^ Mean Euclidean Norm Minus One (ENMO)^22^ provided a classifier-independent summary. To match each questionnaire’s timeframe, Original-version criteria captured activity outside of preschool hours, while the SSF-PAQ-24 criteria captured whole-day amounts.

Parents recorded screen time and sleep in a 24-hour time-use diary^11,23^ over all seven days. The screen-time criterion was defined as diary-recorded sedentary (sitting or lying) screen time; screen use while standing was excluded (eMethods 1). The night-sleep criterion integrated diary-reported bedtimes and wake-up times with bed rest estimated by an algorithm developed for hip-worn data in young children.^24^ Nights were classified as weekday or weekend based on whether the following day was a day off. Detailed processing methods for the accelerometer, diary, and sleep data are available in eMethods 1 and the companion paper.^14^

### Practicality

Practicality was assessed from the first Day 7 response: completion time, perceived response burden, and perceived clarity (eMethods 1).

### Data cleaning and processing

A valid accelerometer day required ≥ 600 minutes of wear^25^ between 07:00 and 21:00 hours.^13^ The primary analysis required at least three valid weekdays and one weekend day or holiday.^26,27^ Children with implausibly high MVPA relative to step counts were excluded (eMethods 1). Daily activity values were truncated at 480 minutes for organized PA and AP, and 360 minutes for MVPA, following the Australian MBQ-C validation study.^11^ The seven categories for SSF-PAQ screen time were converted to category midpoints, and night-sleep duration was calculated from reported bedtime and wake-up time. Weekday and weekend/holiday means were weighted as: (weekday × 5 + weekend/holiday × 2)/7.

### Statistical analysis

Concurrent validity (Day 7 SSF-PAQ versus criterion) was assessed using Spearman rank correlation coefficients (ρ) with 95% confidence intervals (CIs) derived via Fisher z transformation.^28^ Separate analyses were conducted for weekday, weekend/holiday, and weekly-average values. Following Schober et al.,^29^ coefficients were interpreted as weak (0.10–0.39), moderate (0.40–0.69), strong (0.70–0.89), or very strong (≥ 0.90). Activity items were also correlated with mean ENMO. Systematic differences in screen time and night sleep were evaluated using the Wilcoxon signed-rank test and Bland–Altman analysis.

Test–retest reliability (Day 7 versus Day 10; same version and mode) was assessed using intraclass correlation coefficients (ICCs) based on a two-way mixed-effects, single-measurement, absolute-agreement model.^30^ ICCs were interpreted following Koo and Li^31^ as moderate (0.50–0.75), good (0.75–0.90), or excellent (> 0.90). Measurement error was quantified using the within-child coefficient of variation (CV), the standard error of measurement (SEM) from a mixed model with random effects for participant and administration, and the smallest detectable change (SDC) at both the individual (SDCind)^32^ and group levels (SDCgroup).^33^ Paper–electronic agreement (Original version) was assessed with Spearman ρ, ICCs, and Wilcoxon signed-rank tests, while practicality metrics were summarized descriptively.

Validity and reliability were estimated by mode, while reliability and measurement error were calculated for setting-specific activity items. For the SSF-PAQ-24, at-preschool active play and out-of-preschool organized PA and AP were validated against corresponding period accelerometer-derived total PA. The same properties were evaluated for bedtime, wake-up time, midsleep (their midpoint), and social jetlag (the signed weekend−weekday difference in midsleep^34^), against accelerometer-estimated sleep timing. Because the national survey also reports days per week,^13^ Original-version days per week with any organized PA or AP (activity days) and with outdoor active play (outdoor-play days) were validated against accelerometer-derived out-of-preschool total PA (Spearman ρ) and assessed for test–retest reliability (ICC; eMethods 1).

Except for missing diary clock times in the night-sleep criterion (eMethods 1), no missing data were imputed; analyses included all participants with complete data for the required variables (n in each table). Sensitivity analyses re-examined concurrent validity under stricter and more lenient^35^ wear-time criteria, among children aged 3–5 years, and with criteria recalculated from days reported as usual, accurately recorded, or both; for night sleep, criterion weekend nights were also reclassified by calendar weekend (eMethods 1). All analyses were performed using R version 4.5.0. Claude (Anthropic), a generative artificial intelligence tool, assisted with writing and checking the analysis code, which was verified by the corresponding author.

## RESULTS

### Participants

Of the 208 children whose parents consented, 171 provided valid accelerometer data, of whom 167 completed the Day 7 SSF-PAQ and formed the concurrent-validity sample (Original: *n* = 127; 24-hour: *n* = 40; Figure 1). Their mean age was 4.95 years and 52% were girls; children in the 24-hour version were older, included more girls, and more often attended nursery schools than those in the Original version (Table 1).

**Figure 1.**
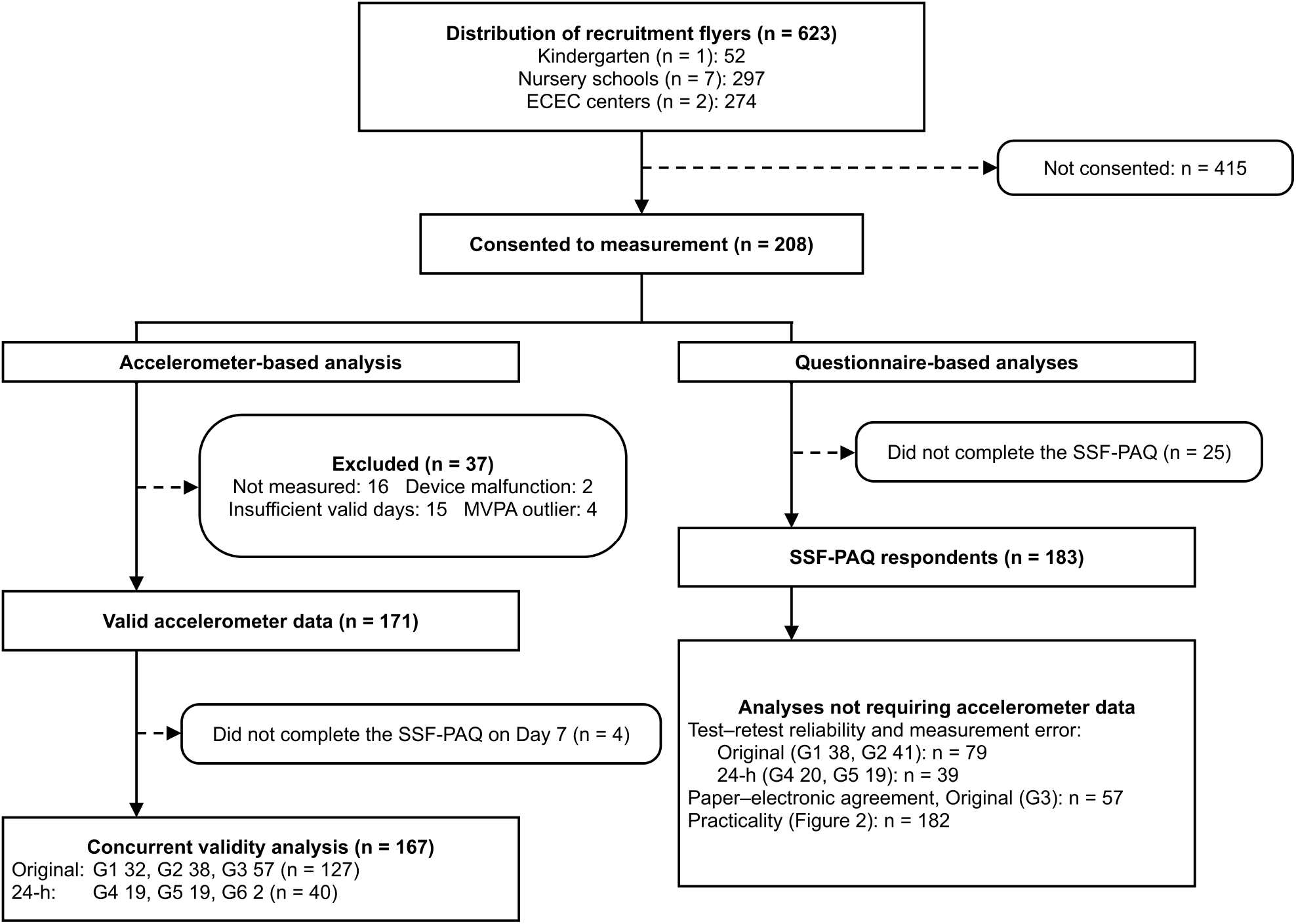
Participant flow and derivation of the analytic samples. The two streams overlap: the 167 children in the concurrent validity analysis were a subset of the 183 SSF-PAQ respondents. As the questionnaire-based analyses did not require accelerometer data, sample sizes differed between the questionnaire and concurrent validity analyses and should not be summed. Likewise, the two G3 samples of 57 participants overlapped but were not identical (54 children in common). Groups 1 and 4 completed the questionnaire on paper at both administrations, Groups 2 and 5 electronically at both, and Groups 3 and 6 switched modes (eTable 2). ECEC, early childhood education and care; G, group; MVPA, moderate-to-vigorous physical activity; SSF-PAQ, Sasakawa Sports Foundation Preschoolers’ Activity Questionnaire.

**Table 1.** Participant characteristics by SSF-PAQ version.

| **Characteristics** | **SSF-PAQ Original version (*N* = 127)** | **SSF-PAQ 24-h version (*N* = 40)** | **Overall (*N* = 167)** |
|---|---|---|---|
| **Children’s age (years)** | 4.76 (0.79) | 5.55 (0.72) | 4.95 (0.84) |
| **Age category** |  |  |  |
| 3 years | 27 (21%) | 2 (5.0%) | 29 (17%) |
| 4 years | 50 (39%) | 8 (20%) | 58 (35%) |
| 5 years | 45 (35%) | 17 (43%) | 62 (37%) |
| 6 years | 5 (3.9%) | 13 (33%) | 18 (11%) |
| **Gender** |  |  |  |
| Boy | 65 (51%) | 15 (38%) | 80 (48%) |
| Girl | 62 (49%) | 25 (63%) | 87 (52%) |
| **Type of preschool** |  |  |  |
| Kindergarten | 5 (3.9%) | 0 (0%) | 5 (3.0%) |
| Nursery school | 32 (25%) | 28 (70%) | 60 (36%) |
| ECEC center | 90 (71%) | 12 (30%) | 102 (61%) |
| **Time at preschool (min/day)** | 426 (83) | 479 (84) | 438 (86) |
| Missing | 1 | 2 | 3 |
| **Respondents** |  |  |  |
| Mother | 113 (89%) | 36 (92%) | 149 (90%) |
| Father | 12 (9.4%) | 3 (7.7%) | 15 (9.0%) |
| Grandparent | 2 (1.6%) | 0 (0%) | 2 (1.2%) |
| Missing | 0 | 1 | 1 |
| **Respondent age (years)** | 37.2 (6.1) | 37.9 (5.2) | 37.4 (5.9) |
| Missing | 0 | 1 | 1 |
| **Parent respondent education** |  |  |  |
| Junior high or high school | 25 (21%) | 5 (13%) | 30 (19%) |
| Technical/junior college or vocational school | 36 (30%) | 13 (34%) | 49 (31%) |
| University or graduate school | 58 (49%) | 20 (53%) | 78 (50%) |
| Missing | 8 | 2 | 10 |
| **Household income (million JPY)** |  |  |  |
| < 3.0 | 4 (4.6%) | 2 (6.9%) | 6 (5.2%) |
| 3.0 to < 5.0 | 15 (17%) | 0 (0%) | 15 (13%) |
| 5.0 to < 8.0 | 32 (37%) | 12 (41%) | 44 (38%) |
| 8.0 to < 10.0 | 22 (25%) | 5 (17%) | 27 (23%) |
| ≥ 10.0 | 14 (16%) | 10 (34%) | 24 (21%) |
| Missing | 40 | 11 | 51 |
| **Accelerometer and diary measures** |  |  |  |
| Total physical activity, accelerometer (min/day) | 402 (53) | 384 (70) | 398 (58) |
| MVPA, accelerometer (min/day) | 82 (39) | 80 (25) | 81 (36) |
| Total screen time, diary (min/day) | 100 (70) | 159 (82) | 115 (77) |
| Total sleep, accelerometer + diary (min/day) | 607 (36) | 609 (45) | 607 (39) |
| Diary days rated as usual (%) | 88 (14) | 68 (26) | 83 (19) |
| Diary days rated as accurately recorded (%) | 91 (15) | 83 (24) | 89 (18) |

ECEC, early childhood education and care; JPY, Japanese yen; MVPA, moderate-to-vigorous physical activity; SD, standard deviation; SSF-PAQ, Sasakawa Sports Foundation Preschoolers' Activity Questionnaire.
Participants with valid accelerometer data who completed the SSF-PAQ on Day 7 (the concurrent-validity sample). Continuous variables are presented as mean (SD) and categorical variables as *n* (%). Total physical activity, MVPA, total screen time, and total sleep represent whole-day values expressed as weighted weekly averages [(weekday × 5 + weekend/holiday × 2)/7]. Total screen time includes standing screen use, and total sleep includes daytime sleep; therefore, these measures differ from the criterion measures in Table 2. Diary days rated as usual and accurately recorded are expressed as the percentage of each child’s diary days so rated by the parent.

### Concurrent validity

Concurrent validity varied by behaviour (Table 2). For the Original version (weekly average), correlations with the criterion were weak for the activity items (organized PA and AP, ρ = 0.24; MVPA, ρ = 0.29) but strong for screen time (ρ = 0.83) and night sleep (ρ = 0.78). In the 24-hour version, the activity correlations were not significant (ρ = 0.17 and 0.06), whereas screen time (ρ = 0.68) and night sleep (ρ = 0.63) correlated moderately. Against mean ENMO, results were comparable except that the Original-version MVPA coefficient rose from 0.29 to 0.44. In the Original version, SSF-PAQ values exceeded the criteria by a mean of 8.2 min/day for screen time and 10.7 min/day for night sleep (95% limits of agreement, −72.3 to 88.8 and −33.2 to 54.7 min/day), without significant proportional bias (eFigure 1). Night-sleep validity was lower on weekends than weekdays in both versions (0.50 versus 0.73; 0.41 versus 0.57; eTable 3, Part A). Weekday 24-hour reports, when divided into out-of-preschool and at-preschool time, yielded weak criterion correlations (0.19 and 0.24, respectively), with both 95% CIs including zero (eTable 3, Part B).

**Table 2.** Validity coefficients of the SSF-PAQ by version.

| **Criterion measure** | **SSF-PAQ item** | **Period** | ***n*** | **ρ [95% CI]** | **ρ versus mean ENMO [95% CI]** | **Criterion Med (Q1–Q3)** | **SSF-PAQ Med (Q1–Q3)** | **P** |
|---|---|---|---|---|---|---|---|---|
| **SSF-PAQ Original version** |  |  |  |  |  |  |  |  |
| Total PA (ACC, out-of-preschool) | Organized PA and AP | Weekday | 127 | **0.38 [0.22, 0.52]** | **0.26 [0.09, 0.41]** | 185 (142–226) | 35 (15–75) | NA |
|  |  | Weekend | 127 | 0.06 [−0.12, 0.23] | 0.14 [−0.04, 0.30] | 394 (332–433) | 150 (90–222) | NA |
|  |  | Weekly | 127 | **0.24 [0.07, 0.39]** | **0.22 [0.05, 0.38]** | 238 (200–282) | 73 (47–101) | NA |
| MVPA (ACC, out-of-preschool) | MVPA | Weekday | 127 | **0.34 [0.18, 0.49]** | **0.39 [0.23, 0.52]** | 34 (23–49) | 5 (0–16) | NA |
|  |  | Weekend | 127 | **0.31 [0.14, 0.46]** | **0.34 [0.18, 0.49]** | 70 (51–99) | 30 (10–50) | NA |
|  |  | Weekly | 127 | **0.29 [0.13, 0.45]** | **0.44 [0.28, 0.57]** | 46 (32–61) | 13 (4–26) | NA |
| Sedentary screen time (diary) | Total screen time | Weekday | 127 | **0.81 [0.74, 0.86]** | — | 72 (27–117) | 90 (45–150) | < 0.001 |
|  |  | Weekend | 126 | **0.78 [0.70, 0.84]** | — | 105 (66–179) | 150 (90–150) | 0.576 |
|  |  | Weekly | 126 | **0.83 [0.77, 0.88]** | — | 87 (43–135) | 91 (58–150) | 0.001 |
| Night sleep (diary + ACC) | Night sleep | Weekday | 127 | **0.73 [0.63, 0.80]** | — | 585 (562–612) | 600 (570–630) | < 0.001 |
|  |  | Weekend | 127 | **0.50 [0.35, 0.62]** | — | 602 (569–637) | 600 (585–630) | 0.170 |
|  |  | Weekly | 127 | **0.78 [0.71, 0.84]** | — | 591 (570–612) | 600 (579–627) | < 0.001 |
| **SSF-PAQ 24-h version** |  |  |  |  |  |  |  |  |
| Total PA (ACC) | Organized PA and AP | Weekday | 40 | 0.20 [−0.12, 0.48] | 0.20 [−0.12, 0.48] | 393 (345–426) | 97 (40–153) | NA |
|  |  | Weekend | 40 | −0.03 [−0.34, 0.28] | 0.18 [−0.14, 0.47] | 391 (340–438) | 95 (60–161) | NA |
|  |  | Weekly | 40 | 0.17 [−0.15, 0.46] | 0.16 [−0.16, 0.45] | 389 (342–429) | 92 (48–160) | NA |
| MVPA (ACC) | MVPA | Weekday | 40 | −0.01 [−0.32, 0.30] | 0.16 [−0.16, 0.45] | 71 (46–92) | 16 (2–26) | NA |
|  |  | Weekend | 40 | 0.09 [−0.23, 0.39] | 0.10 [−0.22, 0.40] | 93 (84–112) | 25 (9–49) | NA |
|  |  | Weekly | 40 | 0.06 [−0.26, 0.36] | 0.14 [−0.18, 0.43] | 82 (63–100) | 21 (8–37) | NA |
| Sedentary screen time (diary) | Total screen time | Weekday | 40 | **0.64 [0.42, 0.80]** | — | 107 (64–213) | 90 (45–150) | 0.049 |
|  |  | Weekend | 40 | **0.53 [0.27, 0.72]** | — | 158 (109–221) | 150 (90–210) | 0.242 |
|  |  | Weekly | 40 | **0.68 [0.47, 0.82]** | — | 136 (78–208) | 107 (58–174) | 0.064 |
| Night sleep (diary + ACC) | Night sleep | Weekday | 40 | **0.57 [0.31, 0.75]** | — | 591 (568–607) | 595 (570–600) | 0.764 |
|  |  | Weekend | 40 | **0.41 [0.12, 0.64]** | — | 599 (575–636) | 600 (570–630) | 0.879 |
|  |  | Weekly | 40 | **0.63 [0.40, 0.79]** | — | 594 (574–610) | 590 (570–609) | 0.806 |

ACC, accelerometer-derived; AP, active play; CI, confidence interval; diary, time-use diary; ENMO, Euclidean Norm Minus One; Med, median; MVPA, moderate-to-vigorous physical activity; organized PA, organized physical activity; PA, physical activity; Q1–Q3, interquartile range; SSF-PAQ, Sasakawa Sports Foundation Preschoolers' Activity Questionnaire.
Concurrent validity at Day 7 among participants with valid accelerometer data. Paper and electronic modes were pooled (mode-specific estimates are presented in eTable 3, Part A); values are expressed as min/day. ρ, Spearman rank correlation coefficient (bold where the 95% CI excludes 0). The ρ versus mean ENMO column shows correlations between physical activity items and mean acceleration over the same period (—, not applicable to screen time or sleep). P, Wilcoxon signed-rank test for systematic differences between paired criterion and SSF-PAQ values; NA, not examined because the accelerometer-derived criteria are not equivalent to the questionnaire estimates.

### Test–retest reliability

Weekly-average behaviours showed good-to-excellent test–retest reliability across both versions (Table 3): ICCs were 0.78–0.96 (Original) and 0.82–0.91 (24-hour), peaking for night sleep in both. Reliability by mode and setting is shown in eTable 3 (Part C) and eTable 4, respectively. In the Original version, weekly-average ICCs were good for six of the nine settings (0.76–0.89) but fell below 0.75 for outdoor active play in parks (0.66), indoor active play at facilities (0.57), and other outdoor active play (0.27). For outdoor active play in parks and on home premises, weekend ICCs (0.55 and 0.34) were lower than weekday ICCs (0.78 and 0.89).

**Table 3.** Test–retest reliability of the SSF-PAQ by version.

| SSF-PAQ item | Period | *n* | ICC [95% CI] | Day 7 Med (Q1–Q3) | Day 10 Med (Q1–Q3) |
|---|---|---|---|---|---|
| **SSF-PAQ Original version** |  |  |  |  |  |
| Organized PA and AP | Weekday | 79 | **0.84 [0.76, 0.89]** | 57 (20–90) | 51 (21–77) |
|  | Weekend | 79 | **0.82 [0.73, 0.88]** | 155 (109–265) | 120 (92–232) |
|  | Weekly | 79 | **0.84 [0.75, 0.89]** | 86 (54–121) | 75 (50–105) |
| MVPA | Weekday | 79 | **0.91 [0.87, 0.94]** | 8 (0–20) | 9 (0–20) |
|  | Weekend | 79 | **0.80 [0.71, 0.87]** | 38 (10–60) | 38 (20–60) |
|  | Weekly | 79 | **0.85 [0.77, 0.90]** | 17 (4–34) | 19 (12–34) |
| Total screen time | Weekday | 78 | **0.76 [0.65, 0.84]** | 90 (45–150) | 90 (45–150) |
|  | Weekend | 78 | **0.79 [0.69, 0.86]** | 120 (90–150) | 120 (90–150) |
|  | Weekly | 77 | **0.78 [0.68, 0.86]** | 107 (58–150) | 107 (45–150) |
| Night sleep | Weekday | 79 | **0.97 [0.95, 0.98]** | 600 (570–630) | 600 (570–630) |
|  | Weekend | 79 | **0.83 [0.75, 0.89]** | 600 (585–630) | 600 (597–630) |
|  | Weekly | 79 | **0.96 [0.94, 0.97]** | 604 (579–621) | 600 (579–621) |
| **SSF-PAQ 24-h version** |  |  |  |  |  |
| Organized PA and AP | Weekday | 39 | **0.79 [0.64, 0.89]** | 96 (40–140) | 90 (54–120) |
|  | Weekend | 39 | **0.72 [0.52, 0.84]** | 95 (60–162) | 80 (56–135) |
|  | Weekly | 39 | **0.86 [0.74, 0.92]** | 91 (47–150) | 87 (60–116) |
| MVPA | Weekday | 39 | **0.66 [0.44, 0.80]** | 12 (1–25) | 24 (4–39) |
|  | Weekend | 39 | **0.75 [0.57, 0.86]** | 25 (8–52) | 25 (14–45) |
|  | Weekly | 39 | **0.82 [0.68, 0.90]** | 18 (8–33) | 24 (9–37) |
| Total screen time | Weekday | 39 | **0.80 [0.66, 0.89]** | 90 (45–150) | 90 (45–150) |
|  | Weekend | 39 | **0.84 [0.72, 0.91]** | 150 (90–210) | 150 (90–210) |
|  | Weekly | 39 | **0.83 [0.70, 0.91]** | 107 (58–167) | 107 (58–159) |
| Night sleep | Weekday | 39 | **0.95 [0.91, 0.97]** | 600 (570–600) | 590 (570–600) |
|  | Weekend | 39 | **0.60 [0.35, 0.77]** | 600 (570–630) | 600 (600–630) |
|  | Weekly | 39 | **0.91 [0.84, 0.95]** | 591 (570–609) | 599 (575–609) |

AP, active play; CI, confidence interval; Med, median; MVPA, moderate-to-vigorous physical activity; organized PA, organized physical activity; PA, physical activity; Q1–Q3, interquartile range; SSF-PAQ, Sasakawa Sports Foundation Preschoolers' Activity Questionnaire.
Test–retest reliability among participants who completed the same SSF-PAQ version in the same mode on Days 7 and 10 (paper at both administrations or electronic at both); paper and electronic groups were pooled. Mode-specific estimates are presented in eTable 3 (Part C). Values are expressed as min/day. ICC, intraclass correlation coefficient (bold where ≥ 0.50).

### Measurement error

Measurement error differed by behaviour (Table 4). In both versions, the weekly-average within-child CV was highest for MVPA and lowest for night sleep, and SDCind was highest for screen time and lowest for night sleep. Setting-specific values are in eTable 5.

**Table 4.** Measurement error in repeated assessments of the SSF-PAQ.

| **SSF-PAQ item** | **Period** | ***n*** | **Day 7**<br>**Mean (SD)** | **Day 10**<br>**Mean (SD)** | **Difference**<br>**Mean (SD)** | **CV**<br>**(%)** | **SEM** | **SDCind** | **SDCgroup**<br>**(SDCind/√*N*)** |
|---|---|---|---|---|---|---|---|---|---|
| **SSF-PAQ Original version** |  |  |  |  |  |  |  |  |  |
| Organized PA and AP | Weekday | 79 | 63 (54) | 57 (50) | 5 (29) | 30.6 | 20.9 | 57.9 | 57.9/√*N* |
|  | Weekend | 79 | 182 (107) | 166 (99) | 17 (60) | 21.3 | 43.8 | 121.5 | 121.5/√*N* |
|  | Weekly | 79 | 98 (62) | 90 (62) | 8 (35) | 18.4 | 25.1 | 69.6 | 69.6/√*N* |
| MVPA | Weekday | 79 | 16 (23) | 16 (24) | 0 (10) | 38.6 | 7.0 | 19.3 | 19.3/√*N* |
|  | Weekend | 79 | 47 (53) | 54 (60) | −6 (35) | 43.3 | 25.3 | 70.1 | 70.1/√*N* |
|  | Weekly | 79 | 25 (28) | 28 (35) | −3 (18) | 40.5 | 12.5 | 34.7 | 34.7/√*N* |
| Total screen time | Weekday | 78 | 94 (64) | 89 (59) | 5 (42) | 16.7 | 30.0 | 83.2 | 83.2/√*N* |
|  | Weekend | 78 | 130 (73) | 124 (75) | 6 (48) | 15.7 | 34.0 | 94.2 | 94.2/√*N* |
|  | Weekly | 77 | 105 (65) | 100 (61) | 5 (42) | 16.6 | 29.5 | 81.6 | 81.6/√*N* |
| Night sleep | Weekday | 79 | 600 (44) | 601 (44) | −1 (12) | 0.7 | 8.2 | 22.8 | 22.8/√*N* |
|  | Weekend | 79 | 609 (36) | 609 (38) | 0 (22) | 1.6 | 15.4 | 42.7 | 42.7/√*N* |
|  | Weekly | 79 | 603 (38) | 603 (39) | 0 (11) | 0.8 | 7.7 | 21.3 | 21.3/√*N* |
| **SSF-PAQ 24-h version** |  |  |  |  |  |  |  |  |  |
| Organized PA and AP | Weekday | 39 | 101 (72) | 92 (59) | 9 (42) | 22.0 | 29.9 | 82.9 | 82.9/√*N* |
|  | Weekend | 39 | 119 (95) | 107 (82) | 11 (67) | 29.0 | 47.3 | 131.0 | 131.0/√*N* |
|  | Weekly | 39 | 106 (71) | 96 (59) | 10 (34) | 21.3 | 24.7 | 68.6 | 68.6/√*N* |
| MVPA | Weekday | 39 | 20 (26) | 26 (24) | −6 (20) | 37.1 | 14.8 | 41.0 | 41.0/√*N* |
|  | Weekend | 39 | 40 (46) | 36 (38) | 4 (30) | 44.7 | 21.0 | 58.3 | 58.3/√*N* |
|  | Weekly | 39 | 26 (28) | 29 (25) | −3 (16) | 37.0 | 11.4 | 31.7 | 31.7/√*N* |
| Total screen time | Weekday | 39 | 108 (78) | 104 (71) | 4 (47) | 24.5 | 33.0 | 91.5 | 91.5/√*N* |
|  | Weekend | 39 | 152 (87) | 156 (81) | −4 (47) | 19.6 | 33.2 | 92.0 | 92.0/√*N* |
|  | Weekly | 39 | 120 (78) | 119 (71) | 2 (43) | 21.2 | 30.4 | 84.2 | 84.2/√*N* |
| Night sleep | Weekday | 39 | 584 (31) | 583 (31) | 1 (10) | 0.5 | 7.0 | 19.5 | 19.5/√*N* |
|  | Weekend | 39 | 602 (40) | 610 (34) | −9 (33) | 2.1 | 23.9 | 66.2 | 66.2/√*N* |
|  | Weekly | 39 | 589 (30) | 591 (28) | −2 (12) | 0.9 | 8.7 | 24.1 | 24.1/√*N* |

AP, active play; CV, mean within-child coefficient of variation; MVPA, moderate-to-vigorous physical activity; organized PA, organized physical activity; PA, physical activity; SD, standard deviation; SDC, smallest detectable change; SEM, standard error of measurement; SSF-PAQ, Sasakawa Sports Foundation Preschoolers' Activity Questionnaire.
Repeated SSF-PAQ assessments were conducted on Days 7 and 10 using the same version and mode (paper at both administrations or electronic at both); paper and electronic groups were pooled. Values are min/day. The difference is Day 7 minus Day 10. SDCind = 1.96 × √2 × SEM is the individual-level smallest detectable change (minimal detectable change at the 95% level, MDC95). SDCgroup = SDCind/√*N* is the smallest detectable change in the mean of *N* participants; the systematic between-occasion difference is unaffected by increasing *N*.

### Agreement between paper and electronic modes

Rank agreement between the paper and electronic modes of the Original version was strong across all behaviours (weekly-average ρ = 0.75–0.87; Table 5). Absolute agreement was moderate for organized PA and AP, MVPA, and screen time (ICC = 0.55–0.70) and excellent for night sleep (ICC = 0.91). Screen time showed a systematic difference, yielding higher values on paper (P = 0.002); the mean difference was similar in both administration orders (Table 5).

**Table 5.** Agreement of the SSF-PAQ Original version between paper and electronic modes.

| SSF-PAQ item | Period | *n* | ρ [95% CI] | ICC [95% CI] | Med 1 (Q1–Q3) | Med 2 (Q1–Q3) | P |
|---|---|---|---|---|---|---|---|
| Organized PA and AP | Weekday | 57 | **0.76 [0.62, 0.85]** | **0.77 [0.63, 0.86]** | 30 (15–50) | 39 (13–60) | 0.009 |
|  | Weekend | 57 | **0.76 [0.63, 0.85]** | **0.67 [0.50, 0.79]** | 140 (78–215) | 130 (75–240) | 0.217 |
|  | Weekly | 57 | **0.77 [0.64, 0.86]** | **0.55 [0.34, 0.71]** | 60 (41–91) | 61 (45–96) | 0.045 |
| MVPA | Weekday | 57 | **0.83 [0.73, 0.90]** | 0.39 [0.15, 0.58] | 5 (0–10) | 5 (0–12) | 0.085 |
|  | Weekend | 57 | **0.68 [0.51, 0.80]** | **0.59 [0.39, 0.74]** | 30 (10–45) | 25 (10–60) | 0.879 |
|  | Weekly | 57 | **0.75 [0.61, 0.85]** | **0.56 [0.35, 0.71]** | 11 (6–21) | 11 (4–26) | 0.518 |
| Total screen time | Weekday | 56 | **0.80 [0.68, 0.88]** | **0.67 [0.46, 0.80]** | 68 (45–150) | 45 (15–90) | 0.004 |
|  | Weekend | 56 | **0.75 [0.61, 0.85]** | **0.74 [0.58, 0.84]** | 120 (90–150) | 90 (45–150) | 0.013 |
|  | Weekly | 56 | **0.78 [0.65, 0.87]** | **0.70 [0.50, 0.82]** | 84 (58–150) | 58 (36–111) | 0.002 |
| Night sleep | Weekday | 57 | **0.91 [0.85, 0.94]** | **0.92 [0.87, 0.95]** | 600 (570–630) | 600 (570–620) | 0.674 |
|  | Weekend | 57 | **0.79 [0.67, 0.87]** | **0.84 [0.74, 0.90]** | 600 (585–630) | 600 (600–630) | 0.512 |
|  | Weekly | 57 | **0.87 [0.79, 0.92]** | **0.91 [0.86, 0.95]** | 600 (579–629) | 600 (587–619) | 0.600 |

AP, active play; CI, confidence interval; ICC, intraclass correlation coefficient; Med, median; MVPA, moderate-to-vigorous physical activity; organized PA, organized physical activity; PA, physical activity; Q1–Q3, interquartile range; ρ, Spearman rank correlation coefficient; SSF-PAQ, Sasakawa Sports Foundation Preschoolers' Activity Questionnaire.
Agreement between the paper and electronic modes of the SSF-PAQ Original version, assessed in the mode-switch subgroup in which each child completed one mode on Day 7 and the other on Day 10. Administration orders were pooled, and each child’s paper and electronic responses were compared. Of the 57 children, 41 completed the paper mode on Day 7 and the electronic mode on Day 10, and 16 completed them in the reverse order (41 and 15 for screen time); the mean paper-minus-electronic difference in weekly-average screen time was 20 and 18 min/day, respectively. Values are min/day. Med 1, paper median; Med 2, electronic median. ρ is shown in bold where the 95% CI excludes 0, and ICC where ≥ 0.50. P, Wilcoxon signed-rank test of within-child paired differences between modes (not a correlation *P* value).

### Practicality

Mean completion time was 6.5–9.7 minutes across groups and was shorter in the electronic mode for both versions (by 0.6 and 3.2 minutes; Figure 2). While 45–53% of parents rated the questionnaire items as hardly or not very burdensome, 46–59% rated them as very or somewhat clear.

**Figure 2.**
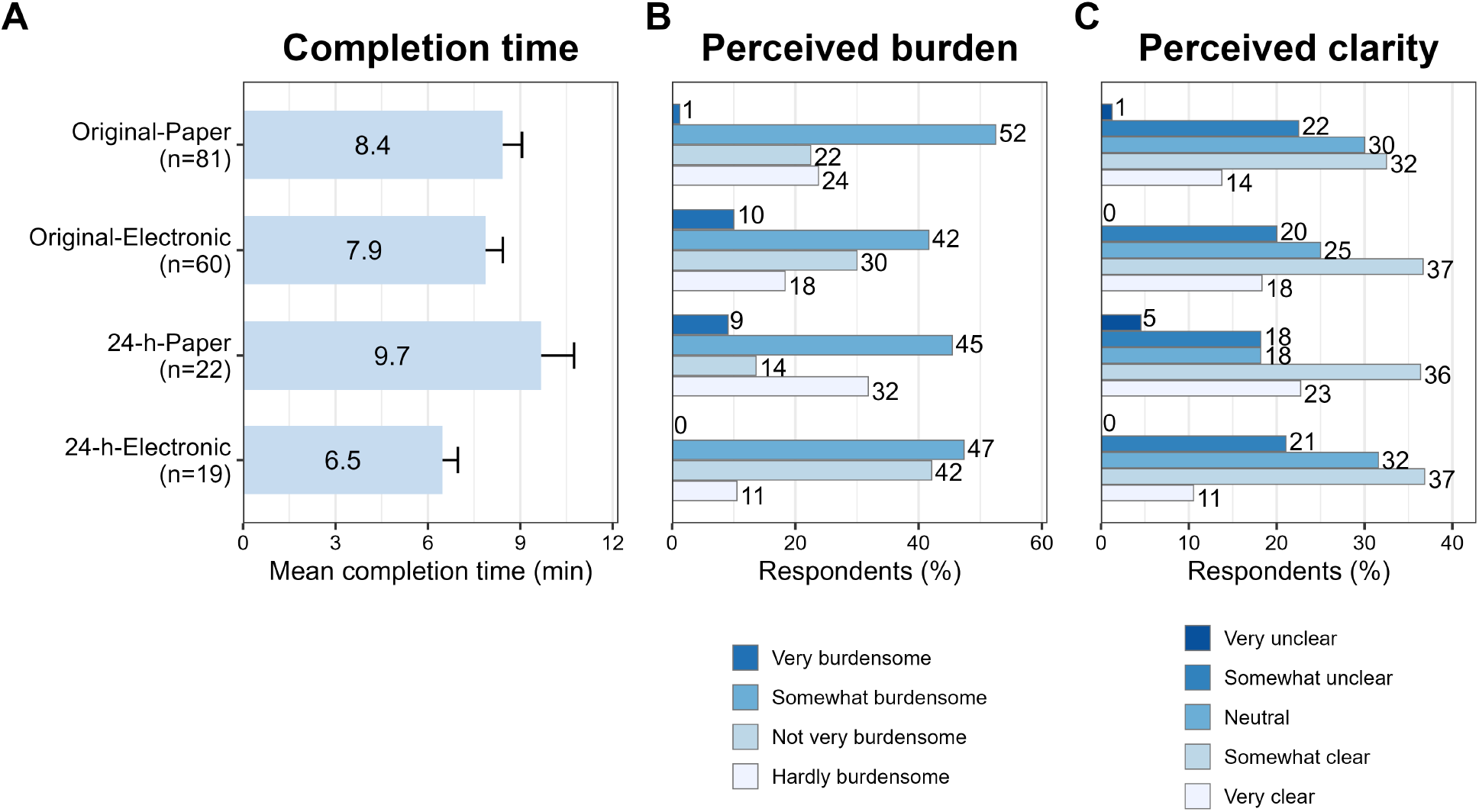
Practicality of the SSF-PAQ. (A) Mean completion time (min); error bars indicate the standard error of the mean, and group sample sizes are shown on the y-axis labels. (B) Perceived burden and (C) perceived clarity, presented as percentages of respondents in each category. In the Original-Paper group, perceived burden and clarity were each based on *n* = 80. SSF-PAQ, Sasakawa Sports Foundation Preschoolers’ Activity Questionnaire.

### Sensitivity analyses

Behaviour-specific patterns were unchanged under other wear-time criteria (360/960 min/day; eTable 6), at ages 3–5 years (eTable 7), and with criteria from usual days, accurately recorded days, or both (eTable 8).

For night sleep (pooled across identical versions), reclassifying weekend nights from ‘nights before a day off’ to ‘calendar weekend nights’ lowered the weekend correlation from 0.49 to 0.33, whereas weekday and weekly-average coefficients remained similar (eTable 9).

### Sleep timing

For sleep timing (pooled versions; eTable 10), SSF-PAQ bedtime, wake-up time, and midsleep correlated strongly to very strongly with the criterion (ρ = 0.78–0.91), showing small systematic differences (median within-child differences ≤ 16 minutes), excellent reliability (ICC = 0.92–0.99), and small measurement error (SEM = 3–15 minutes). Social jetlag correlated weakly with the criterion (ρ = 0.36) but showed good reliability (ICC = 0.83).

### Day-based indicators

Original-version activity days (seven per week in 60% of children) did not correlate significantly with accelerometer-derived out-of-preschool total PA (ρ = 0.12), whereas outdoor-play days correlated weakly (ρ = 0.32); test–retest ICCs were 0.65 and 0.90, respectively (eTable 11).

## DISCUSSION

This study is the first to evaluate the measurement properties of the SSF-PAQ, a questionnaire used in a national survey of preschoolers, alongside an exploratory 24-hour extension (SSF-PAQ-24). Whereas measurement error had been evaluated for only one of 37 proxy-report questionnaires for young children,^7^ this study quantified it by behaviour and setting.

Although the SSF-PAQ activity items were limited to out-of-preschool time, easily observable by parents, and asked for detailed daily and setting-specific information, their correspondence with accelerometer-measured activity time was weak. Correlations were similarly low for the Japanese MBQ-C in the same sample (ρ = 0.12–0.26),^14^ the original MBQ-C (ρ = 0.20–0.39),^11^ and the Preschool-age Children’s Physical Activity Questionnaire, which asks about home activity by type (r = −0.07 to 0.17).^36^ Beyond the conceptual gap between questionnaire-reported time (spent in exercise and play) and accelerometer-measured time^14^ (spent in actual movement), parents may have difficulty in quantifying young children’s intermittent activity into specific durations.^7^ Original-version ‘breathing harder’ time correlated more strongly with mean ENMO (0.44), a summary measure of activity volume, than with accelerometer-derived MVPA time (0.29). This suggests that parents may rely on their overall impression of how active the child is.

In the 24-hour version, correlations were also low and imprecisely estimated (ρ = 0.17 and 0.06). These children, 70% of whom attended nursery schools, spent a mean of 8 hours per day at preschool (Table 1), and parents have difficulty monitoring activity during childcare hours.^11^ However, among the same children, weekday correlations were weak both outside of preschool (ρ = 0.19) and at preschool (ρ = 0.24; eTable 3, Part B). Thus, limited knowledge of preschool hours is unlikely to be the only explanation, and limiting questions to easily observed periods and asking for greater details may not overcome parents’ difficulty in estimating activity duration.

Although setting-specific validity was not tested, the weekly averages of six Original-version items (including organized lessons and indoor active play) showed good reliability, indicating consistent parent reporting. However, weekend ICCs were lower for outdoor active play in parks (0.55) and at home (0.34), suggesting that weekend outdoor-play items warrant further evaluation. Furthermore, because only 16–23% of responses exceeded 0 minutes for screen-based and ‘other’ active play, ICCs alone cannot determine if these low-frequency behaviours are captured reliably (eTable 4).

To help interpret group changes against measurement error, this study reported the SEM and SDC in units of time. In longitudinal surveys, SDCgroup (SDCind/√N) can determine whether a mean change exceeds measurement error.^33^ For example, in a cohort of 100 children, the threshold values are approximately 7.0, 3.5, and 8.2 min/day for organized PA and AP, MVPA, and screen time, respectively. SDCgroup does not account for systematic response shifts or participant turnover, which must be considered separately. In contrast, individual changes smaller than the SDCind (Table 4) cannot be distinguished from measurement error.

Although Original-version screen time was measured in categories, its diary correlation was as high as that of sedentary screen time in the Japanese MBQ-C (0.83 versus 0.81 for the open-ended and 0.78 for the closed-ended formats),^14^ with good reliability (ICC = 0.78). However, questionnaire values were systematically higher than diary values (mean difference, 8.2 min/day), mirroring MBQ-C findings,^14^ with wide limits of agreement. Thus, group surveys can capture relative differences in screen time, but individual values need to be interpreted with caution.

The Original-version night-sleep correlation (ρ = 0.78) exceeded that of the Japanese MBQ-C in the same recruited sample (0.56).^14^ This discrepancy may relate to format differences: the MBQ-C asks directly about usual night-sleep duration, whereas the SSF-PAQ derives it from separate weekday and weekend bedtime/wake-up times. In Iwasaki et al.’s study,^37^ responses on usual sleep over the past 4 weeks (without distinguishing weekday/weekend) correlated with actigraphy-measured sleep on weekdays, but not significantly on weekends. Coupled with the strong-to-very-strong correlations found here for bedtime, wake-up time, and midsleep, these results suggest that a format separating day-of-week differences in daily schedules and using specific clock times as cues is useful. The drop in the weekend night-sleep correlation (0.49 to 0.33) when reclassifying criterion weekend nights as calendar Saturday and Sunday nights suggests that parents answered keeping nights before a day off in mind. Hence, explicitly asking about ‘nights before a day off’ may resolve this ambiguity and improve validity. Nevertheless, the 95% limits of agreement with the criterion were −33.2 to 54.7 min/day, indicating limited precision for individual estimates.

Paper and electronic formats ranked children similarly, but absolute agreement was only moderate for activity and screen-time items, with paper yielding higher mean screen times in both administration orders. Therefore, these two modes should not be mixed when comparing group means.

The Original SSF-PAQ suits large-scale surveys comparing groups on out-of-preschool screen time, night sleep, bedtime, and wake-up time, which correlated strongly with the criteria and showed good-to-excellent test–retest reliability. Its activity items showed good reliability and validity comparable to existing parent-report questionnaires (weak correlations with accelerometry);^11,14,36^ they can describe parent-reported exercise and active play and their settings. Reported time, shorter than accelerometer-measured PA (median, 73 versus 238 min/day; Table 2), should not be interpreted as PA duration. Activity days, mostly seven per week, were not significantly correlated with accelerometer-measured total PA. Outdoor-play days, which can be asked without duration estimates, correlated weakly with it, as did reported time, and showed good reliability; they are a candidate simple indicator. Both the SSF-PAQ and the Japanese MBQ-C^14^ cover exercise or play, screen time, and sleep and have been evaluated in Japanese preschoolers. Because the validity of the SSF-PAQ-24 activity items could not be confirmed here, the Japanese MBQ-C, which most parents rated as low in burden, is currently a candidate for whole-day assessment.

This study has several limitations. First, the screen-time criterion was parent-reported, and the sleep criterion was based on parent-reported diary data refined with accelerometer-derived bed-rest endpoints; therefore, shared parent-report bias between the questionnaire and criterion measures may have influenced the observed correlations. Additionally, the night-sleep criterion represented the sleep interval rather than actual sleep time and was not validated against an objective reference such as polysomnography. Second, the questionnaire’s recall period (the past month) did not match the one-week measurement window. Third, the short 3-day retest interval may have inflated reliability and underestimated measurement error due to participant recall. Fourth, convenience sampling limits these findings’ generalizability. Fifth, the SSF-PAQ-24 sample was small (*n* = 40) and differed from the Original-version sample in age, gender, and preschool type; thus, its estimates were imprecise and differences between versions cannot be attributed to question format alone. Future longitudinal studies should evaluate setting-specific validity and responsiveness to behavioural changes to determine the questionnaire’s utility for programme evaluation.

## Conclusion

In Japanese preschoolers, the Original SSF-PAQ showed good-to-excellent test–retest reliability for its main measures. Concurrent validity was supported for out-of-preschool screen time, night sleep, bedtime, and wake-up time, which correlated strongly with the criteria; validity for activity time, assessed against accelerometer-measured PA, was limited but comparable to that of existing parent-report questionnaires.

Because the SSF-PAQ-24 activity-item correlations were imprecise in a small sample (*n* = 40), their validity should be re-examined in a larger sample.

## ACKNOWLEDGMENTS

The authors thank the participating children, their parents, as well as the kindergartens, nursery schools, and centers for early childhood education and care that supported recruitment and data collection. During the preparation of this manuscript, the authors used a generative artificial intelligence tool (Claude; Anthropic) to assist with English-language editing. The authors reviewed and verified all output and take full responsibility for the content.

## Author contributions

K.K.: Conceptualization, funding acquisition, measurement and data analysis, and drafting of the manuscript. M.N. and R.T.: Development of the questionnaires. A.K.: Support for accelerometer-based data collection. S.I. and M.Y.: Methodological advice. All authors reviewed and edited the manuscript, and approved the final version.

## DATA AVAILABILITY

The raw data of the 2023 Sasakawa Sports Foundation national survey, from which the SSF-PAQ items were drawn, are available free of charge upon application to the Foundation.^13^ The accelerometer, diary, and questionnaire data generated and analysed in this validation study are available from the corresponding author upon reasonable request.

## SUPPLEMENTARY MATERIAL

The following supplementary materials are provided in a single combined file (Supplementary_Material.pdf).

eTable 1. STROBE Statement: checklist of items for cross-sectional studies

eMethods 1. Additional details of the methods

eTable 2. Allocation of participants to the six groups

eMaterial 1. SSF-PAQ (Original version) — paper, Japanese

eMaterial 2. SSF-PAQ (Original version) — paper, English translation

eMaterial 3. SSF-PAQ (Original version) — electronic, Japanese

eMaterial 4. SSF-PAQ (Original version) — electronic, English translation

eMaterial 5. SSF-PAQ-24 (24-hour version) — paper, Japanese

eMaterial 6. SSF-PAQ-24 (24-hour version) — paper, English translation

eMaterial 7. SSF-PAQ-24 (24-hour version) — electronic, Japanese

eMaterial 8. SSF-PAQ-24 (24-hour version) — electronic, English translation

eFigure 1. Bland–Altman plots comparing the SSF-PAQ with criterion measures for total screen time and total (night) sleep

eTable 3. Detailed concurrent validity (overall and within-/out-of-preschool) and test–retest reliability of the SSF-PAQ by version and administration mode

eTable 4. Test–retest reliability of setting-specific physical activity time in the SSF-PAQ

eTable 5. Measurement error of setting-specific physical activity time in the SSF-PAQ

eTable 6. Concurrent validity of the main behaviours by accelerometer wear-time criterion

eTable 7. Concurrent validity and test–retest reliability among children aged 3–5 years

eTable 8. Concurrent validity of the main behaviours under usual-day and diary-quality sensitivity analyses

eTable 9. Sensitivity of night sleep validity to the definition of weekend (free-day) nights

eTable 10. Validity, systematic differences, test–retest reliability, and measurement error of parent-reported sleep timing in the SSF-PAQ

eTable 11. Concurrent validity and test–retest reliability of day-based indicators of exercise and active play in the SSF-PAQ

