## Supplementary Material for "Validity and reliability of the SSF Preschoolers’ Activity Questionnaire (SSF-PAQ) for assessing active play, screen time, and sleep in Japanese preschool children"

**Affiliations**

### Index of supplementary material

#### eTable 1. STROBE Statement: checklist of items for cross-sectional studies

| No. | STROBE recommendation (abridged) | Status | Where reported / reason |
|---|---|---|---|
| **Title and abstract** | | | |
| 1a | Study design indicated with a common term in title or abstract | Partly reported | Title states 'Validity and reliability'; Abstract (Methods) describes the concurrent-validity design against accelerometry and a parent diary; the design term (cross-sectional validation study) appears only in Methods, 'Study design'. |
| 1b | Balanced, informative summary in the abstract | Reported | Structured Abstract (Background/Methods/Results/Conclusion). |
| **Introduction** | | | |
| 2 | Scientific background and rationale | Reported | Introduction, paragraphs 1–3 (24-hour movement guidelines and the need to know activity settings; limited evidence on the measurement properties of proxy-report questionnaires, including measurement error; the SSF-PAQ used in the 2023 national survey; differences in scope from the MBQ-C). |
| 3 | Specific objectives / prespecified aims | Reported | Introduction, final paragraph (primary aim: concurrent validity, test–retest reliability, measurement error, paper–electronic agreement, and practicality of the Original version; secondary exploratory aim: SSF-PAQ-24). |
| **Methods** | | | |
| 4 | Key elements of study design, presented early | Reported | Methods, 'Study design' (cross-sectional validation study sharing data collection with the MBQ-C validation in the same recruited sample). |
| 5 | Setting, locations, relevant dates incl. recruitment and data collection | Reported | 'Study design' (data collected May–June and October–December 2025); 'Participants and group allocation' (10 preschools in four Japanese cities); 'Study protocol' (10-day measurement protocol). |
| 6 | Eligibility criteria; sources and methods of participant selection | Reported | 'Study design' (written informed consent from a parent or guardian); 'Participants and group allocation' (children aged 3–6 years in 10 conveniently sampled preschools; no exclusion criteria; non-random allocation to six groups in roster order; eTable 2); eMethods 1. |
| 7 | Clearly define outcomes, exposures, predictors, modifiers | Reported | SSF-PAQ measures defined in 'The SSF-PAQ' (organized PA and AP, MVPA, screen time, night sleep) and criterion measures in 'Criterion measures'; index-versus-criterion framing rather than exposure/outcome (measurement-property study). |
| 8 | Sources of data and details of measurement; comparability of methods | Reported | 'Criterion measures' (accelerometry with machine-learning classification; 7-day time-use diary; sleep criterion integrating diary and accelerometer); 'The SSF-PAQ' (both versions in paper and electronic modes; eMaterials 1–8); details in eMethods 1; mode comparability analysed in 'Statistical analysis' and Table 5. |
| 9 | Efforts to address potential sources of bias | Reported | eMethods 1 (LINE reminders); 'Criterion measures' (criteria matched to each version's timeframe); 'Data cleaning and processing' and eMethods 1 (exclusion of implausible MVPA via Mahalanobis distance); sensitivity analyses using usual/accurately recorded diary days ('Statistical analysis'); shared reporting error addressed in Discussion (limitations). |
| 10 | How the study size was determined | Reported | 'Participants and group allocation' (a priori calculation for the Original-version activity items: $\rho = 0.25$, $\alpha = 0.05$, 80% power → 123 children with valid accelerometer data); assumptions and distribution target in eMethods 1; documents distributed for 623 children. |
| 11 | Handling of quantitative variables; groupings and rationale | Reported | 'Data cleaning and processing' (screen-time category midpoints; weekly average as (weekday × 5 + weekend/holiday × 2)/7; truncation at the original MBQ-C caps); interpretation thresholds in 'Statistical analysis'. |
| 12a | Statistical methods, including control for confounding | Reported | 'Statistical analysis' (Spearman ρ with Fisher z 95% CIs; ICC single-measurement absolute-agreement two-way mixed model; Wilcoxon signed-rank; Bland–Altman; mixed-model SEM, SDCind, SDCgroup). Confounder adjustment is not applicable to this measurement-property study. |
| 12b | Methods to examine subgroups and interactions | Reported | 'Statistical analysis' (weekday/weekend/weekly values; by version; by mode; setting-specific activity items; within- versus out-of-preschool play for the SSF-PAQ-24); detailed results in eTables 3–5. |

| No. | STROBE recommendation (abridged) | Status | Where reported / reason |
|---|---|---|---|
| 12c | How missing data were addressed | Reported | 'Statistical analysis' (no imputation apart from missing diary clock times in the night-sleep criterion; each analysis included all participants with the required variables, with n in each table); eMethods 1 (analysis sets; imputation of missing diary clock times from the child's mean for the same night type). |
| 12d | Methods accounting for sampling strategy | Reported | 'Participants and group allocation' (convenience sampling at preschool level; non-random allocation in roster order). No probability-sampling weights were applied. |
| 12e | Sensitivity analyses | Reported | 'Statistical analysis', final paragraph, and eMethods 1 (ages 3–5 years; wear-time criteria 360/600/960 min; usual-day and accurate-day diary subsets; calendar-weekend reclassification of criterion nights); Results, 'Sensitivity analyses'; eTables 6–9. |
| **Results** | | | |
| 13a | Numbers of individuals at each stage | Reported | Results, 'Participants' (208 consented → 171 with valid accelerometer data → 167 in the concurrent-validity sample); Figure 1 (183 questionnaire respondents and the n for each analysis). |
| 13b | Reasons for non-participation at each stage | Reported | Figure 1 (16 not measured, 2 device malfunction, 15 insufficient valid days, 4 implausible MVPA). |
| 13c | Flow diagram | Reported | Figure 1. |
| 14a | Characteristics of participants | Reported | Table 1; 'Participants' (age, gender; 24-hour version older, with more girls and more nursery-school children). |
| 14b | Number with missing data for each variable | Reported | 'Statistical analysis' (no imputation of analysis variables; each analysis included all participants with the required variables); number of missing values for each characteristic in Table 1; analysis-specific n in Tables 2–5 and the eTables. |
| 15 | Report outcome data / summary measures | Reported | Tables 2–5; Results, 'Concurrent validity' to 'Day-based indicators'; eTables 9–11. |
| 16a | Estimates and their precision (e.g., 95% CI) | Reported | ρ with 95% CIs (Tables 2 and 5); ICCs with 95% CIs (Tables 3 and 5); SEM/SDC (Table 4); Bland–Altman bias and limits of agreement ('Concurrent validity'; eFigure 1). |
| 16b | Category boundaries when continuous variables were categorized | Reported | 'Statistical analysis' (ρ thresholds per Schober 2018; ICC thresholds per Koo and Li 2016); 'Data cleaning and processing' (screen-time response categories); 'Practicality' and eMethods 1 (burden and clarity scales). |
| 16c | Translate relative risk into absolute risk if relevant | Not applicable | No risk or effect estimates are reported; this is a correlation/agreement (measurement-property) study without risk measures. |
| 17 | Other analyses (subgroups, interactions, sensitivity) | Reported | Results, 'Sensitivity analyses', 'Sleep timing', and 'Day-based indicators'; mode-, setting-, and version-specific results in eTables 3–5; sensitivity results in eTables 6–9; sleep timing in eTable 10; day-based indicators in eTable 11. |
| **Discussion** | | | |
| 18 | Summarise key results with reference to objectives | Reported | Discussion, paragraphs 1–2. |
| 19 | Limitations, including sources of bias/imprecision and their direction/magnitude | Reported | Discussion, limitations paragraph (shared reporting error in the parent-reported criteria for screen time and sleep; night-sleep criterion represents the sleep interval and was not tested against polysomnography; mismatch between the recall period and the measurement week; possible memory of previous answers with the 3-day retest interval; convenience sampling; small SSF-PAQ-24 sample of different composition). |
| 20 | Cautious overall interpretation | Reported | Discussion (behaviour-specific interpretation; activity items describe parent-reported exercise, play, and settings rather than PA duration; group-level interpretation of change based on SDCgroup; SSF-PAQ-24 results treated as exploratory). |
| 21 | Generalisability (external validity) | Reported | Discussion, limitations paragraph (convenience sampling). |
| **Other information** | | | |
| 22 | Source of funding and role of funders (present study; and original study if applicable) | Reported | Acknowledgments: Funding (DAIKO FOUNDATION FY2024 grant; the funder had no role) and Conflicts of interest (role of the Sasakawa Sports Foundation described). |

This validation study used a cross-sectional design. Each STROBE item is marked as Reported or Partly reported (with the manuscript location) or Not applicable (with a brief reason).

Reference: von Elm E, Altman DG, Egger M, Pocock SJ, Gøtzsche PC, Vandenbroucke JP; STROBE Initiative. The Strengthening the Reporting of Observational Studies in Epidemiology (STROBE) statement. Lancet. 2007;370(9596):1453–1457.

#### eMethods 1. Additional details of the methods

**Sample size and recruitment.** The sample size was based on the concurrent validity of the Original version. It was calculated for the activity items because, of the three behaviours, activity items have shown the weakest correlations with criterion measures (reference 11 of the main text). Earlier accelerometer validations of parent-reported play reported correlations of 0.20–0.39 (references 11 and 16). Assuming $\rho = 0.25$, 123 children with valid accelerometer data were required ($\alpha = 0.05$, 80% power; R package pwr). Assuming a consent rate of 30% and 20% missing data (reference 17), documents needed to be distributed for about 513 children. Because we also planned to collect SSF-PAQ-24 data, documents were distributed for more children where practicable. Eleven preschools were selected by convenience sampling and one withdrew beforehand; documents were distributed to the parents of 623 children at the remaining 10 preschools. Participants received an honorarium of up to 3,000 Japanese yen.

**Group allocation.** Consenting children were entered in a participant roster ordered by preschool and grade and were allocated in roster order by rotating a fixed sequence, without randomization, so that the groups were spread across preschools and grades. Because data collection was shared with the companion MBQ-C validation study (reference 14), the rotation followed the seven groups of that study, which combined the format and mode of the MBQ-C with the mode of the SSF-PAQ. For the SSF-PAQ, two of the seven groups completed the paper mode at both administrations (G1), two completed the electronic mode at both (G2), and three switched modes (G3; two completed the paper mode first and one the electronic mode first). The resulting sequence was therefore G1, G2, G1, G2, G3, G3, G3, and two-thirds of G3 were allocated to the paper-first order. The first 161 children (four preschools; May–June and October–November 2025) were allocated to the Original version and the subsequent 47 children (seven preschools, including one that also took part in the Original version; October–December 2025) to the SSF-PAQ-24. For the SSF-PAQ-24, the sequence was initially G4, G5, G4, G5, G6. Allocation to G6 was discontinued after three children because an adequate sample for assessing paper–electronic agreement was unlikely to be obtained, and priority was given to test–retest reliability in each mode; the sequence thereafter was G4, G5, G4, G5. One child allocated to G1 completed the Day 10 questionnaire in the electronic mode and was analysed in G3.

**Study protocol.** Through the preschools, each family received the accelerometer, the 7-day time-use diary, the questionnaires, and a QR code for reminders sent through the LINE messaging app. The kits were collected through the preschools.

**Accelerometer processing.** The accelerometer sampled at 100 Hz. Because machine-learning classification of raw data assesses young children's activity more accurately than cut-point methods (references 18 and 19), raw data were classified with the Preschool Hip Random Forest free-living classifier (reference 18) in the R package actimetric (reference 20). Because a processing issue affecting 100-Hz data starting at 00:00 was identified in the package, the .gt3x files were processed directly with the underlying R script after consultation with the classifier developer (the issue has since been corrected in actimetric; reference 14). Non-wear time was identified with the vector-magnitude standard-deviation algorithm (reference 21). Mean Euclidean Norm Minus One (ENMO; reference 22) was calculated over wear epochs. Out-of-preschool time was identified from the preschool arrival and departure times recorded in the diary.

**Time-use diary.** The diary was based on the criterion diary of the Australian MBQ-C validation study (reference 11), which was adapted from the diary of Mendoza et al. (reference 23). Parents recorded sleep, passive and interactive screen time, standing time during screen use, and accelerometer non-wear in 15-minute units, entered preschool arrival and departure times, and rated how usual each day was and how accurately it had been recorded. These ratings were used in the sensitivity analyses. The SSF-PAQ screen-time item asks parents to exclude active video games and time spent being physically active while watching videos but does not specify how viewing while standing still should be reported. In the diary, parents ticked the 15-minute units in which the child used screens while standing, without distinguishing standing still from moving; all standing screen time was therefore excluded from the criterion.

**Night-sleep criterion.** Sleep intervals defined by diary bedtime and wake-up time were combined with the onset and end of bed rest estimated by an algorithm developed for hip-worn accelerometer data in young children (reference 24). Missing diary clock times were imputed with the child's own mean for the same night type. Nights were classified according to whether the following day was a day off because children may go to bed and get up later before a day off. Diary and sleep processing are described further in the companion paper (reference 14).

**Practicality items.** Completion time was calculated from the clock times that parents entered at the start and end of the questionnaire. Perceived response burden was rated on a four-point scale ('hardly burdensome' to 'very burdensome') and perceived clarity on a five-point scale ('very clear' to 'very unclear').

**Data cleaning.** The 07:00–21:00 window covers the main waking hours of Japanese preschoolers (reference 13), and the requirement of ≥ 600 minutes of wear is consistent with the common 10-hour criterion (reference 25). Children with implausibly high MVPA relative to their step counts were excluded when the Mahalanobis distance for weekly steps and MVPA exceeded the 99.9th percentile of the $\chi^2$ distribution with 2 degrees of freedom. Further details of accelerometer processing are given in the companion paper (reference 14). Truncation of the activity values was applied only to the weekend/holiday values of three Original-version children (five responses). SSF-PAQ screen time is asked in seven categories (‘less than 30 min’ to ‘5 h or more’), which were converted to category midpoints (15–270 minutes; 300 minutes for the top category).

**Statistical software and analysis sets.** Confidence intervals based on the Fisher z transformation were obtained with the DescTools package (reference 28) and ICCs with the irr package (reference 30). Bland–Altman analysis was used to assess bias, limits of agreement, and proportional bias. Participant characteristics were summarized as means and standard deviations or as numbers and percentages. Concurrent validity required valid accelerometer data and a Day 7 response; reliability, measurement error, and paper–electronic agreement required responses on both Day 7 and Day 10; and practicality required a Day 7 response. For social jetlag, positive values indicate later midsleep on weekends.

**Day-based indicators.** The 2023 national survey reports the number of days per week on which preschoolers play outdoors outside of preschool hours (reference 13). Two day-based indicators were therefore derived from the Original version: activity days, the number of days of the week (0–7) on which the parent reported at least 1 minute of organized lessons, outdoor active play, or indoor active play; and outdoor-play days, the number of days with at least 1 minute of outdoor active play. Days with organized lessons and days with indoor active play were also counted (eTable 11). In the electronic mode, parents first selected the days on which each activity took place and then entered its duration; in the paper mode, they entered a duration for each day of the week. Concurrent validity was assessed with the Spearman correlation between the Day 7 questionnaire and accelerometer-derived out-of-preschool total PA (weekly average; correlations with MVPA and mean ENMO are also shown in eTable 11), and test–retest reliability with the ICC between Days 7 and 10 in the same mode. Because an accelerometer cannot identify the days on which a child played, the accuracy of the reported number of days itself was not examined.

**Sensitivity analyses.** The stricter wear-time criterion was ≥ 960 minutes of wear over 24 hours, and the more lenient criterion was ≥ 360 minutes between 07:00 and 21:00, because ≥ 6 hours of wear on any 3 days has been reported to give acceptable reliability in young children (reference 35). The criteria were recalculated from days reported as usual, days rated as accurately recorded, or both, because the SSF-PAQ asks about usual behaviour over the past month, whereas the diary-based criteria depend on the quality of recording. For night sleep, the SSF-PAQ asks for bedtime and wake-up time on weekdays (Monday to Friday) and on weekend days (Saturday and Sunday). A weekend bedtime can be understood as the bedtime on the night before a weekend day or as the bedtime on Saturday and Sunday nights, so criterion weekend nights were also reclassified by the calendar day on which the night began (Saturday and Sunday nights).

#### eTable 2. Allocation of participants to the six groups

| Group | n | Day 0 | Days 1–7 (T1) | Day 7 (T2) | Day 10 (T3) |
|---|---|---|---|---|---|
| G1 | 45 | Received the accelerometer and diary | Accelerometer monitoring and diary recording | SSF-PAQ Original (paper) | SSF-PAQ Original (paper) |
| G2 | 46 | | | SSF-PAQ Original (electronic) | SSF-PAQ Original (electronic) |
| G3 | 70 | | | SSF-PAQ Original (paper or electronic) | SSF-PAQ Original (the other mode) |
| G4 | 22 | | | SSF-PAQ-24 (paper) | SSF-PAQ-24 (paper) |
| G5 | 22 | | | SSF-PAQ-24 (electronic) | SSF-PAQ-24 (electronic) |
| G6 | 3 | | | SSF-PAQ-24 (paper or electronic) | SSF-PAQ-24 (the other mode) |

| Analysis | n | Questionnaire data | Groups (n) |
|---|---|---|---|
| Concurrent validity (SSF-PAQ Original) | 161 | Day 7 | G1 (45), G2 (46), G3 (70) |
| Concurrent validity (SSF-PAQ-24) | 47 | Day 7 | G4 (22), G5 (22), G6 (3) |
| Test–retest reliability and measurement error (SSF-PAQ Original) | 91 | Day 7 and Day 10 | G1 (45), G2 (46) |
| Test–retest reliability and measurement error (SSF-PAQ-24) | 44 | Day 7 and Day 10 | G4 (22), G5 (22) |
| Paper–electronic agreement (SSF-PAQ Original) | 70 | Day 7 and Day 10 | G3 (70) |

Values are the number of consenting participants in each group (n = 208). One child allocated to G1 completed the Day 10 questionnaire in the electronic mode and was included in G3.

T1, accelerometer-based assessment of physical activity together with diary-based recording of screen time and sleep (Days 1–7); T2, first questionnaire administration (Day 7); T3, second questionnaire administration (Day 10), using the same questionnaire version with either the same administration mode (G1, G2, G4, G5) or the other mode (G3, G6). The concurrent-validity analyses used the accelerometer-derived measures from T1 and the questionnaire data from T2; the other analyses used the questionnaire data from T2 and T3. Practicality (completion time, burden, and clarity) was assessed in all groups at T2.

SSF-PAQ, Sasakawa Sports Foundation Preschoolers' Activity Questionnaire; SSF-PAQ-24, 24-hour version of the SSF-PAQ. Paper and electronic denote the administration mode. Paper–electronic agreement for the 24-hour version (G6, n = 3) is not reported because of the small sample.

eMaterial 1. SSF-PAQ (Original version) — paper, Japanese

### 幼児の身体活動、睡眠、スクリーンタイムに関する質問票

#### SSF Preschoolers' Activity Questionnaire (SSF-PAQ)

**質問1．** ここ1ヶ月について、お子様は運動の習いごとをしていますか？（幼稚園や保育園などにいる時間は含めません。ただし、園が終了してから、園の施設や場所を使って家庭が任意で習わせる運動の時間は含みます）（ひとつだけ）

[1] はい　　[2] いいえ　→ **質問2へ**

**質問 1_1.** お子様の運動の習いごとについて、ここ1ヶ月を思い出し、曜日ごとに平均的な1日あたりの時間（分）を回答してください。**運動の習いごとをしていない曜日は「0分」と記入してください。**

| | 月曜日 | 火曜日 | 水曜日 | 木曜日 | 金曜日 | 土曜日 | 日曜日 |
|---|---|---|---|---|---|---|---|
| 運動の習いごとをしている時間 | 分 | 分 | 分 | 分 | 分 | 分 | 分 |

**質問 1_3.** これらの運動の習いごとのうち、息がはずむ程度のきつさの運動時間は何分くらいありますか？**ない場合は「0分」と記入してください。**

| | 月曜日 | 火曜日 | 水曜日 | 木曜日 | 金曜日 | 土曜日 | 日曜日 |
|---|---|---|---|---|---|---|---|
| 息がはずむ程度のきつさの運動時間 | 分 | 分 | 分 | 分 | 分 | 分 | 分 |

**ここからは、お子様が幼稚園・保育園・運動の習いごと以外で過ごす「体を動かして遊ぶ時間」についての質問です。**

**質問 2.** 園にいる時や運動の習いごと以外で、お子様はどれくらいの時間、体を動かす外遊びをしていますか？ここ 1 ヶ月を思い出し、各曜日の遊んでいる場所ごとに、平均的な 1 日あたりの時間（分）を回答してください。
例：日曜日に公園で 1 時間半遊んでいる場合、日曜日の「公園や屋外の遊び場」の欄に「90」と入力してください。**外で体を動かしていない曜日は「0分」と記入してください。**

| | 月曜日 | 火曜日 | 水曜日 | 木曜日 | 金曜日 | 土曜日 | 日曜日 |
|---|---|---|---|---|---|---|---|
| 公園や屋外の遊び場 | 分 | 分 | 分 | 分 | 分 | 分 | 分 |
| 自宅の敷地内（庭など） | 分 | 分 | 分 | 分 | 分 | 分 | 分 |
| 路上（道路や路地裏など） | 分 | 分 | 分 | 分 | 分 | 分 | 分 |
| その他の外遊びの場所（　　　　　　　） | 分 | 分 | 分 | 分 | 分 | 分 | 分 |

**質問 2_1.** これらの外遊びのうち、息がはずむ程度のきつさの運動時間は何分くらいありますか？**ない場合は「0分」と記入してください。**

| | 月曜日 | 火曜日 | 水曜日 | 木曜日 | 金曜日 | 土曜日 | 日曜日 |
|---|---|---|---|---|---|---|---|
| 息がはずむ程度のきつさの運動時間 | 分 | 分 | 分 | 分 | 分 | 分 | 分 |

**質問 3.** 園にいる時や運動の習いごと以外で、お子様はどれくらいの時間、室内や屋内施設で体を動かす遊びをしていますか？ここ 1 ヶ月を思い出し、曜日ごとに平均的な 1 日あたりの時間（分）を回答してください。**室内で体を動かして遊ばない曜日は「0 分」と記入してください。**

| | 月曜日 | 火曜日 | 水曜日 | 木曜日 | 金曜日 | 土曜日 | 日曜日 |
|---|---|---|---|---|---|---|---|
| 室内でゲーム機や動画を見ながら体を動かす運動（Wii Sports、ニンテンドースイッチなどのゲーム、YouTube など） | 分 | 分 | 分 | 分 | 分 | 分 | 分 |
| 室内で体を動かす運動（ゲーム機や動画以外。走り回る、親子で体を動かして遊ぶなど） | 分 | 分 | 分 | 分 | 分 | 分 | 分 |
| 屋内運動施設（例 体育館・プール、児童館、商業施設の遊び場など） | 分 | 分 | 分 | 分 | 分 | 分 | 分 |
| その他の室内で体を動かす場面（ ） | 分 | 分 | 分 | 分 | 分 | 分 | 分 |

**質問 3_1.** これらの室内や屋内施設で体を動かす遊びのうち、息がはずむ程度のきつさの運動時間は何分くらいありますか？**ない場合は「0 分」と記入してください。**

| | 月曜日 | 火曜日 | 水曜日 | 木曜日 | 金曜日 | 土曜日 | 日曜日 |
|---|---|---|---|---|---|---|---|
| 息がはずむ程度のきつさの運動時間 | 分 | 分 | 分 | 分 | 分 | 分 | 分 |

#### お子様の生活習慣についてお答えください

**質問 4.** お子様の平日（月～金）と休日（土・日）の就寝時刻と起床時刻を 24 時間表記で教えてください。（例：午後 10 時⇒22 時）

| | 就寝時刻 | | 起床時刻 | |
|---|---|---|---|---|
| 平日 | 時 | 分 | 時 | 分 |
| 休日 | 時 | 分 | 時 | 分 |

**質問 5.** ここ 1 ヶ月、お子様は、園（幼稚園・保育園など含む）の活動以外で、テレビや DVD をみたり、パソコン、ゲーム（テレビ、パソコン、携帯式のゲーム機などを含む）、スマートフォンなどを使用したりする時間は 1 日あたりどのくらいですか。（ただし、Wii Sports などの体を動かすゲーム、YouTube などの動画を見ながら体を動かす時間は含めません）

| | 30 分未満 | 30 分～1 時間未満 | 1～2 時間未満 | 2～3 時間未満 | 3～4 時間未満 | 4～5 時間未満 | 5 時間以上 |
|---|---|---|---|---|---|---|---|
| 平日（月～金） | [1] | [2] | [3] | [4] | [5] | [6] | [7] |
| 休日（土・日） | [1] | [2] | [3] | [4] | [5] | [6] | [7] |

ご回答、ありがとうございました。

**eMaterial 2. SSF-PAQ (Original version) — paper, English translation**

*Note: This English version was translated by the authors to help readers understand the original Japanese questionnaire. It has not undergone a formal translation and cross-cultural adaptation process.*

#### Questionnaire on Physical Activity, Sleep, and Screen Time in Preschool Children

### SSF Preschoolers' Activity Questionnaire (SSF-PAQ)

**Q1.** Over the past month, has your child taken any organized sports or exercise lessons? (Do not include time spent at preschool: kindergarten, nursery school, or center for early childhood education and care. However, do include organized sports or exercise lessons that your family voluntarily arranges after preschool hours using the preschool's facilities or grounds.) (Select one)

[1] Yes [2] No → **Go to Question 2**

**Q1_1.** Thinking back over the past month, please enter your child's average daily time (minutes) spent in organized sports or exercise lessons for each day of the week. **Enter "0" for days with no lessons.**

| | Mon | Tue | Wed | Thu | Fri | Sat | Sun |
|---|---|---|---|---|---|---|---|
| Time spent in organized sports or exercise lessons | min | min | min | min | min | min | min |

**Q1_3.** Of these lessons, about how many minutes per day were spent in activity that makes your child breathe harder? **Enter "0" if none.**

| | Mon | Tue | Wed | Thu | Fri | Sat | Sun |
|---|---|---|---|---|---|---|---|
| Time in activity that makes your child breathe harder | min | min | min | min | min | min | min |

**The following questions ask about your child's active play time outside of preschool and organized sports or exercise lessons.**

**Q2.** Outside of preschool and organized sports or exercise lessons, how much time does your child spend in outdoor active play? Thinking back over the past month, please enter the average daily time (minutes) for each play location on each day of the week.

Example: If your child plays at a park for an hour and a half on Sundays, enter "90" in the "Parks or outdoor playgrounds" row for Sunday. **Enter "0" for days with no outdoor active play.**

| | Mon | Tue | Wed | Thu | Fri | Sat | Sun |
|---|---|---|---|---|---|---|---|
| Parks or outdoor playgrounds | min | min | min | min | min | min | min |
| Home premises (yard, etc.) | min | min | min | min | min | min | min |
| Streets (roads, alleys, etc.) | min | min | min | min | min | min | min |
| Other outdoor play locations (________) | min | min | min | min | min | min | min |

**Q2_1.** Of this outdoor play, about how many minutes per day were spent in activity that makes your child breathe harder? **Enter "0" if none.**

| | Mon | Tue | Wed | Thu | Fri | Sat | Sun |
|---|---|---|---|---|---|---|---|
| Time in activity that makes your child breathe harder | min | min | min | min | min | min | min |

**Q3.** Outside of preschool and organized sports or exercise lessons, how much time does your child spend in active play indoors or at indoor facilities? Thinking back over the past month, please enter the average daily time (minutes) for each day of the week. **Enter "0" for days with no indoor active play.**

| | Mon | Tue | Wed | Thu | Fri | Sat | Sun |
|---|---|---|---|---|---|---|---|
| Active play indoors with video games or videos (e.g., Wii Sports, Nintendo Switch games, YouTube) | min | min | min | min | min | min | min |
| Active play indoors without games or videos (e.g., running around, active play with parents) | min | min | min | min | min | min | min |
| Indoor exercise facilities (e.g., gymnasium, swimming pool, children's center, indoor playgrounds in commercial facilities) | min | min | min | min | min | min | min |
| Other indoor active play situations (________) | min | min | min | min | min | min | min |

**Q3_1.** Of this indoor play, about how many minutes per day were spent in activity that makes your child breathe harder? **Enter "0" if none.**

| | Mon | Tue | Wed | Thu | Fri | Sat | Sun |
|---|---|---|---|---|---|---|---|
| Time in activity that makes your child breathe harder | min | min | min | min | min | min | min |

**Please answer the following questions about your child's daily routine.**

**Q4.** Please indicate your child's bedtime and wake-up time on weekdays (Mon–Fri) and weekend days (Sat–Sun), using the 24-hour clock (e.g., 10 p.m. → 22:00).

| | Bedtime | | Wake-up time | |
|---|---|---|---|---|
| Weekdays | h | min | h | min |
| Weekend days | h | min | h | min |

**Q5.** Over the past month, outside of preschool activities, about how much time per day has your child spent watching TV or DVDs, or using computers, video games (including TV, computer, and handheld game consoles), smartphones, and similar devices? (Do not include active video games such as Wii Sports or time spent being physically active while watching videos such as YouTube.)

| | Less than 30 min | 30 min to <1 h | 1 to <2 h | 2 to <3 h | 3 to <4 h | 4 to <5 h | 5 h or more |
|---|---|---|---|---|---|---|---|
| Weekdays (Mon–Fri) | [1] | [2] | [3] | [4] | [5] | [6] | [7] |
| Weekend days (Sat–Sun) | [1] | [2] | [3] | [4] | [5] | [6] | [7] |

**Thank you for completing this questionnaire.**

### 幼児の身体活動、睡眠、スクリーンタイムに関する質問票

#### SSF Preschoolers' Activity Questionnaire (SSF-PAQ)

**質問１.** ここ１ヶ月について、お子様は運動の習いごとをしていますか？（幼稚園や保育園などにいる時間は含めません。ただし、園が終了してから、園の施設や場所を使って家庭が任意で習わせる運動の時間は含みます）（ひとつだけ）

○　はい
○　いいえ　【条件】『いいえ』を選択した場合は質問２へ

**質問１_1.** お子様が運動の習いごとをしている曜日をすべてお答えください。（あてはまるものすべて）

☐月曜日　☐火曜日　☐水曜日　☐木曜日　☐金曜日　☐土曜日　☐日曜日

**質問 1_2.** お子様の運動の習いごとについて、ここ１ヶ月を思い出し、曜日ごとに平均的な１日あたりの時間（分）を回答してください。
※1～480 の範囲でお答えください。※半角でご入力ください。

月曜日[　　]分　【条件】質問 1_1 で『月曜日』を選択した場合。入力値は 1～480。

火曜日[　　]分　【条件】質問 1_1 で『火曜日』を選択した場合。入力値は 1～480。

水曜日[　　]分　【条件】質問 1_1 で『水曜日』を選択した場合。入力値は 1～480。

木曜日[　　]分　【条件】質問 1_1 で『木曜日』を選択した場合。入力値は 1～480。

金曜日[　　]分　【条件】質問 1_1 で『金曜日』を選択した場合。入力値は 1～480。

土曜日[　　]分　【条件】質問 1_1 で『土曜日』を選択した場合。入力値は 1～480。

日曜日[　　]分　【条件】質問 1_1 で『日曜日』を選択した場合。入力値は 1～480。

**質問 1_3.** これらの運動の習いごとのうち、息がはずむ程度のきつさの運動時間は何分くらいありますか？ **ない場合は「0 分」と入力してください。**
※0～480 の範囲でお答えください。※半角でご入力ください。

月曜日[　　]分　【条件】質問 1_1 で『月曜日』を選択した場合。入力値は 0～480。

火曜日[　　]分　【条件】質問 1_1 で『火曜日』を選択した場合。入力値は 0～480。

水曜日[　　]分　【条件】質問 1_1 で『水曜日』を選択した場合。入力値は 0～480。

木曜日[　　]分　【条件】質問 1_1 で『木曜日』を選択した場合。入力値は 0～480。

金曜日[　　]分　【条件】質問 1_1 で『金曜日』を選択した場合。入力値は 0～480。

土曜日[　　]分　【条件】質問 1_1 で『土曜日』を選択した場合。入力値は 0～480。

日曜日[　　]分　【条件】質問 1_1 で『日曜日』を選択した場合。入力値は 0～480。

**ここからは、お子様が幼稚園・保育園・運動の習いごと以外で過ごす「体を動かして遊ぶ時間」についての質問です。**

**質問 2.** 園にいる時や運動の習いごと以外で、お子様は体を動かす外遊びをしていますか？ここ 1 ヶ月を思い出し、曜日ごとに遊んでいる場所をすべて選択してください。**体を動かす外遊びをしない曜日は「この曜日は体を動かす外遊びをしていない」を選択してください。**

＜月曜日＞
☐ 公園や屋外の遊び場
☐ 自宅の敷地内（庭など）
☐ 路上（道路や路地裏など）
☐ その他の外遊びの場所
☐ この曜日は体を動かす外遊びをしていない

＜火曜日＞
☐ 公園や屋外の遊び場
☐ 自宅の敷地内（庭など）
☐ 路上（道路や路地裏など）
☐ その他の外遊びの場所
☐ この曜日は体を動かす外遊びをしていない

＜水曜日＞
☐ 公園や屋外の遊び場
☐ 自宅の敷地内（庭など）
☐ 路上（道路や路地裏など）
☐ その他の外遊びの場所
☐ この曜日は体を動かす外遊びをしていない

＜木曜日＞
☐ 公園や屋外の遊び場
☐ 自宅の敷地内（庭など）
☐ 路上（道路や路地裏など）
☐ その他の外遊びの場所
☐ この曜日は体を動かす外遊びをしていない

＜金曜日＞
☐ 公園や屋外の遊び場
☐ 自宅の敷地内（庭など）
☐ 路上（道路や路地裏など）
☐ その他の外遊びの場所
☐ この曜日は体を動かす外遊びをしていない

＜土曜日＞
☐ 公園や屋外の遊び場
☐ 自宅の敷地内（庭など）
☐ 路上（道路や路地裏など）
☐ その他の外遊びの場所
☐ この曜日は体を動かす外遊びをしていない

＜日曜日＞
☐ 公園や屋外の遊び場
☐ 自宅の敷地内（庭など）
☐ 路上（道路や路地裏など）
☐ その他の外遊びの場所
☐ この曜日は体を動かす外遊びをしていない

**質問 2_1.** 園にいる時や運動の習いごと以外で、お子様はどれくらいの時間、体を動かす外遊びをしていますか？ここ 1 ヶ月を思い出し、各曜日の遊んでいる場所ごとに、平均的な 1 日あたりの時間（分）を回答してください。
例：日曜日に公園で 1 時間半遊んでいる場合、日曜日の「公園や屋外の遊び場」の欄に「90」と入力してください。
※1～480 の範囲でお答えください。※半角でご入力ください。

＜月曜日＞
公園や屋外の遊び場
分　【条件】質問 2 で『月曜日』の該当項目を選択した場合。入力値は 1～480。
自宅の敷地内（庭など）
分　【条件】質問 2 で『月曜日』の該当項目を選択した場合。入力値は 1～480。
路上（道路や路地裏など）
分　【条件】質問 2 で『月曜日』の該当項目を選択した場合。入力値は 1～480。
その他の外遊びの場所
分　【条件】質問 2 で『月曜日』の該当項目を選択した場合。入力値は 1～480。

＜火曜日＞
公園や屋外の遊び場
分　【条件】質問 2 で『火曜日』の該当項目を選択した場合。入力値は 1～480。
自宅の敷地内（庭など）
分　【条件】質問 2 で『火曜日』の該当項目を選択した場合。入力値は 1～480。
路上（道路や路地裏など）
分　【条件】質問 2 で『火曜日』の該当項目を選択した場合。入力値は 1～480。
その他の外遊びの場所
分　【条件】質問 2 で『火曜日』の該当項目を選択した場合。入力値は 1～480。

＜水曜日＞
公園や屋外の遊び場
分　【条件】質問 2 で『水曜日』の該当項目を選択した場合。入力値は 1～480。
自宅の敷地内（庭など）
分　【条件】質問 2 で『水曜日』の該当項目を選択した場合。入力値は 1～480。
路上（道路や路地裏など）
分　【条件】質問 2 で『水曜日』の該当項目を選択した場合。入力値は 1～480。
その他の外遊びの場所
分　【条件】質問 2 で『水曜日』の該当項目を選択した場合。入力値は 1～480。

＜木曜日＞
公園や屋外の遊び場
分　【条件】質問 2 で『木曜日』の該当項目を選択した場合。入力値は 1～480。
自宅の敷地内（庭など）
分　【条件】質問 2 で『木曜日』の該当項目を選択した場合。入力値は 1～480。
路上（道路や路地裏など）
分　【条件】質問 2 で『木曜日』の該当項目を選択した場合。入力値は 1～480。
その他の外遊びの場所
分　【条件】質問 2 で『木曜日』の該当項目を選択した場合。入力値は 1～480。

＜金曜日＞
公園や屋外の遊び場
分　【条件】質問 2 で『金曜日』の該当項目を選択した場合。入力値は 1～480。
自宅の敷地内（庭など）
分　【条件】質問 2 で『金曜日』の該当項目を選択した場合。入力値は 1～480。
路上（道路や路地裏など）
分　【条件】質問 2 で『金曜日』の該当項目を選択した場合。入力値は 1～480。

その他の外遊びの場所
[ ]分 【条件】質問 2 で『金曜日』の該当項目を選択した場合。入力値は 1～480。

＜土曜日＞
公園や屋外の遊び場
[ ]分 【条件】質問 2 で『土曜日』の該当項目を選択した場合。入力値は 1～480。
自宅の敷地内（庭など）
[ ]分 【条件】質問 2 で『土曜日』の該当項目を選択した場合。入力値は 1～480。
路上（道路や路地裏など）
[ ]分 【条件】質問 2 で『土曜日』の該当項目を選択した場合。入力値は 1～480。
その他の外遊びの場所
[ ]分 【条件】質問 2 で『土曜日』の該当項目を選択した場合。入力値は 1～480。

＜日曜日＞
公園や屋外の遊び場
[ ]分 【条件】質問 2 で『日曜日』の該当項目を選択した場合。入力値は 1～480。
自宅の敷地内（庭など）
[ ]分 【条件】質問 2 で『日曜日』の該当項目を選択した場合。入力値は 1～480。
路上（道路や路地裏など）
[ ]分 【条件】質問 2 で『日曜日』の該当項目を選択した場合。入力値は 1～480。
その他の外遊びの場所
[ ]分 【条件】質問 2 で『日曜日』の該当項目を選択した場合。入力値は 1～480。

**質問 2_2.** これらの外遊びのうち、息がはずむ程度のきつさの運動時間は何分くらいありますか？**ない場合は「0 分」と入力してください。**
※0～480 の範囲でお答えください。※半角でご入力ください。

月曜日[ ]分 【条件】質問 2 で『月曜日』の遊び場所を選択した場合。入力値は 0～480。

火曜日[ ]分 【条件】質問 2 で『火曜日』の遊び場所を選択した場合。入力値は 0～480。

水曜日[ ]分 【条件】質問 2 で『水曜日』の遊び場所を選択した場合。入力値は 0～480。

木曜日[ ]分 【条件】質問 2 で『木曜日』の遊び場所を選択した場合。入力値は 0～480。

金曜日[ ]分 【条件】質問 2 で『金曜日』の遊び場所を選択した場合。入力値は 0～480。

土曜日[ ]分 【条件】質問 2 で『土曜日』の遊び場所を選択した場合。入力値は 0～480。

日曜日[ ]分 【条件】質問 2 で『日曜日』の遊び場所を選択した場合。入力値は 0～480。

**質問 3.** 園にいる時や運動の習いごと以外で、お子様は室内や屋内施設で体を動かす遊びをしていますか？ここ１ヶ月を思い出し、曜日ごとに遊んでいる項目をすべて選択してください。**室内で体を動かして遊ばない曜日は「この曜日は室内で体を動かして遊ばない」を選択してください。**

＜月曜日＞
☐ 室内でゲーム機や動画を見ながら体を動かす運動（例：Wii Sports、ニンテンドースイッチなどのゲーム、YouTube など）
☐ 室内で体を動かす運動（例：ゲーム機や動画以外。走り回る、親子で体を動かして遊ぶなど）
☐ 屋内運動施設（例：体育館・プール、児童館、商業施設の遊び場など）
☐ その他の室内で体を動かす場面
☐ この曜日は室内で体を動かして遊ばない

＜火曜日＞
☐ 室内でゲーム機や動画を見ながら体を動かす運動（例：Wii Sports、ニンテンドースイッチなどのゲーム、YouTube など）
☐ 室内で体を動かす運動（例：ゲーム機や動画以外。走り回る、親子で体を動かして遊ぶなど）
☐ 屋内運動施設（例：体育館・プール、児童館、商業施設の遊び場など）
☐ その他の室内で体を動かす場面
☐ この曜日は室内で体を動かして遊ばない

＜水曜日＞
☐ 室内でゲーム機や動画を見ながら体を動かす運動（例：Wii Sports、ニンテンドースイッチなどのゲーム、YouTube など）
☐ 室内で体を動かす運動（例：ゲーム機や動画以外。走り回る、親子で体を動かして遊ぶなど）
☐ 屋内運動施設（例：体育館・プール、児童館、商業施設の遊び場など）
☐ その他の室内で体を動かす場面
☐ この曜日は室内で体を動かして遊ばない

＜木曜日＞
☐ 室内でゲーム機や動画を見ながら体を動かす運動（例：Wii Sports、ニンテンドースイッチなどのゲーム、YouTube など）
☐ 室内で体を動かす運動（例：ゲーム機や動画以外。走り回る、親子で体を動かして遊ぶなど）
☐ 屋内運動施設（例：体育館・プール、児童館、商業施設の遊び場など）
☐ その他の室内で体を動かす場面
☐ この曜日は室内で体を動かして遊ばない

＜金曜日＞
☐ 室内でゲーム機や動画を見ながら体を動かす運動（例：Wii Sports、ニンテンドースイッチなどのゲーム、YouTube など）
☐ 室内で体を動かす運動（例：ゲーム機や動画以外。走り回る、親子で体を動かして遊ぶなど）
☐ 屋内運動施設（例：体育館・プール、児童館、商業施設の遊び場など）
☐ その他の室内で体を動かす場面
☐ この曜日は室内で体を動かして遊ばない

＜土曜日＞
☐ 室内でゲーム機や動画を見ながら体を動かす運動（例：Wii Sports、ニンテンドースイッチなどのゲーム、YouTube など）
☐ 室内で体を動かす運動（例：ゲーム機や動画以外。走り回る、親子で体を動かして遊ぶなど）
☐ 屋内運動施設（例：体育館・プール、児童館、商業施設の遊び場など）
☐ その他の室内で体を動かす場面
☐ この曜日は室内で体を動かして遊ばない

＜日曜日＞
☐ 室内でゲーム機や動画を見ながら体を動かす運動（例：Wii Sports、ニンテンドースイッチなどのゲーム、YouTube など）
☐ 室内で体を動かす運動（例：ゲーム機や動画以外。走り回る、親子で体を動かして遊ぶなど）
☐ 屋内運動施設（例：体育館・プール、児童館、商業施設の遊び場など）
☐ その他の室内で体を動かす場面
☐ この曜日は室内で体を動かして遊ばない

**質問 3_1.** 園にいる時や運動の習いごと以外で、お子様はどれくらいの時間、室内や屋内施設で体を動かす遊びをしていますか？ここ 1 ヶ月を思い出し、各曜日の遊んでいる項目ごとに、平均的な 1 日あたりの時間（分）を回答してください。
※1～480 の範囲でお答えください。※半角でご入力ください。

＜月曜日＞

室内でゲーム機や動画を見ながら体を動かす運動（例：Wii Sports、ニンテンドースイッチなどのゲーム、YouTube など）
□分　【条件】質問 3 で『月曜日』の該当項目を選択した場合。入力値は 1～480。
室内で体を動かす運動（例：ゲーム機や動画以外。走り回る、親子で体を動かして遊ぶなど）
□分　【条件】質問 3 で『月曜日』の該当項目を選択した場合。入力値は 1～480。
屋内運動施設（例：体育館・プール、児童館、商業施設の遊び場など）
□分　【条件】質問 3 で『月曜日』の該当項目を選択した場合。入力値は 1～480。
その他の室内で体を動かす場面
□分　【条件】質問 3 で『月曜日』の該当項目を選択した場合。入力値は 1～480。

＜火曜日＞
室内でゲーム機や動画を見ながら体を動かす運動（例：Wii Sports、ニンテンドースイッチなどのゲーム、YouTube など）
□分　【条件】質問 3 で『火曜日』の該当項目を選択した場合。入力値は 1～480。
室内で体を動かす運動（例：ゲーム機や動画以外。走り回る、親子で体を動かして遊ぶなど）
□分　【条件】質問 3 で『火曜日』の該当項目を選択した場合。入力値は 1～480。
屋内運動施設（例：体育館・プール、児童館、商業施設の遊び場など）
□分　【条件】質問 3 で『火曜日』の該当項目を選択した場合。入力値は 1～480。
その他の室内で体を動かす場面
□分　【条件】質問 3 で『火曜日』の該当項目を選択した場合。入力値は 1～480。

＜水曜日＞
室内でゲーム機や動画を見ながら体を動かす運動（例：Wii Sports、ニンテンドースイッチなどのゲーム、YouTube など）
□分　【条件】質問 3 で『水曜日』の該当項目を選択した場合。入力値は 1～480。
室内で体を動かす運動（例：ゲーム機や動画以外。走り回る、親子で体を動かして遊ぶなど）
□分　【条件】質問 3 で『水曜日』の該当項目を選択した場合。入力値は 1～480。
屋内運動施設（例：体育館・プール、児童館、商業施設の遊び場など）
□分　【条件】質問 3 で『水曜日』の該当項目を選択した場合。入力値は 1～480。
その他の室内で体を動かす場面
□分　【条件】質問 3 で『水曜日』の該当項目を選択した場合。入力値は 1～480。

＜木曜日＞
室内でゲーム機や動画を見ながら体を動かす運動（例：Wii Sports、ニンテンドースイッチなどのゲーム、YouTube など）
□分　【条件】質問 3 で『木曜日』の該当項目を選択した場合。入力値は 1～480。
室内で体を動かす運動（例：ゲーム機や動画以外。走り回る、親子で体を動かして遊ぶなど）
□分　【条件】質問 3 で『木曜日』の該当項目を選択した場合。入力値は 1～480。
屋内運動施設（例：体育館・プール、児童館、商業施設の遊び場など）
□分　【条件】質問 3 で『木曜日』の該当項目を選択した場合。入力値は 1～480。
その他の室内で体を動かす場面
□分　【条件】質問 3 で『木曜日』の該当項目を選択した場合。入力値は 1～480。

＜金曜日＞
室内でゲーム機や動画を見ながら体を動かす運動（例：Wii Sports、ニンテンドースイッチなどのゲーム、YouTube など）
□分　【条件】質問 3 で『金曜日』の該当項目を選択した場合。入力値は 1～480。
室内で体を動かす運動（例：ゲーム機や動画以外。走り回る、親子で体を動かして遊ぶなど）
□分　【条件】質問 3 で『金曜日』の該当項目を選択した場合。入力値は 1～480。
屋内運動施設（例：体育館・プール、児童館、商業施設の遊び場など）
□分　【条件】質問 3 で『金曜日』の該当項目を選択した場合。入力値は 1～480。
その他の室内で体を動かす場面
□分　【条件】質問 3 で『金曜日』の該当項目を選択した場合。入力値は 1～480。

＜土曜日＞
室内でゲーム機や動画を見ながら体を動かす運動（例：Wii Sports、ニンテンドースイッチなどのゲーム、YouTube など）
[　　]分　【条件】質問 3 で『土曜日』の該当項目を選択した場合。入力値は 1～480。
室内で体を動かす運動（例：ゲーム機や動画以外。走り回る、親子で体を動かして遊ぶなど）
[　　]分　【条件】質問 3 で『土曜日』の該当項目を選択した場合。入力値は 1～480。
屋内運動施設（例：体育館・プール、児童館、商業施設の遊び場など）
[　　]分　【条件】質問 3 で『土曜日』の該当項目を選択した場合。入力値は 1～480。
その他の室内で体を動かす場面
[　　]分　【条件】質問 3 で『土曜日』の該当項目を選択した場合。入力値は 1～480。

＜日曜日＞
室内でゲーム機や動画を見ながら体を動かす運動（例：Wii Sports、ニンテンドースイッチなどのゲーム、YouTube など）
[　　]分　【条件】質問 3 で『日曜日』の該当項目を選択した場合。入力値は 1～480。
室内で体を動かす運動（例：ゲーム機や動画以外。走り回る、親子で体を動かして遊ぶなど）
[　　]分　【条件】質問 3 で『日曜日』の該当項目を選択した場合。入力値は 1～480。
屋内運動施設（例：体育館・プール、児童館、商業施設の遊び場など）
[　　]分　【条件】質問 3 で『日曜日』の該当項目を選択した場合。入力値は 1～480。
その他の室内で体を動かす場面
[　　]分　【条件】質問 3 で『日曜日』の該当項目を選択した場合。入力値は 1～480。

**質問 3_2.** これらの室内や屋内施設で体を動かす遊びのうち、息がはずむ程度のきつさの運動時間は何分くらいありますか？**ない場合は「0 分」と入力してください。**
※0～480 の範囲でお答えください。※半角でご入力ください。

月曜日[　　]分　【条件】質問 3 で『月曜日』の遊び項目を選択した場合。入力値は 0～480。

火曜日[　　]分　【条件】質問 3 で『火曜日』の遊び項目を選択した場合。入力値は 0～480。

水曜日[　　]分　【条件】質問 3 で『水曜日』の遊び項目を選択した場合。入力値は 0～480。

木曜日[　　]分　【条件】質問 3 で『木曜日』の遊び項目を選択した場合。入力値は 0～480。

金曜日[　　]分　【条件】質問 3 で『金曜日』の遊び項目を選択した場合。入力値は 0～480。

土曜日[　　]分　【条件】質問 3 で『土曜日』の遊び項目を選択した場合。入力値は 0～480。

日曜日[　　]分　【条件】質問 3 で『日曜日』の遊び項目を選択した場合。入力値は 0～480。

#### お子様の生活習慣についてお答えください

**質問 4.** お子様の平日（月～金）と休日（土・日）の就寝時刻と起床時刻を 24 時間表記で教えてください。（例：午後 10 時⇒22 時）

＜平日＞就寝時刻
[　　]時 [　　]分　【条件】時（18 時から 1 時間刻み）。分（5 分刻み）。選択式。

＜平日＞起床時刻
[　　]時 [　　]分　【条件】時（5 時から 1 時間刻み）。分（5 分刻み）。選択式。

＜休日＞就寝時刻
[　　]時 [　　]分　【条件】時（18 時から 1 時間刻み）。分（5 分刻み）。選択式。

＜休日＞起床時刻

[ ]時 [ ]分　【条件】時（5 時から 1 時間刻み）。分（5 分刻み）。選択式。

**質問 5.** ここ 1 ヶ月、お子様は、園（幼稚園・保育園など含む）の活動以外で、テレビや DVD をみたり、パソコン、ゲーム（テレビ、パソコン、携帯式のゲーム機などを含む）、スマートフォンなどを使用したりする時間は 1 日あたりどのくらいですか。（ただし、Wii Sports などの体を動かすゲーム、YouTube などの動画を見ながら体を動かす時間は含めません）

＜平日（月～金）＞

○ 30 分未満
○ 30 分～1 時間未満
○ 1～2 時間未満
○ 2～3 時間未満
○ 3～4 時間未満
○ 4～5 時間未満
○ 5 時間以上

＜休日（土・日）＞

○ 30 分未満
○ 30 分～1 時間未満
○ 1～2 時間未満
○ 2～3 時間未満
○ 3～4 時間未満
○ 4～5 時間未満
○ 5 時間以上

[回答を提出する]

**eMaterial 4. SSF-PAQ (Original version) — electronic, English translation**

*Note: This English version was translated by the authors to help readers understand the original Japanese questionnaire. It has not undergone a formal translation and cross-cultural adaptation process.*

#### Questionnaire on Physical Activity, Sleep, and Screen Time in Preschool Children
#### SSF Preschoolers' Activity Questionnaire (SSF-PAQ)

**Q1.** Over the past month, has your child taken any organized sports or exercise lessons? (Do not include time spent at preschool: kindergarten, nursery school, or center for early childhood education and care. However, do include organized sports or exercise lessons that your family voluntarily arranges after preschool hours using the preschool's facilities or grounds.) (Select one)

○ Yes
○ No [If "No" is selected, skip to Question 2]

**Q1_1.** On which days of the week does your child take organized sports or exercise lessons? (Select all that apply)

☐ Monday ☐ Tuesday ☐ Wednesday ☐ Thursday ☐ Friday ☐ Saturday ☐ Sunday

**Q1_2.** Thinking back over the past month, please enter your child's average daily time (minutes) spent in organized sports or exercise lessons for each day of the week.

*Enter a value between 1 and 480. *Enter numbers as half-width (single-byte) digits.

Monday ______ min [Shown if "Monday" was selected in Q1_1; range: 1–480]
Tuesday ______ min [Shown if "Tuesday" was selected in Q1_1; range: 1–480]
Wednesday ______ min [Shown if "Wednesday" was selected in Q1_1; range: 1–480]
Thursday ______ min [Shown if "Thursday" was selected in Q1_1; range: 1–480]
Friday ______ min [Shown if "Friday" was selected in Q1_1; range: 1–480]
Saturday ______ min [Shown if "Saturday" was selected in Q1_1; range: 1–480]
Sunday ______ min [Shown if "Sunday" was selected in Q1_1; range: 1–480]

**Q1_3.** Of these lessons, about how many minutes per day were spent in activity that makes your child breathe harder? **Enter "0" if none.**

*Enter a value between 0 and 480. *Enter numbers as half-width (single-byte) digits.

Monday ______ min [Shown if "Monday" was selected in Q1_1; range: 0–480]
Tuesday ______ min [Shown if "Tuesday" was selected in Q1_1; range: 0–480]
Wednesday ______ min [Shown if "Wednesday" was selected in Q1_1; range: 0–480]
Thursday ______ min [Shown if "Thursday" was selected in Q1_1; range: 0–480]
Friday ______ min [Shown if "Friday" was selected in Q1_1; range: 0–480]
Saturday ______ min [Shown if "Saturday" was selected in Q1_1; range: 0–480]
Sunday ______ min [Shown if "Sunday" was selected in Q1_1; range: 0–480]

**The following questions ask about your child's active play time outside of preschool and organized sports or exercise lessons.**

**Q2.** Outside of preschool and organized sports or exercise lessons, does your child engage in outdoor active play? Thinking back over the past month, select all the locations where your child plays, for each day of the week. **For days with no outdoor active play, select “No outdoor active play on this day.”**

**<Monday>**

- ☐ Parks or outdoor playgrounds
- ☐ Home premises (yard, etc.)
- ☐ Streets (roads, alleys, etc.)
- ☐ Other outdoor play locations
- ☐ No outdoor active play on this day

**<Tuesday>**

- ☐ Parks or outdoor playgrounds
- ☐ Home premises (yard, etc.)
- ☐ Streets (roads, alleys, etc.)
- ☐ Other outdoor play locations
- ☐ No outdoor active play on this day

**<Wednesday>**

- ☐ Parks or outdoor playgrounds
- ☐ Home premises (yard, etc.)
- ☐ Streets (roads, alleys, etc.)
- ☐ Other outdoor play locations
- ☐ No outdoor active play on this day

**<Thursday>**

- ☐ Parks or outdoor playgrounds
- ☐ Home premises (yard, etc.)
- ☐ Streets (roads, alleys, etc.)
- ☐ Other outdoor play locations
- ☐ No outdoor active play on this day

**<Friday>**

- ☐ Parks or outdoor playgrounds
- ☐ Home premises (yard, etc.)
- ☐ Streets (roads, alleys, etc.)
- ☐ Other outdoor play locations
- ☐ No outdoor active play on this day

**<Saturday>**

- ☐ Parks or outdoor playgrounds
- ☐ Home premises (yard, etc.)
- ☐ Streets (roads, alleys, etc.)
- ☐ Other outdoor play locations
- ☐ No outdoor active play on this day

**<Sunday>**

- ☐ Parks or outdoor playgrounds
- ☐ Home premises (yard, etc.)
- ☐ Streets (roads, alleys, etc.)
- ☐ Other outdoor play locations
- ☐ No outdoor active play on this day

**Q2_1.** Outside of preschool and organized sports or exercise lessons, how much time does your child spend in outdoor active play? Thinking back over the past month, please enter the average daily time (minutes) for each play location on each day of the week.

Example: If your child plays at a park for an hour and a half on Sundays, enter “90” in the “Parks or outdoor playgrounds” field under Sunday.

*Enter a value between 1 and 480. *Enter numbers as half-width (single-byte) digits.

**<Monday>**

Parks or outdoor playgrounds
_______ min [Shown if this location was selected for Monday in Q2; range: 1–480]

Home premises (yard, etc.)
_______ min [Shown if this location was selected for Monday in Q2; range: 1–480]

Streets (roads, alleys, etc.)
_______ min [Shown if this location was selected for Monday in Q2; range: 1–480]

Other outdoor play locations
_______ min [Shown if this location was selected for Monday in Q2; range: 1–480]

**<Tuesday>**

Parks or outdoor playgrounds
_______ min [Shown if this location was selected for Tuesday in Q2; range: 1–480]

Home premises (yard, etc.)
_______ min [Shown if this location was selected for Tuesday in Q2; range: 1–480]

Streets (roads, alleys, etc.)
_______ min [Shown if this location was selected for Tuesday in Q2; range: 1–480]

Other outdoor play locations
_______ min [Shown if this location was selected for Tuesday in Q2; range: 1–480]

**<Wednesday>**

Parks or outdoor playgrounds
_______ min [Shown if this location was selected for Wednesday in Q2; range: 1–480]

Home premises (yard, etc.)
_______ min [Shown if this location was selected for Wednesday in Q2; range: 1–480]

Streets (roads, alleys, etc.)
_______ min [Shown if this location was selected for Wednesday in Q2; range: 1–480]

Other outdoor play locations
_______ min [Shown if this location was selected for Wednesday in Q2; range: 1–480]

**<Thursday>**

Parks or outdoor playgrounds
_______ min [Shown if this location was selected for Thursday in Q2; range: 1–480]

Home premises (yard, etc.)
_______ min [Shown if this location was selected for Thursday in Q2; range: 1–480]

Streets (roads, alleys, etc.)
_______ min [Shown if this location was selected for Thursday in Q2; range: 1–480]

Other outdoor play locations
_______ min [Shown if this location was selected for Thursday in Q2; range: 1–480]

**<Friday>**

Parks or outdoor playgrounds
_______ min [Shown if this location was selected for Friday in Q2; range: 1–480]

Home premises (yard, etc.)
_______ min [Shown if this location was selected for Friday in Q2; range: 1–480]

Streets (roads, alleys, etc.)
_______ min [Shown if this location was selected for Friday in Q2; range: 1–480]

Other outdoor play locations
_______ min [Shown if this location was selected for Friday in Q2; range: 1–480]

**<Saturday>**

Parks or outdoor playgrounds
_______ min [Shown if this location was selected for Saturday in Q2; range: 1–480]

Home premises (yard, etc.)
_______ min [Shown if this location was selected for Saturday in Q2; range: 1–480]

Streets (roads, alleys, etc.)
_______ min [Shown if this location was selected for Saturday in Q2; range: 1–480]

Other outdoor play locations
_______ min [Shown if this location was selected for Saturday in Q2; range: 1–480]

**<Sunday>**

Parks or outdoor playgrounds
_______ min [Shown if this location was selected for Sunday in Q2; range: 1–480]

Home premises (yard, etc.)
_______ min [Shown if this location was selected for Sunday in Q2; range: 1–480]

Streets (roads, alleys, etc.)
_______ min [Shown if this location was selected for Sunday in Q2; range: 1–480]

Other outdoor play locations
_______ min [Shown if this location was selected for Sunday in Q2; range: 1–480]

**Q2_2.** Of this outdoor play, about how many minutes per day were spent in activity that makes your child breathe harder? **Enter "0" if none.**

*Enter a value between 0 and 480. *Enter numbers as half-width (single-byte) digits.

Monday _______ min [Shown if any location was selected for Monday in Q2; range: 0–480]

Tuesday _______ min [Shown if any location was selected for Tuesday in Q2; range: 0–480]

Wednesday _______ min [Shown if any location was selected for Wednesday in Q2; range: 0–480]

Thursday _______ min [Shown if any location was selected for Thursday in Q2; range: 0–480]

Friday _______ min [Shown if any location was selected for Friday in Q2; range: 0–480]

Saturday _______ min [Shown if any location was selected for Saturday in Q2; range: 0–480]

Sunday _______ min [Shown if any location was selected for Sunday in Q2; range: 0–480]

**Q3.** Outside of preschool and organized sports or exercise lessons, does your child engage in active play indoors or at indoor facilities? Thinking back over the past month, select all applicable items for each day of the week. **For days with no indoor active play, select "No indoor active play on this day."**

**<Monday>**

- ☐ Active play indoors with video games or videos (e.g., Wii Sports, Nintendo Switch games, YouTube)
- ☐ Active play indoors without games or videos (e.g., running around, active play with parents)
- ☐ Indoor exercise facilities (e.g., gymnasium, swimming pool, children's center, indoor playgrounds in commercial facilities)
- ☐ Other indoor active play situations
- ☐ No indoor active play on this day

eMaterial 4. Continued.

**<Tuesday>**

- ☐ Active play indoors with video games or videos (e.g., Wii Sports, Nintendo Switch games, YouTube)
- ☐ Active play indoors without games or videos (e.g., running around, active play with parents)
- ☐ Indoor exercise facilities (e.g., gymnasium, swimming pool, children's center, indoor playgrounds in commercial facilities)
- ☐ Other indoor active play situations
- ☐ No indoor active play on this day

**<Wednesday>**

- ☐ Active play indoors with video games or videos (e.g., Wii Sports, Nintendo Switch games, YouTube)
- ☐ Active play indoors without games or videos (e.g., running around, active play with parents)
- ☐ Indoor exercise facilities (e.g., gymnasium, swimming pool, children's center, indoor playgrounds in commercial facilities)
- ☐ Other indoor active play situations
- ☐ No indoor active play on this day

**<Thursday>**

- ☐ Active play indoors with video games or videos (e.g., Wii Sports, Nintendo Switch games, YouTube)
- ☐ Active play indoors without games or videos (e.g., running around, active play with parents)
- ☐ Indoor exercise facilities (e.g., gymnasium, swimming pool, children's center, indoor playgrounds in commercial facilities)
- ☐ Other indoor active play situations
- ☐ No indoor active play on this day

**<Friday>**

- ☐ Active play indoors with video games or videos (e.g., Wii Sports, Nintendo Switch games, YouTube)
- ☐ Active play indoors without games or videos (e.g., running around, active play with parents)
- ☐ Indoor exercise facilities (e.g., gymnasium, swimming pool, children's center, indoor playgrounds in commercial facilities)
- ☐ Other indoor active play situations
- ☐ No indoor active play on this day

**<Saturday>**

- ☐ Active play indoors with video games or videos (e.g., Wii Sports, Nintendo Switch games, YouTube)
- ☐ Active play indoors without games or videos (e.g., running around, active play with parents)
- ☐ Indoor exercise facilities (e.g., gymnasium, swimming pool, children's center, indoor playgrounds in commercial facilities)
- ☐ Other indoor active play situations
- ☐ No indoor active play on this day

**<Sunday>**

- ☐ Active play indoors with video games or videos (e.g., Wii Sports, Nintendo Switch games, YouTube)
- ☐ Active play indoors without games or videos (e.g., running around, active play with parents)
- ☐ Indoor exercise facilities (e.g., gymnasium, swimming pool, children's center, indoor playgrounds in commercial facilities)
- ☐ Other indoor active play situations
- ☐ No indoor active play on this day

**Q3_1.** Outside of preschool and organized sports or exercise lessons, how much time does your child spend in active play indoors or at indoor facilities? Thinking back over the past month, please enter the average daily time (minutes) for each item on each day of the week.

*Enter a value between 1 and 480. *Enter numbers as half-width (single-byte) digits.

**<Monday>**

Active play indoors with video games or videos (e.g., Wii Sports, Nintendo Switch games, YouTube)

_______ min [Shown if this item was selected for Monday in Q3; range: 1–480]

Active play indoors without games or videos (e.g., running around, active play with parents)

_______ min [Shown if this item was selected for Monday in Q3; range: 1–480]

Indoor exercise facilities (e.g., gymnasium, swimming pool, children's center, indoor playgrounds in commercial facilities)

_______ min [Shown if this item was selected for Monday in Q3; range: 1–480]

Other indoor active play situations

_______ min [Shown if this item was selected for Monday in Q3; range: 1–480]

**<Tuesday>**

Active play indoors with video games or videos (e.g., Wii Sports, Nintendo Switch games, YouTube)

_______ min [Shown if this item was selected for Tuesday in Q3; range: 1–480]

Active play indoors without games or videos (e.g., running around, active play with parents)

_______ min [Shown if this item was selected for Tuesday in Q3; range: 1–480]

Indoor exercise facilities (e.g., gymnasium, swimming pool, children's center, indoor playgrounds in commercial facilities)

_______ min [Shown if this item was selected for Tuesday in Q3; range: 1–480]

Other indoor active play situations

_______ min [Shown if this item was selected for Tuesday in Q3; range: 1–480]

**<Wednesday>**

Active play indoors with video games or videos (e.g., Wii Sports, Nintendo Switch games, YouTube)

_______ min [Shown if this item was selected for Wednesday in Q3; range: 1–480]

Active play indoors without games or videos (e.g., running around, active play with parents)

_______ min [Shown if this item was selected for Wednesday in Q3; range: 1–480]

Indoor exercise facilities (e.g., gymnasium, swimming pool, children's center, indoor playgrounds in commercial facilities)

_______ min [Shown if this item was selected for Wednesday in Q3; range: 1–480]

Other indoor active play situations

_______ min [Shown if this item was selected for Wednesday in Q3; range: 1–480]

**<Thursday>**

Active play indoors with video games or videos (e.g., Wii Sports, Nintendo Switch games, YouTube)

_______ min [Shown if this item was selected for Thursday in Q3; range: 1–480]

Active play indoors without games or videos (e.g., running around, active play with parents)

_______ min [Shown if this item was selected for Thursday in Q3; range: 1–480]

Indoor exercise facilities (e.g., gymnasium, swimming pool, children's center, indoor playgrounds in commercial facilities)

_______ min [Shown if this item was selected for Thursday in Q3; range: 1–480]

Other indoor active play situations

_______ min [Shown if this item was selected for Thursday in Q3; range: 1–480]

**<Friday>**

Active play indoors with video games or videos (e.g., Wii Sports, Nintendo Switch games, YouTube)

_______ min [Shown if this item was selected for Friday in Q3; range: 1–480]

Active play indoors without games or videos (e.g., running around, active play with parents)

_______ min [Shown if this item was selected for Friday in Q3; range: 1–480]

Indoor exercise facilities (e.g., gymnasium, swimming pool, children's center, indoor playgrounds in commercial facilities)

_______ min [Shown if this item was selected for Friday in Q3; range: 1–480]

Other indoor active play situations

_______ min [Shown if this item was selected for Friday in Q3; range: 1–480]

**<Saturday>**

Active play indoors with video games or videos (e.g., Wii Sports, Nintendo Switch games, YouTube)

_______ min [Shown if this item was selected for Saturday in Q3; range: 1–480]

Active play indoors without games or videos (e.g., running around, active play with parents)

_______ min [Shown if this item was selected for Saturday in Q3; range: 1–480]

Indoor exercise facilities (e.g., gymnasium, swimming pool, children's center, indoor playgrounds in commercial facilities)

_______ min [Shown if this item was selected for Saturday in Q3; range: 1–480]

Other indoor active play situations

_______ min [Shown if this item was selected for Saturday in Q3; range: 1–480]

**<Sunday>**

Active play indoors with video games or videos (e.g., Wii Sports, Nintendo Switch games, YouTube)

_______ min [Shown if this item was selected for Sunday in Q3; range: 1–480]

Active play indoors without games or videos (e.g., running around, active play with parents)

_______ min [Shown if this item was selected for Sunday in Q3; range: 1–480]

Indoor exercise facilities (e.g., gymnasium, swimming pool, children's center, indoor playgrounds in commercial facilities)

_______ min [Shown if this item was selected for Sunday in Q3; range: 1–480]

Other indoor active play situations

_______ min [Shown if this item was selected for Sunday in Q3; range: 1–480]

**Q3_2.** Of this indoor play, about how many minutes per day were spent in activity that makes your child breathe harder? **Enter "0" if none.**

*Enter a value between 0 and 480. *Enter numbers as half-width (single-byte) digits.

Monday _______ min [Shown if any item was selected for Monday in Q3; range: 0–480]

Tuesday _______ min [Shown if any item was selected for Tuesday in Q3; range: 0–480]

Wednesday _______ min [Shown if any item was selected for Wednesday in Q3; range: 0–480]

Thursday _______ min [Shown if any item was selected for Thursday in Q3; range: 0–480]

Friday _______ min [Shown if any item was selected for Friday in Q3; range: 0–480]

Saturday _______ min [Shown if any item was selected for Saturday in Q3; range: 0–480]

Sunday _______ min [Shown if any item was selected for Sunday in Q3; range: 0–480]

**Please answer the following questions about your child's daily routine.**

**Q4.** Please indicate your child's bedtime and wake-up time on weekdays (Mon–Fri) and weekend days (Sat–Sun), using the 24-hour clock (e.g., 10 p.m. → 22:00).

**<Weekdays> Bedtime**

_______ h _______ min [Drop-down: hour from 18:00 in 1-hour steps; minutes in 5-minute steps]

**<Weekdays> Wake-up time**

_______ h _______ min [Drop-down: hour from 5:00 in 1-hour steps; minutes in 5-minute steps]

**<Weekend days> Bedtime**

_______ h _______ min [Drop-down: hour from 18:00 in 1-hour steps; minutes in 5-minute steps]

**<Weekend days> Wake-up time**

_______ h _______ min [Drop-down: hour from 5:00 in 1-hour steps; minutes in 5-minute steps]

eMaterial 4. Continued.

**Q5.** Over the past month, outside of preschool activities, about how much time per day has your child spent watching TV or DVDs, or using computers, video games (including TV, computer, and handheld game consoles), smartphones, and similar devices? (Do not include active video games such as Wii Sports or time spent being physically active while watching videos such as YouTube.)

**<Weekdays (Mon–Fri)>**

- ○ Less than 30 min
- ○ 30 min to <1 h
- ○ 1 to <2 h
- ○ 2 to <3 h
- ○ 3 to <4 h
- ○ 4 to <5 h
- ○ 5 h or more

**<Weekend days (Sat–Sun)>**

- ○ Less than 30 min
- ○ 30 min to <1 h
- ○ 1 to <2 h
- ○ 2 to <3 h
- ○ 3 to <4 h
- ○ 4 to <5 h
- ○ 5 h or more

**[Submit responses]**

### 幼児の身体活動、睡眠、スクリーンタイムに関する質問票（24 時間版）

#### SSF Preschoolers' Activity Questionnaire-24h (SSF-PAQ-24)

**質問 1．** ここ 1 ヶ月について、お子様は運動の習いごとをしていますか？（幼稚園や保育園などにいる時間は含めません。ただし、園が終了してから、園の施設や場所を使って家庭が任意で習わせる運動の時間は含みます）（ひとつだけ）

[1] はい　　[2] いいえ　→ **質問 2 へ**

**質問 1_1.** お子様の運動の習いごとについて、ここ 1 ヶ月を思い出し、曜日ごとに平均的な 1 日あたりの時間（分）を回答してください。**運動の習いごとをしていない曜日は「0 分」と記入してください。**

| | 月曜日 | 火曜日 | 水曜日 | 木曜日 | 金曜日 | 土曜日 | 日曜日 |
|---|---|---|---|---|---|---|---|
| 運動の習いごとをしている時間 | 分 | 分 | 分 | 分 | 分 | 分 | 分 |

**質問 1_3.** これらの運動の習いごとのうち、息がはずむ程度のきつさの運動時間は何分くらいありますか？**ない場合は「0 分」と記入してください。**

| | 月曜日 | 火曜日 | 水曜日 | 木曜日 | 金曜日 | 土曜日 | 日曜日 |
|---|---|---|---|---|---|---|---|
| 息がはずむ程度のきつさの運動時間 | 分 | 分 | 分 | 分 | 分 | 分 | 分 |

**ここからは、お子様が運動の習いごと以外で過ごす「体を動かして遊ぶ時間」についての質問です。**

**質問 2.**運動の習いごと以外で、お子様はどれくらいの時間、体を動かす外遊びをしていますか？ここ 1 ヶ月を思い出し、各曜日の遊んでいる場所ごとに、平均的な 1 日あたりの時間（分）を回答してください。幼稚園など園にいる時間については、あなたが把握している範囲で「園の屋外（園庭やグラウンドなど）」欄に回答してください。
例：日曜日に公園で 1 時間半遊んでいる場合、日曜日の「公園や屋外の遊び場」の欄に「90」と入力してください。**外で体を動かしていない曜日は「0 分」と記入してください。**

| | 月曜日 | 火曜日 | 水曜日 | 木曜日 | 金曜日 | 土曜日 | 日曜日 |
|---|---|---|---|---|---|---|---|
| 園の屋外（園庭やグラウンドなど） | 分 | 分 | 分 | 分 | 分 | 分 | 分 |
| 公園や屋外の遊び場 | 分 | 分 | 分 | 分 | 分 | 分 | 分 |
| 自宅の敷地内（庭など） | 分 | 分 | 分 | 分 | 分 | 分 | 分 |
| 路上（道路や路地裏など） | 分 | 分 | 分 | 分 | 分 | 分 | 分 |
| その他の外遊びの場所（　　　　　　） | 分 | 分 | 分 | 分 | 分 | 分 | 分 |

**質問 2_1.** これらの外遊びのうち、息がはずむ程度のきつさの運動時間は何分くらいありますか？**ない場合は「0 分」と記入してください。**

| | 月曜日 | 火曜日 | 水曜日 | 木曜日 | 金曜日 | 土曜日 | 日曜日 |
|---|---|---|---|---|---|---|---|
| 息がはずむ程度のきつさの運動時間 | 分 | 分 | 分 | 分 | 分 | 分 | 分 |

**質問 3.** 運動の習いごと以外で、お子様はどれくらいの時間、室内や屋内施設で体を動かす遊びをしていますか？ここ 1 ヶ月を思い出し、曜日ごとに平均的な 1 日あたりの時間（分）を回答してください。幼稚園など園にいる時間については、あなたが把握している範囲で「園の屋内での運動遊び（ホールや遊戯室など）」欄に回答してください。
**室内で体を動かして遊ばない曜日は「0 分」と記入してください。**

| | 月曜日 | 火曜日 | 水曜日 | 木曜日 | 金曜日 | 土曜日 | 日曜日 |
|---|---|---|---|---|---|---|---|
| 園の屋内での運動遊び（ホールや遊戯室など） | 分 | 分 | 分 | 分 | 分 | 分 | 分 |
| 室内でゲーム機や動画を 見ながら体を動かす運動（Wii Sports、ニンテンドースイッチなどのゲーム、YouTube など） | 分 | 分 | 分 | 分 | 分 | 分 | 分 |
| 室内で体を動かす運動（ゲーム機や動画以外。走り回る、親子で体を動かして遊ぶなど） | 分 | 分 | 分 | 分 | 分 | 分 | 分 |
| 屋内運動施設（例 体育館・プール、児童館、商業施設の遊び場など） | 分 | 分 | 分 | 分 | 分 | 分 | 分 |
| その他の室内で体を動かす場面（　　　　　　　） | 分 | 分 | 分 | 分 | 分 | 分 | 分 |

**質問 3_1.** これらの室内や屋内施設で体を動かす遊びのうち、息がはずむ程度のきつさの運動時間は何分くらいありますか？**ない場合は「0 分」と記入してください。**

| | 月曜日 | 火曜日 | 水曜日 | 木曜日 | 金曜日 | 土曜日 | 日曜日 |
|---|---|---|---|---|---|---|---|
| 息がはずむ程度のきつさの運動時間 | 分 | 分 | 分 | 分 | 分 | 分 | 分 |

#### お子様の生活習慣についてお答えください

**質問 4.** お子様の平日（月～金）と休日（土・日）の就寝時刻と起床時刻を 24 時間表記で教えてください。（例：午後 10 時⇒22 時）

| | 就寝時刻 | | 起床時刻 | |
|---|---|---|---|---|
| 平日 | 時 | 分 | 時 | 分 |
| 休日 | 時 | 分 | 時 | 分 |

**質問 5.** ここ 1 ヶ月、お子様は、テレビや DVD をみたり、パソコン、ゲーム（テレビ、パソコン、携帯式のゲーム機などを含む）、スマートフォンなどを使用したりする時間は 1 日あたりどのくらいですか。（ただし、Wii Sports などの体を動かすゲーム、YouTube などの動画を見ながら体を動かす時間は含めません）

| | 30 分未満 | 30 分～1 時間未満 | 1～2 時間未満 | 2～3 時間未満 | 3～4 時間未満 | 4～5 時間未満 | 5 時間以上 |
|---|---|---|---|---|---|---|---|
| 平日（月～金） | [1] | [2] | [3] | [4] | [5] | [6] | [7] |
| 休日（土・日） | [1] | [2] | [3] | [4] | [5] | [6] | [7] |

ご回答、ありがとうございました。

**eMaterial 6. SSF-PAQ-24 (24-hour version) — paper, English translation**

*Note: This English version was translated by the authors to help readers understand the original Japanese questionnaire. It has not undergone a formal translation and cross-cultural adaptation process.*

**Questionnaire on Physical Activity, Sleep, and Screen Time in Preschool Children (24-Hour Version)**

#### SSF Preschoolers' Activity Questionnaire-24h (SSF-PAQ-24)

**Q1.** Over the past month, has your child taken any organized sports or exercise lessons? (Do not include time spent at preschool: kindergarten, nursery school, or center for early childhood education and care. However, do include organized sports or exercise lessons that your family voluntarily arranges after preschool hours using the preschool's facilities or grounds.) (Select one)

[1] Yes [2] No → **Go to Question 2**

**Q1_1.** Thinking back over the past month, please enter your child's average daily time (minutes) spent in organized sports or exercise lessons for each day of the week. **Enter "0" for days with no lessons.**

| | Mon | Tue | Wed | Thu | Fri | Sat | Sun |
|---|---|---|---|---|---|---|---|
| Time spent in organized sports or exercise lessons | min | min | min | min | min | min | min |

**Q1_3.** Of these lessons, about how many minutes per day were spent in activity that makes your child breathe harder? **Enter "0" if none.**

| | Mon | Tue | Wed | Thu | Fri | Sat | Sun |
|---|---|---|---|---|---|---|---|
| Time in activity that makes your child breathe harder | min | min | min | min | min | min | min |

**The following questions ask about your child's active play time outside of organized sports or exercise lessons.**

**Q2.** Outside of organized sports or exercise lessons, how much time does your child spend in outdoor active play? Thinking back over the past month, please enter the average daily time (minutes) for each play location on each day of the week. For time spent at preschool, answer in the "Preschool outdoor areas (playground, sports ground, etc.)" row to the best of your knowledge.

Example: If your child plays at a park for an hour and a half on Sundays, enter "90" in the "Parks or outdoor playgrounds" row for Sunday. **Enter "0" for days with no outdoor active play.**

| | Mon | Tue | Wed | Thu | Fri | Sat | Sun |
|---|---|---|---|---|---|---|---|
| Preschool outdoor areas (playground, sports ground, etc.) | min | min | min | min | min | min | min |
| Parks or outdoor playgrounds | min | min | min | min | min | min | min |
| Home premises (yard, etc.) | min | min | min | min | min | min | min |
| Streets (roads, alleys, etc.) | min | min | min | min | min | min | min |
| Other outdoor play locations (________) | min | min | min | min | min | min | min |

**Q2_1.** Of this outdoor play, about how many minutes per day were spent in activity that makes your child breathe harder? **Enter "0" if none.**

| | Mon | Tue | Wed | Thu | Fri | Sat | Sun |
|---|---|---|---|---|---|---|---|
| Time in activity that makes your child breathe harder | min | min | min | min | min | min | min |

**Q3.** Outside of organized sports or exercise lessons, how much time does your child spend in active play indoors or at indoor facilities? Thinking back over the past month, please enter the average daily time (minutes) for each day of the week. For time spent at preschool, answer in the “Indoor active play at preschool (hall, playroom, etc.)” row to the best of your knowledge. **Enter “0” for days with no indoor active play.**

| | Mon | Tue | Wed | Thu | Fri | Sat | Sun |
|---|---|---|---|---|---|---|---|
| Indoor active play at preschool (hall, playroom, etc.) | min | min | min | min | min | min | min |
| Active play indoors with video games or videos (e.g., Wii Sports, Nintendo Switch games, YouTube) | min | min | min | min | min | min | min |
| Active play indoors without games or videos (e.g., running around, active play with parents) | min | min | min | min | min | min | min |
| Indoor exercise facilities (e.g., gymnasium, swimming pool, children’s center, indoor playgrounds in commercial facilities) | min | min | min | min | min | min | min |
| Other indoor active play situations (________) | min | min | min | min | min | min | min |

**Q3_1.** Of this indoor play, about how many minutes per day were spent in activity that makes your child breathe harder? **Enter “0” if none.**

| | Mon | Tue | Wed | Thu | Fri | Sat | Sun |
|---|---|---|---|---|---|---|---|
| Time in activity that makes your child breathe harder | min | min | min | min | min | min | min |

**Please answer the following questions about your child’s daily routine.**

**Q4.** Please indicate your child’s bedtime and wake-up time on weekdays (Mon–Fri) and weekend days (Sat–Sun), using the 24-hour clock (e.g., 10 p.m. → 22:00).

| | Bedtime | | Wake-up time | |
|---|---|---|---|---|
| Weekdays | h | min | h | min |
| Weekend days | h | min | h | min |

**Q5.** Over the past month, about how much time per day has your child spent watching TV or DVDs, or using computers, video games (including TV, computer, and handheld game consoles), smartphones, and similar devices? (Do not include active video games such as Wii Sports or time spent being physically active while watching videos such as YouTube.)

| | Less than 30 min | 30 min to <1 h | 1 to <2 h | 2 to <3 h | 3 to <4 h | 4 to <5 h | 5 h or more |
|---|---|---|---|---|---|---|---|
| Weekdays (Mon–Fri) | [1] | [2] | [3] | [4] | [5] | [6] | [7] |
| Weekend days (Sat–Sun) | [1] | [2] | [3] | [4] | [5] | [6] | [7] |

**Thank you for completing this questionnaire.**

**eMaterial 7. SSF-PAQ-24 (24-hour version) — electronic, Japanese**

幼児の身体活動、睡眠、スクリーンタイムに関する質問票（24 時間版）

**SSF Preschoolers' Activity Questionnaire-24h (SSF-PAQ-24)**

**質問 1．** ここ 1 ヶ月について、お子様は運動の習いごとをしていますか？（幼稚園や保育園などにいる時間は含めません。ただし、園が終了してから、園の施設や場所を使って家庭が任意で習わせる運動の時間は含みます）（ひとつだけ）

○ はい
○ いいえ 【条件】『いいえ』を選択した場合は質問 2 へ

**質問 1_1.** お子様が運動の習いごとをしている曜日をすべてお答えください。（あてはまるものすべて）

☐月曜日 ☐火曜日 ☐水曜日 ☐木曜日 ☐金曜日 ☐土曜日 ☐日曜日

**質問 1_2.** お子様の運動の習いごとについて、ここ 1 ヶ月を思い出し、曜日ごとに平均的な 1 日あたりの時間（分）を回答してください。
※1～480 の範囲でお答えください。※半角でご入力ください。

月曜日[ ]分 【条件】質問 1_1 で『月曜日』を選択した場合。入力値は 1～480。

火曜日[ ]分 【条件】質問 1_1 で『火曜日』を選択した場合。入力値は 1～480。

水曜日[ ]分 【条件】質問 1_1 で『水曜日』を選択した場合。入力値は 1～480。

木曜日[ ]分 【条件】質問 1_1 で『木曜日』を選択した場合。入力値は 1～480。

金曜日[ ]分 【条件】質問 1_1 で『金曜日』を選択した場合。入力値は 1～480。

土曜日[ ]分 【条件】質問 1_1 で『土曜日』を選択した場合。入力値は 1～480。

日曜日[ ]分 【条件】質問 1_1 で『日曜日』を選択した場合。入力値は 1～480。

**質問 1_3.** これらの運動の習いごとのうち、息がはずむ程度のきつさの運動時間は何分くらいありますか？ **ない場合は「0 分」と入力してください。**
※0～480 の範囲でお答えください。※半角でご入力ください。

月曜日[ ]分 【条件】質問 1_1 で『月曜日』を選択した場合。入力値は 0～480。

火曜日[ ]分 【条件】質問 1_1 で『火曜日』を選択した場合。入力値は 0～480。

水曜日[ ]分 【条件】質問 1_1 で『水曜日』を選択した場合。入力値は 0～480。

木曜日[ ]分 【条件】質問 1_1 で『木曜日』を選択した場合。入力値は 0～480。

金曜日[ ]分 【条件】質問 1_1 で『金曜日』を選択した場合。入力値は 0～480。

土曜日[ ]分 【条件】質問 1_1 で『土曜日』を選択した場合。入力値は 0～480。

日曜日[ ]分 【条件】質問 1_1 で『日曜日』を選択した場合。入力値は 0～480。

**ここからは、お子様が運動の習いごと以外で過ごす「体を動かして遊ぶ時間」についての質問です。**

**質問 2.** 運動の習いごと以外で、お子様は体を動かす外遊びをしていますか？ここ 1 ヶ月を思い出し、曜日ごとに遊んでいる場所をすべて選択してください。幼稚園など園にいる時間については、あなたが把握している範囲で「園の屋外（園庭やグラウンドなど）」を選択してください。**体を動かす外遊びをしない曜日は「この曜日は体を動かす外遊びをしていない」を選択してください。**

＜月曜日＞
☐ 園の屋外（園庭やグラウンドなど）
☐ 公園や屋外の遊び場
☐ 自宅の敷地内（庭など）
☐ 路上（道路や路地裏など）
☐ その他の外遊びの場所
☐ この曜日は体を動かす外遊びをしていない

＜火曜日＞
☐ 園の屋外（園庭やグラウンドなど）
☐ 公園や屋外の遊び場
☐ 自宅の敷地内（庭など）
☐ 路上（道路や路地裏など）
☐ その他の外遊びの場所
☐ この曜日は体を動かす外遊びをしていない

＜水曜日＞
☐ 園の屋外（園庭やグラウンドなど）
☐ 公園や屋外の遊び場
☐ 自宅の敷地内（庭など）
☐ 路上（道路や路地裏など）
☐ その他の外遊びの場所
☐ この曜日は体を動かす外遊びをしていない

＜木曜日＞
☐ 園の屋外（園庭やグラウンドなど）
☐ 公園や屋外の遊び場
☐ 自宅の敷地内（庭など）
☐ 路上（道路や路地裏など）
☐ その他の外遊びの場所
☐ この曜日は体を動かす外遊びをしていない

＜金曜日＞
☐ 園の屋外（園庭やグラウンドなど）
☐ 公園や屋外の遊び場
☐ 自宅の敷地内（庭など）
☐ 路上（道路や路地裏など）
☐ その他の外遊びの場所
☐ この曜日は体を動かす外遊びをしていない

＜土曜日＞
☐ 園の屋外（園庭やグラウンドなど）
☐ 公園や屋外の遊び場

□ 自宅の敷地内（庭など）
□ 路上（道路や路地裏など）
□ その他の外遊びの場所
□ この曜日は体を動かす外遊びをしていない

＜日曜日＞
□ 園の屋外（園庭やグラウンドなど）
□ 公園や屋外の遊び場
□ 自宅の敷地内（庭など）
□ 路上（道路や路地裏など）
□ その他の外遊びの場所
□ この曜日は体を動かす外遊びをしていない

**質問 2_1.** 運動の習いごと以外で、お子様はどれくらいの時間、体を動かす外遊びをしていますか？ここ 1 ヶ月を思い出し、各曜日の遊んでいる場所ごとに、平均的な 1 日あたりの時間（分）を回答してください。幼稚園など園にいる時間については、あなたが把握している範囲で「園の屋外（園庭やグラウンドなど）」欄に回答してください。
例：日曜日に公園で 1 時間半遊んでいる場合、日曜日の「公園や屋外の遊び場」の欄に「90」と入力してください。
※1～480 の範囲でお答えください。※半角でご入力ください。

＜月曜日＞
園の屋外（園庭やグラウンドなど）
[　　]分　【条件】質問 2 で『月曜日』の該当項目を選択した場合。入力値は 1～480。
公園や屋外の遊び場
[　　]分　【条件】質問 2 で『月曜日』の該当項目を選択した場合。入力値は 1～480。
自宅の敷地内（庭など）
[　　]分　【条件】質問 2 で『月曜日』の該当項目を選択した場合。入力値は 1～480。
路上（道路や路地裏など）
[　　]分　【条件】質問 2 で『月曜日』の該当項目を選択した場合。入力値は 1～480。
その他の外遊びの場所
[　　]分　【条件】質問 2 で『月曜日』の該当項目を選択した場合。入力値は 1～480。

＜火曜日＞
園の屋外（園庭やグラウンドなど）
[　　]分　【条件】質問 2 で『火曜日』の該当項目を選択した場合。入力値は 1～480。
公園や屋外の遊び場
[　　]分　【条件】質問 2 で『火曜日』の該当項目を選択した場合。入力値は 1～480。
自宅の敷地内（庭など）
[　　]分　【条件】質問 2 で『火曜日』の該当項目を選択した場合。入力値は 1～480。
路上（道路や路地裏など）
[　　]分　【条件】質問 2 で『火曜日』の該当項目を選択した場合。入力値は 1～480。
その他の外遊びの場所
[　　]分　【条件】質問 2 で『火曜日』の該当項目を選択した場合。入力値は 1～480。

＜水曜日＞
園の屋外（園庭やグラウンドなど）
[　　]分　【条件】質問 2 で『水曜日』の該当項目を選択した場合。入力値は 1～480。
公園や屋外の遊び場
[　　]分　【条件】質問 2 で『水曜日』の該当項目を選択した場合。入力値は 1～480。
自宅の敷地内（庭など）
[　　]分　【条件】質問 2 で『水曜日』の該当項目を選択した場合。入力値は 1～480。
路上（道路や路地裏など）
[　　]分　【条件】質問 2 で『水曜日』の該当項目を選択した場合。入力値は 1～480。

その他の外遊びの場所
□分　【条件】質問 2 で『水曜日』の該当項目を選択した場合。入力値は 1～480。

＜木曜日＞
園の屋外（園庭やグラウンドなど）
□分　【条件】質問 2 で『木曜日』の該当項目を選択した場合。入力値は 1～480。
公園や屋外の遊び場
□分　【条件】質問 2 で『木曜日』の該当項目を選択した場合。入力値は 1～480。
自宅の敷地内（庭など）
□分　【条件】質問 2 で『木曜日』の該当項目を選択した場合。入力値は 1～480。
路上（道路や路地裏など）
□分　【条件】質問 2 で『木曜日』の該当項目を選択した場合。入力値は 1～480。
その他の外遊びの場所
□分　【条件】質問 2 で『木曜日』の該当項目を選択した場合。入力値は 1～480。

＜金曜日＞
園の屋外（園庭やグラウンドなど）
□分　【条件】質問 2 で『金曜日』の該当項目を選択した場合。入力値は 1～480。
公園や屋外の遊び場
□分　【条件】質問 2 で『金曜日』の該当項目を選択した場合。入力値は 1～480。
自宅の敷地内（庭など）
□分　【条件】質問 2 で『金曜日』の該当項目を選択した場合。入力値は 1～480。
路上（道路や路地裏など）
□分　【条件】質問 2 で『金曜日』の該当項目を選択した場合。入力値は 1～480。
その他の外遊びの場所
□分　【条件】質問 2 で『金曜日』の該当項目を選択した場合。入力値は 1～480。

＜土曜日＞
園の屋外（園庭やグラウンドなど）
□分　【条件】質問 2 で『土曜日』の該当項目を選択した場合。入力値は 1～480。
公園や屋外の遊び場
□分　【条件】質問 2 で『土曜日』の該当項目を選択した場合。入力値は 1～480。
自宅の敷地内（庭など）
□分　【条件】質問 2 で『土曜日』の該当項目を選択した場合。入力値は 1～480。
路上（道路や路地裏など）
□分　【条件】質問 2 で『土曜日』の該当項目を選択した場合。入力値は 1～480。
その他の外遊びの場所
□分　【条件】質問 2 で『土曜日』の該当項目を選択した場合。入力値は 1～480。

＜日曜日＞
園の屋外（園庭やグラウンドなど）
□分　【条件】質問 2 で『日曜日』の該当項目を選択した場合。入力値は 1～480。
公園や屋外の遊び場
□分　【条件】質問 2 で『日曜日』の該当項目を選択した場合。入力値は 1～480。
自宅の敷地内（庭など）
□分　【条件】質問 2 で『日曜日』の該当項目を選択した場合。入力値は 1～480。
路上（道路や路地裏など）
□分　【条件】質問 2 で『日曜日』の該当項目を選択した場合。入力値は 1～480。
その他の外遊びの場所
□分　【条件】質問 2 で『日曜日』の該当項目を選択した場合。入力値は 1～480。

**質問 2_2.** これらの外遊びのうち、息がはずむ程度のきつさの運動時間は何分くらいありますか？**ない場合は「0 分」と入力してください。**
※0～480 の範囲でお答えください。※半角でご入力ください。

月曜日[　　　]分　【条件】質問 2 で『月曜日』の遊び場所を選択した場合。入力値は 0～480。

火曜日[　　　]分　【条件】質問 2 で『火曜日』の遊び場所を選択した場合。入力値は 0～480。

水曜日[　　　]分　【条件】質問 2 で『水曜日』の遊び場所を選択した場合。入力値は 0～480。

木曜日[　　　]分　【条件】質問 2 で『木曜日』の遊び場所を選択した場合。入力値は 0～480。

金曜日[　　　]分　【条件】質問 2 で『金曜日』の遊び場所を選択した場合。入力値は 0～480。

土曜日[　　　]分　【条件】質問 2 で『土曜日』の遊び場所を選択した場合。入力値は 0～480。

日曜日[　　　]分　【条件】質問 2 で『日曜日』の遊び場所を選択した場合。入力値は 0～480。

**質問 3.** 運動の習いごと以外で、お子様は室内や屋内施設で体を動かす遊びをしていますか？ここ 1 ヶ月を思い出し、曜日ごとに遊んでいる項目をすべて選択してください。幼稚園など園にいる時間については、あなたが把握している範囲で「園の屋内での運動遊び（ホールや遊戯室など）」を選択してください。**室内で体を動かして遊ばない曜日は「この曜日は室内で体を動かして遊ばない」を選択してください。**

＜月曜日＞
- ☐ 園の屋内での運動遊び（ホールや遊戯室など）
- ☐ 室内でゲーム機や動画を見ながら体を動かす運動（例：Wii Sports、ニンテンドースイッチなどのゲーム、YouTube など）
- ☐ 室内で体を動かす運動（例：ゲーム機や動画以外。走り回る、親子で体を動かして遊ぶなど）
- ☐ 屋内運動施設（例：体育館・プール、児童館、商業施設の遊び場など）
- ☐ その他の室内で体を動かす場面
- ☐ この曜日は室内で体を動かして遊ばない

＜火曜日＞
- ☐ 園の屋内での運動遊び（ホールや遊戯室など）
- ☐ 室内でゲーム機や動画を見ながら体を動かす運動（例：Wii Sports、ニンテンドースイッチなどのゲーム、YouTube など）
- ☐ 室内で体を動かす運動（例：ゲーム機や動画以外。走り回る、親子で体を動かして遊ぶなど）
- ☐ 屋内運動施設（例：体育館・プール、児童館、商業施設の遊び場など）
- ☐ その他の室内で体を動かす場面
- ☐ この曜日は室内で体を動かして遊ばない

＜水曜日＞
- ☐ 園の屋内での運動遊び（ホールや遊戯室など）
- ☐ 室内でゲーム機や動画を見ながら体を動かす運動（例：Wii Sports、ニンテンドースイッチなどのゲーム、YouTube など）
- ☐ 室内で体を動かす運動（例：ゲーム機や動画以外。走り回る、親子で体を動かして遊ぶなど）
- ☐ 屋内運動施設（例：体育館・プール、児童館、商業施設の遊び場など）
- ☐ その他の室内で体を動かす場面
- ☐ この曜日は室内で体を動かして遊ばない

＜木曜日＞
- ☐ 園の屋内での運動遊び（ホールや遊戯室など）
- ☐ 室内でゲーム機や動画を見ながら体を動かす運動（例：Wii Sports、ニンテンドースイッチなどのゲーム、YouTube など）
- ☐ 室内で体を動かす運動（例：ゲーム機や動画以外。走り回る、親子で体を動かして遊ぶなど）
- ☐ 屋内運動施設（例：体育館・プール、児童館、商業施設の遊び場など）

□ その他の室内で体を動かす場面
□ この曜日は室内で体を動かして遊ばない

＜金曜日＞
□ 園の屋内での運動遊び（ホールや遊戯室など）
□ 室内でゲーム機や動画を見ながら体を動かす運動（例：Wii Sports、ニンテンドースイッチなどのゲーム、YouTube など）
□ 室内で体を動かす運動（例：ゲーム機や動画以外。走り回る、親子で体を動かして遊ぶなど）
□ 屋内運動施設（例：体育館・プール、児童館、商業施設の遊び場など）
□ その他の室内で体を動かす場面
□ この曜日は室内で体を動かして遊ばない

＜土曜日＞
□ 園の屋内での運動遊び（ホールや遊戯室など）
□ 室内でゲーム機や動画を見ながら体を動かす運動（例：Wii Sports、ニンテンドースイッチなどのゲーム、YouTube など）
□ 室内で体を動かす運動（例：ゲーム機や動画以外。走り回る、親子で体を動かして遊ぶなど）
□ 屋内運動施設（例：体育館・プール、児童館、商業施設の遊び場など）
□ その他の室内で体を動かす場面
□ この曜日は室内で体を動かして遊ばない

＜日曜日＞
□ 園の屋内での運動遊び（ホールや遊戯室など）
□ 室内でゲーム機や動画を見ながら体を動かす運動（例：Wii Sports、ニンテンドースイッチなどのゲーム、YouTube など）
□ 室内で体を動かす運動（例：ゲーム機や動画以外。走り回る、親子で体を動かして遊ぶなど）
□ 屋内運動施設（例：体育館・プール、児童館、商業施設の遊び場など）
□ その他の室内で体を動かす場面
□ この曜日は室内で体を動かして遊ばない

**質問 3_1.** 運動の習いごと以外で、お子様はどれくらいの時間、室内や屋内施設で体を動かす遊びをしていますか？ここ 1 ヶ月を思い出し、各曜日の遊んでいる項目ごとに、平均的な 1 日あたりの時間（分）を回答してください。幼稚園など園にいる時間については、あなたが把握している範囲で「園の屋内での運動遊び（ホールや遊戯室など）」欄に回答してください。
※1～480 の範囲でお答えください。※半角でご入力ください。

＜月曜日＞
園の屋内での運動遊び（ホールや遊戯室など）
____分　【条件】質問 3 で『月曜日』の該当項目を選択した場合。入力値は 1～480。
室内でゲーム機や動画を見ながら体を動かす運動（例：Wii Sports、ニンテンドースイッチなどのゲーム、YouTube など）
____分　【条件】質問 3 で『月曜日』の該当項目を選択した場合。入力値は 1～480。
室内で体を動かす運動（例：ゲーム機や動画以外。走り回る、親子で体を動かして遊ぶなど）
____分　【条件】質問 3 で『月曜日』の該当項目を選択した場合。入力値は 1～480。
屋内運動施設（例：体育館・プール、児童館、商業施設の遊び場など）
____分　【条件】質問 3 で『月曜日』の該当項目を選択した場合。入力値は 1～480。
その他の室内で体を動かす場面
____分　【条件】質問 3 で『月曜日』の該当項目を選択した場合。入力値は 1～480。

＜火曜日＞
園の屋内での運動遊び（ホールや遊戯室など）
____分　【条件】質問 3 で『火曜日』の該当項目を選択した場合。入力値は 1～480。
室内でゲーム機や動画を見ながら体を動かす運動（例：Wii Sports、ニンテンドースイッチなどのゲーム、YouTube など）

[　　]分　【条件】質問 3 で『火曜日』の該当項目を選択した場合。入力値は 1～480。
室内で体を動かす運動（例：ゲーム機や動画以外。走り回る、親子で体を動かして遊ぶなど）
[　　]分　【条件】質問 3 で『火曜日』の該当項目を選択した場合。入力値は 1～480。
屋内運動施設（例：体育館・プール、児童館、商業施設の遊び場など）
[　　]分　【条件】質問 3 で『火曜日』の該当項目を選択した場合。入力値は 1～480。
その他の室内で体を動かす場面
[　　]分　【条件】質問 3 で『火曜日』の該当項目を選択した場合。入力値は 1～480。

＜水曜日＞
園の屋内での運動遊び（ホールや遊戯室など）
[　　]分　【条件】質問 3 で『水曜日』の該当項目を選択した場合。入力値は 1～480。
室内でゲーム機や動画を見ながら体を動かす運動（例：Wii Sports、ニンテンドースイッチなどのゲーム、YouTube など）
[　　]分　【条件】質問 3 で『水曜日』の該当項目を選択した場合。入力値は 1～480。
室内で体を動かす運動（例：ゲーム機や動画以外。走り回る、親子で体を動かして遊ぶなど）
[　　]分　【条件】質問 3 で『水曜日』の該当項目を選択した場合。入力値は 1～480。
屋内運動施設（例：体育館・プール、児童館、商業施設の遊び場など）
[　　]分　【条件】質問 3 で『水曜日』の該当項目を選択した場合。入力値は 1～480。
その他の室内で体を動かす場面
[　　]分　【条件】質問 3 で『水曜日』の該当項目を選択した場合。入力値は 1～480。

＜木曜日＞
園の屋内での運動遊び（ホールや遊戯室など）
[　　]分　【条件】質問 3 で『木曜日』の該当項目を選択した場合。入力値は 1～480。
室内でゲーム機や動画を見ながら体を動かす運動（例：Wii Sports、ニンテンドースイッチなどのゲーム、YouTube など）
[　　]分　【条件】質問 3 で『木曜日』の該当項目を選択した場合。入力値は 1～480。
室内で体を動かす運動（例：ゲーム機や動画以外。走り回る、親子で体を動かして遊ぶなど）
[　　]分　【条件】質問 3 で『木曜日』の該当項目を選択した場合。入力値は 1～480。
屋内運動施設（例：体育館・プール、児童館、商業施設の遊び場など）
[　　]分　【条件】質問 3 で『木曜日』の該当項目を選択した場合。入力値は 1～480。
その他の室内で体を動かす場面
[　　]分　【条件】質問 3 で『木曜日』の該当項目を選択した場合。入力値は 1～480。

＜金曜日＞
園の屋内での運動遊び（ホールや遊戯室など）
[　　]分　【条件】質問 3 で『金曜日』の該当項目を選択した場合。入力値は 1～480。
室内でゲーム機や動画を見ながら体を動かす運動（例：Wii Sports、ニンテンドースイッチなどのゲーム、YouTube など）
[　　]分　【条件】質問 3 で『金曜日』の該当項目を選択した場合。入力値は 1～480。
室内で体を動かす運動（例：ゲーム機や動画以外。走り回る、親子で体を動かして遊ぶなど）
[　　]分　【条件】質問 3 で『金曜日』の該当項目を選択した場合。入力値は 1～480。
屋内運動施設（例：体育館・プール、児童館、商業施設の遊び場など）
[　　]分　【条件】質問 3 で『金曜日』の該当項目を選択した場合。入力値は 1～480。
その他の室内で体を動かす場面
[　　]分　【条件】質問 3 で『金曜日』の該当項目を選択した場合。入力値は 1～480。

＜土曜日＞
園の屋内での運動遊び（ホールや遊戯室など）
[　　]分　【条件】質問 3 で『土曜日』の該当項目を選択した場合。入力値は 1～480。
室内でゲーム機や動画を見ながら体を動かす運動（例：Wii Sports、ニンテンドースイッチなどのゲーム、YouTube など）
[　　]分　【条件】質問 3 で『土曜日』の該当項目を選択した場合。入力値は 1～480。
室内で体を動かす運動（例：ゲーム機や動画以外。走り回る、親子で体を動かして遊ぶなど）

[ ]分　【条件】質問 3 で『土曜日』の該当項目を選択した場合。入力値は 1～480。
屋内運動施設（例：体育館・プール、児童館、商業施設の遊び場など）
[ ]分　【条件】質問 3 で『土曜日』の該当項目を選択した場合。入力値は 1～480。
その他の室内で体を動かす場面
[ ]分　【条件】質問 3 で『土曜日』の該当項目を選択した場合。入力値は 1～480。

＜日曜日＞
園の屋内での運動遊び（ホールや遊戯室など）
[ ]分　【条件】質問 3 で『日曜日』の該当項目を選択した場合。入力値は 1～480。
室内でゲーム機や動画を見ながら体を動かす運動（例：Wii Sports、ニンテンドースイッチなどのゲーム、YouTube など）
[ ]分　【条件】質問 3 で『日曜日』の該当項目を選択した場合。入力値は 1～480。
室内で体を動かす運動（例：ゲーム機や動画以外。走り回る、親子で体を動かして遊ぶなど）
[ ]分　【条件】質問 3 で『日曜日』の該当項目を選択した場合。入力値は 1～480。
屋内運動施設（例：体育館・プール、児童館、商業施設の遊び場など）
[ ]分　【条件】質問 3 で『日曜日』の該当項目を選択した場合。入力値は 1～480。
その他の室内で体を動かす場面
[ ]分　【条件】質問 3 で『日曜日』の該当項目を選択した場合。入力値は 1～480。

**質問 3_2.** これらの室内や屋内施設で体を動かす遊びのうち、息がはずむ程度のきつさの運動時間は何分くらいありますか？**ない場合は「0 分」と入力してください。**
※0～480 の範囲でお答えください。※半角でご入力ください。

月曜日[ ]分　【条件】質問 3 で『月曜日』の遊び項目を選択した場合。入力値は 0～480。

火曜日[ ]分　【条件】質問 3 で『火曜日』の遊び項目を選択した場合。入力値は 0～480。

水曜日[ ]分　【条件】質問 3 で『水曜日』の遊び項目を選択した場合。入力値は 0～480。

木曜日[ ]分　【条件】質問 3 で『木曜日』の遊び項目を選択した場合。入力値は 0～480。

金曜日[ ]分　【条件】質問 3 で『金曜日』の遊び項目を選択した場合。入力値は 0～480。

土曜日[ ]分　【条件】質問 3 で『土曜日』の遊び項目を選択した場合。入力値は 0～480。

日曜日[ ]分　【条件】質問 3 で『日曜日』の遊び項目を選択した場合。入力値は 0～480。

#### お子様の生活習慣についてお答えください

**質問 4.** お子様の平日（月～金）と休日（土・日）の就寝時刻と起床時刻を 24 時間表記で教えてください。（例：午後 10 時⇒22 時）

＜平日＞就寝時刻
[ ]時　[ ]分　【条件】時（18 時から 1 時間刻み）。分（5 分刻み）。選択式。

＜平日＞起床時刻
[ ]時　[ ]分　【条件】時（5 時から 1 時間刻み）。分（5 分刻み）。選択式。

＜休日＞就寝時刻
[ ]時　[ ]分　【条件】時（18 時から 1 時間刻み）。分（5 分刻み）。選択式。

＜休日＞起床時刻

☐時　☐分　【条件】時（5 時から 1 時間刻み）。分（5 分刻み）。選択式。

**質問 5.** ここ 1 ヶ月、お子様は、テレビや DVD をみたり、パソコン、ゲーム（テレビ、パソコン、携帯式のゲーム機などを含む）、スマートフォンなどを使用したりする時間は 1 日あたりどのくらいですか。（ただし、Wii Sports などの体を動かすゲーム、YouTube などの動画を見ながら体を動かす時間は含めません）

＜平日（月～金）＞

○ 30 分未満
○ 30 分～1 時間未満
○ 1～2 時間未満
○ 2～3 時間未満
○ 3～4 時間未満
○ 4～5 時間未満
○ 5 時間以上

＜休日（土・日）＞

○ 30 分未満
○ 30 分～1 時間未満
○ 1～2 時間未満
○ 2～3 時間未満
○ 3～4 時間未満
○ 4～5 時間未満
○ 5 時間以上

[回答を提出する]

**eMaterial 8. SSF-PAQ-24 (24-hour version) — electronic, English translation**

*Note: This English version was translated by the authors to help readers understand the original Japanese questionnaire. It has not undergone a formal translation and cross-cultural adaptation process.*

**Questionnaire on Physical Activity, Sleep, and Screen Time in Preschool Children (24-Hour Version)**

#### SSF Preschoolers' Activity Questionnaire-24h (SSF-PAQ-24)

**Q1.** Over the past month, has your child taken any organized sports or exercise lessons? (Do not include time spent at preschool: kindergarten, nursery school, or center for early childhood education and care. However, do include organized sports or exercise lessons that your family voluntarily arranges after preschool hours using the preschool's facilities or grounds.) (Select one)

○ Yes
○ No [If "No" is selected, skip to Question 2]

**Q1_1.** On which days of the week does your child take organized sports or exercise lessons? (Select all that apply)

☐ Monday ☐ Tuesday ☐ Wednesday ☐ Thursday ☐ Friday ☐ Saturday ☐ Sunday

**Q1_2.** Thinking back over the past month, please enter your child's average daily time (minutes) spent in organized sports or exercise lessons for each day of the week.

*Enter a value between 1 and 480. *Enter numbers as half-width (single-byte) digits.

Monday ______ min [Shown if "Monday" was selected in Q1_1; range: 1–480]
Tuesday ______ min [Shown if "Tuesday" was selected in Q1_1; range: 1–480]
Wednesday ______ min [Shown if "Wednesday" was selected in Q1_1; range: 1–480]
Thursday ______ min [Shown if "Thursday" was selected in Q1_1; range: 1–480]
Friday ______ min [Shown if "Friday" was selected in Q1_1; range: 1–480]
Saturday ______ min [Shown if "Saturday" was selected in Q1_1; range: 1–480]
Sunday ______ min [Shown if "Sunday" was selected in Q1_1; range: 1–480]

**Q1_3.** Of these lessons, about how many minutes per day were spent in activity that makes your child breathe harder? **Enter "0" if none.**

*Enter a value between 0 and 480. *Enter numbers as half-width (single-byte) digits.

Monday ______ min [Shown if "Monday" was selected in Q1_1; range: 0–480]
Tuesday ______ min [Shown if "Tuesday" was selected in Q1_1; range: 0–480]
Wednesday ______ min [Shown if "Wednesday" was selected in Q1_1; range: 0–480]
Thursday ______ min [Shown if "Thursday" was selected in Q1_1; range: 0–480]
Friday ______ min [Shown if "Friday" was selected in Q1_1; range: 0–480]
Saturday ______ min [Shown if "Saturday" was selected in Q1_1; range: 0–480]
Sunday ______ min [Shown if "Sunday" was selected in Q1_1; range: 0–480]

**The following questions ask about your child's active play time outside of organized sports or exercise lessons.**

**Q2.** Outside of organized sports or exercise lessons, does your child engage in outdoor active play? Thinking back over the past month, select all the locations where your child plays, for each day of the week. For time spent at preschool, select “Preschool outdoor areas (playground, sports ground, etc.)” to the best of your knowledge. **For days with no outdoor active play, select “No outdoor active play on this day.”**

**<Monday>**
- ☐ Preschool outdoor areas (playground, sports ground, etc.)
- ☐ Parks or outdoor playgrounds
- ☐ Home premises (yard, etc.)
- ☐ Streets (roads, alleys, etc.)
- ☐ Other outdoor play locations
- ☐ No outdoor active play on this day

**<Tuesday>**
- ☐ Preschool outdoor areas (playground, sports ground, etc.)
- ☐ Parks or outdoor playgrounds
- ☐ Home premises (yard, etc.)
- ☐ Streets (roads, alleys, etc.)
- ☐ Other outdoor play locations
- ☐ No outdoor active play on this day

**<Wednesday>**
- ☐ Preschool outdoor areas (playground, sports ground, etc.)
- ☐ Parks or outdoor playgrounds
- ☐ Home premises (yard, etc.)
- ☐ Streets (roads, alleys, etc.)
- ☐ Other outdoor play locations
- ☐ No outdoor active play on this day

**<Thursday>**
- ☐ Preschool outdoor areas (playground, sports ground, etc.)
- ☐ Parks or outdoor playgrounds
- ☐ Home premises (yard, etc.)
- ☐ Streets (roads, alleys, etc.)
- ☐ Other outdoor play locations
- ☐ No outdoor active play on this day

**<Friday>**
- ☐ Preschool outdoor areas (playground, sports ground, etc.)
- ☐ Parks or outdoor playgrounds
- ☐ Home premises (yard, etc.)
- ☐ Streets (roads, alleys, etc.)
- ☐ Other outdoor play locations
- ☐ No outdoor active play on this day

**<Saturday>**
- ☐ Preschool outdoor areas (playground, sports ground, etc.)
- ☐ Parks or outdoor playgrounds
- ☐ Home premises (yard, etc.)
- ☐ Streets (roads, alleys, etc.)
- ☐ Other outdoor play locations
- ☐ No outdoor active play on this day

eMaterial 8. Continued.

**<Sunday>**

- ☐ Preschool outdoor areas (playground, sports ground, etc.)
- ☐ Parks or outdoor playgrounds
- ☐ Home premises (yard, etc.)
- ☐ Streets (roads, alleys, etc.)
- ☐ Other outdoor play locations
- ☐ No outdoor active play on this day

**Q2_1.** Outside of organized sports or exercise lessons, how much time does your child spend in outdoor active play? Thinking back over the past month, please enter the average daily time (minutes) for each play location on each day of the week. For time spent at preschool, answer under "Preschool outdoor areas (playground, sports ground, etc.)" to the best of your knowledge.

Example: If your child plays at a park for an hour and a half on Sundays, enter "90" in the "Parks or outdoor playgrounds" field under Sunday.

*Enter a value between 1 and 480. *Enter numbers as half-width (single-byte) digits.

**<Monday>**

Preschool outdoor areas (playground, sports ground, etc.)
_______ min [Shown if this location was selected for Monday in Q2; range: 1–480]

Parks or outdoor playgrounds
_______ min [Shown if this location was selected for Monday in Q2; range: 1–480]

Home premises (yard, etc.)
_______ min [Shown if this location was selected for Monday in Q2; range: 1–480]

Streets (roads, alleys, etc.)
_______ min [Shown if this location was selected for Monday in Q2; range: 1–480]

Other outdoor play locations
_______ min [Shown if this location was selected for Monday in Q2; range: 1–480]

**<Tuesday>**

Preschool outdoor areas (playground, sports ground, etc.)
_______ min [Shown if this location was selected for Tuesday in Q2; range: 1–480]

Parks or outdoor playgrounds
_______ min [Shown if this location was selected for Tuesday in Q2; range: 1–480]

Home premises (yard, etc.)
_______ min [Shown if this location was selected for Tuesday in Q2; range: 1–480]

Streets (roads, alleys, etc.)
_______ min [Shown if this location was selected for Tuesday in Q2; range: 1–480]

Other outdoor play locations
_______ min [Shown if this location was selected for Tuesday in Q2; range: 1–480]

**<Wednesday>**

Preschool outdoor areas (playground, sports ground, etc.)
_______ min [Shown if this location was selected for Wednesday in Q2; range: 1–480]

Parks or outdoor playgrounds
_______ min [Shown if this location was selected for Wednesday in Q2; range: 1–480]

Home premises (yard, etc.)
_______ min [Shown if this location was selected for Wednesday in Q2; range: 1–480]

Streets (roads, alleys, etc.)
_______ min [Shown if this location was selected for Wednesday in Q2; range: 1–480]

Other outdoor play locations
_______ min [Shown if this location was selected for Wednesday in Q2; range: 1–480]

eMaterial 8. Continued.

**<Thursday>**

Preschool outdoor areas (playground, sports ground, etc.)

_______ min [Shown if this location was selected for Thursday in Q2; range: 1–480]

Parks or outdoor playgrounds

_______ min [Shown if this location was selected for Thursday in Q2; range: 1–480]

Home premises (yard, etc.)

_______ min [Shown if this location was selected for Thursday in Q2; range: 1–480]

Streets (roads, alleys, etc.)

_______ min [Shown if this location was selected for Thursday in Q2; range: 1–480]

Other outdoor play locations

_______ min [Shown if this location was selected for Thursday in Q2; range: 1–480]

**<Friday>**

Preschool outdoor areas (playground, sports ground, etc.)

_______ min [Shown if this location was selected for Friday in Q2; range: 1–480]

Parks or outdoor playgrounds

_______ min [Shown if this location was selected for Friday in Q2; range: 1–480]

Home premises (yard, etc.)

_______ min [Shown if this location was selected for Friday in Q2; range: 1–480]

Streets (roads, alleys, etc.)

_______ min [Shown if this location was selected for Friday in Q2; range: 1–480]

Other outdoor play locations

_______ min [Shown if this location was selected for Friday in Q2; range: 1–480]

**<Saturday>**

Preschool outdoor areas (playground, sports ground, etc.)

_______ min [Shown if this location was selected for Saturday in Q2; range: 1–480]

Parks or outdoor playgrounds

_______ min [Shown if this location was selected for Saturday in Q2; range: 1–480]

Home premises (yard, etc.)

_______ min [Shown if this location was selected for Saturday in Q2; range: 1–480]

Streets (roads, alleys, etc.)

_______ min [Shown if this location was selected for Saturday in Q2; range: 1–480]

Other outdoor play locations

_______ min [Shown if this location was selected for Saturday in Q2; range: 1–480]

**<Sunday>**

Preschool outdoor areas (playground, sports ground, etc.)

_______ min [Shown if this location was selected for Sunday in Q2; range: 1–480]

Parks or outdoor playgrounds

_______ min [Shown if this location was selected for Sunday in Q2; range: 1–480]

Home premises (yard, etc.)

_______ min [Shown if this location was selected for Sunday in Q2; range: 1–480]

Streets (roads, alleys, etc.)

_______ min [Shown if this location was selected for Sunday in Q2; range: 1–480]

Other outdoor play locations

_______ min [Shown if this location was selected for Sunday in Q2; range: 1–480]

**Q2_2.** Of this outdoor play, about how many minutes per day were spent in activity that makes your child breathe harder? **Enter “0” if none.**

*Enter a value between 0 and 480. *Enter numbers as half-width (single-byte) digits.

Monday _______ min [Shown if any location was selected for Monday in Q2; range: 0–480]

Tuesday ______ min [Shown if any location was selected for Tuesday in Q2; range: 0–480]

Wednesday ______ min [Shown if any location was selected for Wednesday in Q2; range: 0–480]

Thursday ______ min [Shown if any location was selected for Thursday in Q2; range: 0–480]

Friday ______ min [Shown if any location was selected for Friday in Q2; range: 0–480]

Saturday ______ min [Shown if any location was selected for Saturday in Q2; range: 0–480]

Sunday ______ min [Shown if any location was selected for Sunday in Q2; range: 0–480]

**Q3.** Outside of organized sports or exercise lessons, does your child engage in active play indoors or at indoor facilities? Thinking back over the past month, select all applicable items for each day of the week. For time spent at preschool, select “Indoor active play at preschool (hall, playroom, etc.)” to the best of your knowledge. **For days with no indoor active play, select “No indoor active play on this day.”**

**<Monday>**

☐ Indoor active play at preschool (hall, playroom, etc.)
☐ Active play indoors with video games or videos (e.g., Wii Sports, Nintendo Switch games, YouTube)
☐ Active play indoors without games or videos (e.g., running around, active play with parents)
☐ Indoor exercise facilities (e.g., gymnasium, swimming pool, children’s center, indoor playgrounds in commercial facilities)
☐ Other indoor active play situations
☐ No indoor active play on this day

**<Tuesday>**

☐ Indoor active play at preschool (hall, playroom, etc.)
☐ Active play indoors with video games or videos (e.g., Wii Sports, Nintendo Switch games, YouTube)
☐ Active play indoors without games or videos (e.g., running around, active play with parents)
☐ Indoor exercise facilities (e.g., gymnasium, swimming pool, children’s center, indoor playgrounds in commercial facilities)
☐ Other indoor active play situations
☐ No indoor active play on this day

**<Wednesday>**

☐ Indoor active play at preschool (hall, playroom, etc.)
☐ Active play indoors with video games or videos (e.g., Wii Sports, Nintendo Switch games, YouTube)
☐ Active play indoors without games or videos (e.g., running around, active play with parents)
☐ Indoor exercise facilities (e.g., gymnasium, swimming pool, children’s center, indoor playgrounds in commercial facilities)
☐ Other indoor active play situations
☐ No indoor active play on this day

**<Thursday>**

☐ Indoor active play at preschool (hall, playroom, etc.)
☐ Active play indoors with video games or videos (e.g., Wii Sports, Nintendo Switch games, YouTube)
☐ Active play indoors without games or videos (e.g., running around, active play with parents)
☐ Indoor exercise facilities (e.g., gymnasium, swimming pool, children’s center, indoor playgrounds in commercial facilities)
☐ Other indoor active play situations
☐ No indoor active play on this day

**<Friday>**

- ☐ Indoor active play at preschool (hall, playroom, etc.)
- ☐ Active play indoors with video games or videos (e.g., Wii Sports, Nintendo Switch games, YouTube)
- ☐ Active play indoors without games or videos (e.g., running around, active play with parents)
- ☐ Indoor exercise facilities (e.g., gymnasium, swimming pool, children's center, indoor playgrounds in commercial facilities)
- ☐ Other indoor active play situations
- ☐ No indoor active play on this day

**<Saturday>**

- ☐ Indoor active play at preschool (hall, playroom, etc.)
- ☐ Active play indoors with video games or videos (e.g., Wii Sports, Nintendo Switch games, YouTube)
- ☐ Active play indoors without games or videos (e.g., running around, active play with parents)
- ☐ Indoor exercise facilities (e.g., gymnasium, swimming pool, children's center, indoor playgrounds in commercial facilities)
- ☐ Other indoor active play situations
- ☐ No indoor active play on this day

**<Sunday>**

- ☐ Indoor active play at preschool (hall, playroom, etc.)
- ☐ Active play indoors with video games or videos (e.g., Wii Sports, Nintendo Switch games, YouTube)
- ☐ Active play indoors without games or videos (e.g., running around, active play with parents)
- ☐ Indoor exercise facilities (e.g., gymnasium, swimming pool, children's center, indoor playgrounds in commercial facilities)
- ☐ Other indoor active play situations
- ☐ No indoor active play on this day

**Q3_1.** Outside of organized sports or exercise lessons, how much time does your child spend in active play indoors or at indoor facilities? Thinking back over the past month, please enter the average daily time (minutes) for each item on each day of the week. For time spent at preschool, answer under "Indoor active play at preschool (hall, playroom, etc.)" to the best of your knowledge.

*Enter a value between 1 and 480. *Enter numbers as half-width (single-byte) digits.

**<Monday>**

Indoor active play at preschool (hall, playroom, etc.)
_______ min [Shown if this item was selected for Monday in Q3; range: 1–480]

Active play indoors with video games or videos (e.g., Wii Sports, Nintendo Switch games, YouTube)
_______ min [Shown if this item was selected for Monday in Q3; range: 1–480]

Active play indoors without games or videos (e.g., running around, active play with parents)
_______ min [Shown if this item was selected for Monday in Q3; range: 1–480]

Indoor exercise facilities (e.g., gymnasium, swimming pool, children's center, indoor playgrounds in commercial facilities)
_______ min [Shown if this item was selected for Monday in Q3; range: 1–480]

Other indoor active play situations
_______ min [Shown if this item was selected for Monday in Q3; range: 1–480]

**<Tuesday>**

Indoor active play at preschool (hall, playroom, etc.)
_______ min [Shown if this item was selected for Tuesday in Q3; range: 1–480]

Active play indoors with video games or videos (e.g., Wii Sports, Nintendo Switch games, YouTube)
_______ min [Shown if this item was selected for Tuesday in Q3; range: 1–480]

Active play indoors without games or videos (e.g., running around, active play with parents)
_______ min [Shown if this item was selected for Tuesday in Q3; range: 1–480]

Indoor exercise facilities (e.g., gymnasium, swimming pool, children's center, indoor playgrounds in commercial facilities)
_______ min [Shown if this item was selected for Tuesday in Q3; range: 1–480]

Other indoor active play situations
_______ min [Shown if this item was selected for Tuesday in Q3; range: 1–480]

**<Wednesday>**

Indoor active play at preschool (hall, playroom, etc.)
_______ min [Shown if this item was selected for Wednesday in Q3; range: 1–480]

Active play indoors with video games or videos (e.g., Wii Sports, Nintendo Switch games, YouTube)
_______ min [Shown if this item was selected for Wednesday in Q3; range: 1–480]

Active play indoors without games or videos (e.g., running around, active play with parents)
_______ min [Shown if this item was selected for Wednesday in Q3; range: 1–480]

Indoor exercise facilities (e.g., gymnasium, swimming pool, children's center, indoor playgrounds in commercial facilities)
_______ min [Shown if this item was selected for Wednesday in Q3; range: 1–480]

Other indoor active play situations
_______ min [Shown if this item was selected for Wednesday in Q3; range: 1–480]

**<Thursday>**

Indoor active play at preschool (hall, playroom, etc.)
_______ min [Shown if this item was selected for Thursday in Q3; range: 1–480]

Active play indoors with video games or videos (e.g., Wii Sports, Nintendo Switch games, YouTube)
_______ min [Shown if this item was selected for Thursday in Q3; range: 1–480]

Active play indoors without games or videos (e.g., running around, active play with parents)
_______ min [Shown if this item was selected for Thursday in Q3; range: 1–480]

Indoor exercise facilities (e.g., gymnasium, swimming pool, children's center, indoor playgrounds in commercial facilities)
_______ min [Shown if this item was selected for Thursday in Q3; range: 1–480]

Other indoor active play situations
_______ min [Shown if this item was selected for Thursday in Q3; range: 1–480]

**<Friday>**

Indoor active play at preschool (hall, playroom, etc.)
_______ min [Shown if this item was selected for Friday in Q3; range: 1–480]

Active play indoors with video games or videos (e.g., Wii Sports, Nintendo Switch games, YouTube)
_______ min [Shown if this item was selected for Friday in Q3; range: 1–480]

Active play indoors without games or videos (e.g., running around, active play with parents)
_______ min [Shown if this item was selected for Friday in Q3; range: 1–480]

Indoor exercise facilities (e.g., gymnasium, swimming pool, children's center, indoor playgrounds in commercial facilities)
_______ min [Shown if this item was selected for Friday in Q3; range: 1–480]

Other indoor active play situations
_______ min [Shown if this item was selected for Friday in Q3; range: 1–480]

**<Saturday>**

Indoor active play at preschool (hall, playroom, etc.)
_______ min [Shown if this item was selected for Saturday in Q3; range: 1–480]

Active play indoors with video games or videos (e.g., Wii Sports, Nintendo Switch games, YouTube)
_______ min [Shown if this item was selected for Saturday in Q3; range: 1–480]

Active play indoors without games or videos (e.g., running around, active play with parents)
_______ min [Shown if this item was selected for Saturday in Q3; range: 1–480]

Indoor exercise facilities (e.g., gymnasium, swimming pool, children's center, indoor playgrounds in commercial facilities)
_______ min [Shown if this item was selected for Saturday in Q3; range: 1–480]

Other indoor active play situations
_______ min [Shown if this item was selected for Saturday in Q3; range: 1–480]

**<Sunday>**

Indoor active play at preschool (hall, playroom, etc.)
_______ min [Shown if this item was selected for Sunday in Q3; range: 1–480]

Active play indoors with video games or videos (e.g., Wii Sports, Nintendo Switch games, YouTube)
_______ min [Shown if this item was selected for Sunday in Q3; range: 1–480]

Active play indoors without games or videos (e.g., running around, active play with parents)
_______ min [Shown if this item was selected for Sunday in Q3; range: 1–480]

Indoor exercise facilities (e.g., gymnasium, swimming pool, children's center, indoor playgrounds in commercial facilities)
_______ min [Shown if this item was selected for Sunday in Q3; range: 1–480]

Other indoor active play situations
_______ min [Shown if this item was selected for Sunday in Q3; range: 1–480]

**Q3_2.** Of this indoor play, about how many minutes per day were spent in activity that makes your child breathe harder? **Enter "0" if none.**

*Enter a value between 0 and 480. *Enter numbers as half-width (single-byte) digits.

Monday _______ min [Shown if any item was selected for Monday in Q3; range: 0–480]

Tuesday _______ min [Shown if any item was selected for Tuesday in Q3; range: 0–480]

Wednesday _______ min [Shown if any item was selected for Wednesday in Q3; range: 0–480]

Thursday _______ min [Shown if any item was selected for Thursday in Q3; range: 0–480]

Friday _______ min [Shown if any item was selected for Friday in Q3; range: 0–480]

Saturday _______ min [Shown if any item was selected for Saturday in Q3; range: 0–480]

Sunday _______ min [Shown if any item was selected for Sunday in Q3; range: 0–480]

**Please answer the following questions about your child's daily routine.**

**Q4.** Please indicate your child's bedtime and wake-up time on weekdays (Mon–Fri) and weekend days (Sat–Sun), using the 24-hour clock (e.g., 10 p.m. → 22:00).

**<Weekdays> Bedtime**
_______ h _______ min [Drop-down: hour from 18:00 in 1-hour steps; minutes in 5-minute steps]

**<Weekdays> Wake-up time**
_______ h _______ min [Drop-down: hour from 5:00 in 1-hour steps; minutes in 5-minute steps]

**<Weekend days> Bedtime**
_______ h _______ min [Drop-down: hour from 18:00 in 1-hour steps; minutes in 5-minute steps]

**<Weekend days> Wake-up time**
_______ h _______ min [Drop-down: hour from 5:00 in 1-hour steps; minutes in 5-minute steps]

**Q5.** Over the past month, about how much time per day has your child spent watching TV or DVDs, or using computers, video games (including TV, computer, and handheld game consoles), smartphones, and similar devices? (Do not include active video games such as Wii Sports or time spent being physically active while watching videos such as YouTube.)

**<Weekdays (Mon–Fri)>**

- ○ Less than 30 min
- ○ 30 min to <1 h
- ○ 1 to <2 h
- ○ 2 to <3 h
- ○ 3 to <4 h
- ○ 4 to <5 h
- ○ 5 h or more

**<Weekend days (Sat–Sun)>**

- ○ Less than 30 min
- ○ 30 min to <1 h
- ○ 1 to <2 h
- ○ 2 to <3 h
- ○ 3 to <4 h
- ○ 4 to <5 h
- ○ 5 h or more

**[Submit responses]**

**eFigure 1. Bland–Altman plots comparing the SSF-PAQ with criterion measures for total screen time and total (night) sleep.** The four panels show total screen time and night sleep by version (Original and 24-h). The x-axis shows the mean of the SSF-PAQ and criterion values (min/day), and the y-axis shows the difference between them (SSF-PAQ − criterion, min/day); positive values indicate overestimation by the SSF-PAQ. The blue solid line represents the mean bias, the red dashed lines represent the 95% limits of agreement, and the green line represents the regression line used to assess proportional bias. The P value for proportional bias is shown below each panel title. The criterion for screen time was sedentary (sitting/lying) screen time from the parent-reported time-use diary (excluding screen time while standing), and the criterion for night sleep was derived by integrating the diary with accelerometer data. Within each behaviour, the y-axis scale is harmonized across versions. n denotes the number of participants in each panel. Mean bias (95% limits of agreement) was 8.2 (−72.3 to 88.8) min/day for total screen time and 10.7 (−33.2 to 54.7) min/day for night sleep in the Original version, and −22.2 (−139.2 to 94.8) and −2.6 (−55.4 to 50.2) min/day, respectively, in the 24-h version.

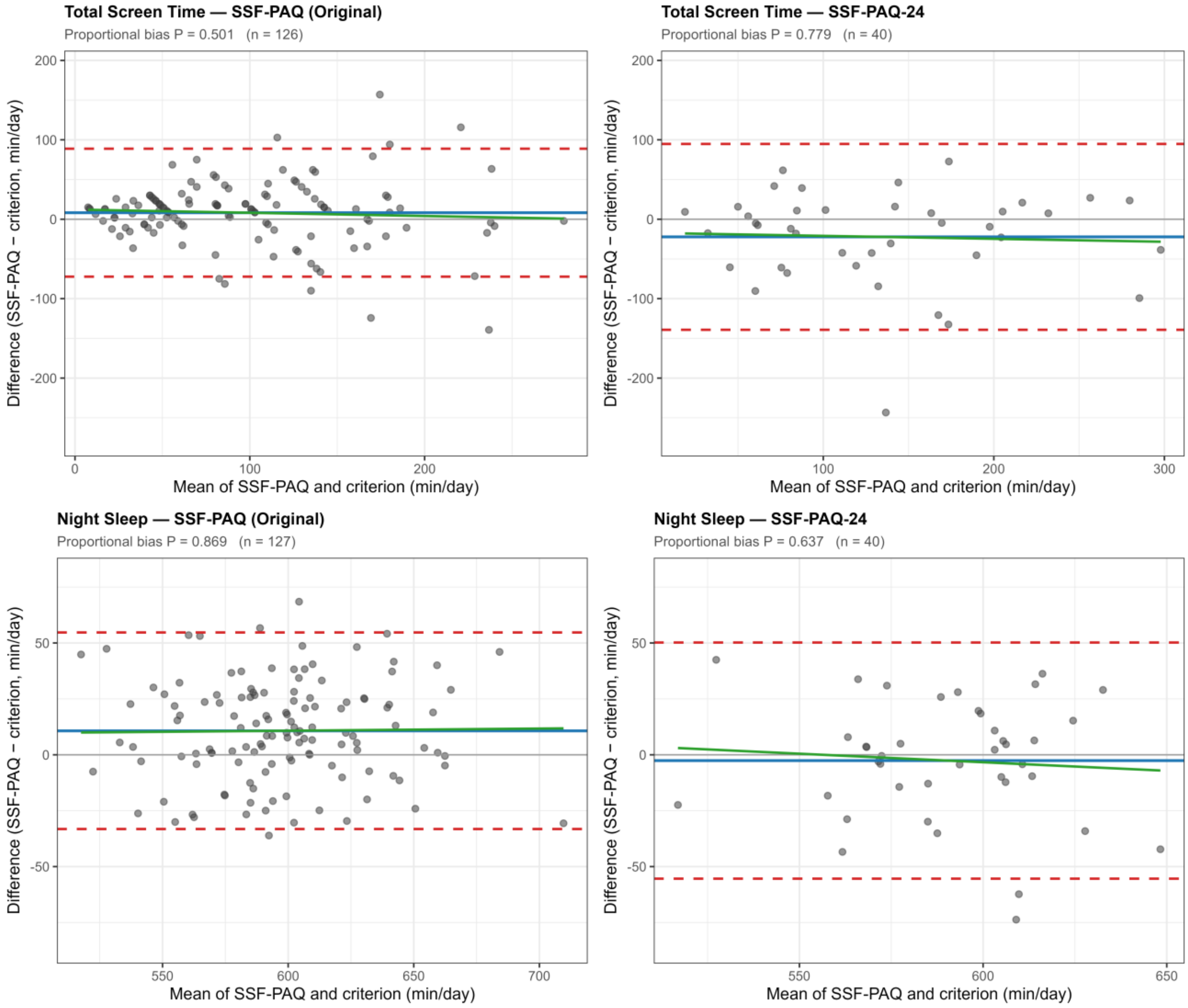

#### eTable 3. Detailed concurrent validity (overall and within-/out-of-preschool) and test–retest reliability of the SSF-PAQ by version and administration mode

##### Part A. Concurrent validity (Spearman ρ), by version and mode

| Criterion measure | SSF-PAQ item | Period | n | ρ [95% CI] | Criterion Med (Q1–Q3) | SSF-PAQ Med (Q1–Q3) | P |
|---|---|---|---|---|---|---|---|
| **SSF-PAQ Original version, overall (paper + electronic)** | | | | | | | |
| Total PA (ACC, out-of-preschool) | Organized PA and AP | Weekday | 127 | **0.38 [0.22, 0.52]** | 185 (142–226) | 35 (15–75) | NA |
| | | Weekend | 127 | 0.06 [−0.12, 0.23] | 394 (332–433) | 150 (90–222) | NA |
| | | Weekly | 127 | **0.24 [0.07, 0.39]** | 238 (200–282) | 73 (47–101) | NA |
| MVPA (ACC, out-of-preschool) | MVPA | Weekday | 127 | **0.34 [0.18, 0.49]** | 34 (23–49) | 5 (0–16) | NA |
| | | Weekend | 127 | **0.31 [0.14, 0.46]** | 70 (51–99) | 30 (10–50) | NA |
| | | Weekly | 127 | **0.29 [0.13, 0.45]** | 46 (32–61) | 13 (4–26) | NA |
| Sedentary screen time (diary) | Total screen time | Weekday | 127 | **0.81 [0.74, 0.86]** | 72 (27–117) | 90 (45–150) | < 0.001 |
| | | Weekend | 126 | **0.78 [0.70, 0.84]** | 105 (66–179) | 150 (90–150) | 0.576 |
| | | Weekly | 126 | **0.83 [0.77, 0.88]** | 87 (43–135) | 91 (58–150) | 0.001 |
| Night sleep (diary + ACC) | Night sleep | Weekday | 127 | **0.73 [0.63, 0.80]** | 585 (562–612) | 600 (570–630) | < 0.001 |
| | | Weekend | 127 | **0.50 [0.35, 0.62]** | 602 (569–637) | 600 (585–630) | 0.170 |
| | | Weekly | 127 | **0.78 [0.71, 0.84]** | 591 (570–612) | 600 (579–627) | < 0.001 |
| **SSF-PAQ Original version, paper** | | | | | | | |
| Total PA (ACC, out-of-preschool) | Organized PA and AP | Weekday | 72 | **0.25 [0.02, 0.45]** | 184 (146–223) | 34 (15–68) | NA |
| | | Weekend | 72 | 0.10 [−0.13, 0.33] | 393 (336–438) | 135 (90–192) | NA |
| | | Weekly | 72 | 0.17 [−0.06, 0.39] | 236 (207–282) | 63 (41–99) | NA |
| MVPA (ACC, out-of-preschool) | MVPA | Weekday | 72 | 0.21 [−0.02, 0.42] | 35 (22–50) | 5 (0–15) | NA |
| | | Weekend | 72 | 0.21 [−0.03, 0.42] | 67 (49–98) | 30 (10–50) | NA |
| | | Weekly | 72 | 0.15 [−0.08, 0.37] | 46 (33–62) | 11 (4–26) | NA |
| Sedentary screen time (diary) | Total screen time | Weekday | 72 | **0.78 [0.67, 0.86]** | 74 (27–111) | 90 (45–150) | 0.001 |
| | | Weekend | 71 | **0.77 [0.65, 0.85]** | 105 (62–182) | 150 (90–150) | 0.574 |
| | | Weekly | 71 | **0.82 [0.72, 0.88]** | 88 (43–136) | 107 (58–150) | 0.002 |
| Night sleep (diary + ACC) | Night sleep | Weekday | 72 | **0.72 [0.59, 0.82]** | 582 (563–614) | 600 (570–630) | < 0.001 |
| | | Weekend | 72 | **0.40 [0.19, 0.58]** | 604 (580–641) | 600 (589–630) | 0.832 |
| | | Weekly | 72 | **0.74 [0.61, 0.83]** | 588 (570–618) | 600 (587–625) | < 0.001 |
| **SSF-PAQ Original version, electronic** | | | | | | | |
| Total PA (ACC, out-of-preschool) | Organized PA and AP | Weekday | 55 | **0.56 [0.34, 0.72]** | 189 (134–234) | 42 (16–76) | NA |
| | | Weekend | 55 | 0.02 [−0.25, 0.28] | 397 (332–426) | 180 (90–240) | NA |
| | | Weekly | 55 | **0.32 [0.06, 0.54]** | 239 (193–289) | 81 (55–118) | NA |
| MVPA (ACC, out-of-preschool) | MVPA | Weekday | 55 | **0.51 [0.29, 0.69]** | 32 (23–49) | 4 (0–19) | NA |
| | | Weekend | 55 | **0.43 [0.19, 0.63]** | 79 (53–101) | 30 (10–60) | NA |
| | | Weekly | 55 | **0.47 [0.24, 0.66]** | 45 (32–59) | 14 (4–24) | NA |
| Sedentary screen time (diary) | Total screen time | Weekday | 55 | **0.84 [0.73, 0.90]** | 60 (28–123) | 90 (45–150) | 0.128 |
| | | Weekend | 55 | **0.81 [0.70, 0.89]** | 98 (68–169) | 90 (90–150) | 0.736 |
| | | Weekly | 55 | **0.84 [0.74, 0.91]** | 78 (41–131) | 90 (45–141) | 0.206 |
| Night sleep (diary + ACC) | Night sleep | Weekday | 55 | **0.76 [0.61, 0.85]** | 591 (561–608) | 600 (570–625) | 0.003 |
| | | Weekend | 55 | **0.63 [0.43, 0.76]** | 597 (564–632) | 615 (575–633) | 0.061 |
| | | Weekly | 55 | **0.85 [0.76, 0.91]** | 594 (574–606) | 604 (574–627) | < 0.001 |
| **SSF-PAQ 24-h version, overall (paper + electronic)** | | | | | | | |
| Total PA (ACC) | Organized PA and AP | Weekday | 40 | 0.20 [−0.12, 0.48] | 393 (345–426) | 97 (40–153) | NA |
| | | Weekend | 40 | −0.03 [−0.34, 0.28] | 391 (340–438) | 95 (60–161) | NA |
| | | Weekly | 40 | 0.17 [−0.15, 0.46] | 389 (342–429) | 92 (48–160) | NA |
| MVPA (ACC) | MVPA | Weekday | 40 | −0.01 [−0.32, 0.30] | 71 (46–92) | 16 (2–26) | NA |
| | | Weekend | 40 | 0.09 [−0.23, 0.39] | 93 (84–112) | 25 (9–49) | NA |
| | | Weekly | 40 | 0.06 [−0.26, 0.36] | 82 (63–100) | 21 (8–37) | NA |
| Sedentary screen time (diary) | Total screen time | Weekday | 40 | **0.64 [0.42, 0.80]** | 107 (64–213) | 90 (45–150) | 0.049 |
| | | Weekend | 40 | **0.53 [0.27, 0.72]** | 158 (109–221) | 150 (90–210) | 0.242 |
| | | Weekly | 40 | **0.68 [0.47, 0.82]** | 136 (78–208) | 107 (58–174) | 0.064 |
| Night sleep (diary + ACC) | Night sleep | Weekday | 40 | **0.57 [0.31, 0.75]** | 591 (568–607) | 595 (570–600) | 0.764 |
| | | Weekend | 40 | **0.41 [0.12, 0.64]** | 599 (575–636) | 600 (570–630) | 0.879 |
| | | Weekly | 40 | **0.63 [0.40, 0.79]** | 594 (574–610) | 590 (570–609) | 0.806 |
| **SSF-PAQ 24-h version, paper** | | | | | | | |
| Total PA (ACC) | Organized PA and AP | Weekday | 21 | 0.34 [−0.11, 0.67] | 400 (359–422) | 100 (38–150) | NA |
| | | Weekend | 21 | 0.14 [−0.31, 0.54] | 392 (354–440) | 95 (75–135) | NA |
| | | Weekly | 21 | 0.36 [−0.08, 0.69] | 395 (349–429) | 104 (46–157) | NA |

eTable 3. Continued.

| Criterion measure | SSF-PAQ item | Period | n | ρ [95% CI] | Criterion Med (Q1–Q3) | SSF-PAQ Med (Q1–Q3) | P |
|---|---|---|---|---|---|---|---|
| MVPA (ACC) | MVPA | Weekday | 21 | 0.07 [−0.37, 0.49] | 70 (55–91) | 15 (0–30) | NA |
| | | Weekend | 21 | −0.13 [−0.53, 0.32] | 94 (85–122) | 28 (10–45) | NA |
| | | Weekly | 21 | 0.16 [−0.29, 0.56] | 84 (63–94) | 21 (7–41) | NA |
| Sedentary screen time (diary) | Total screen time | Weekday | 21 | **0.71 [0.39, 0.87]** | 94 (68–199) | 90 (45–210) | 0.562 |
| | | Weekend | 21 | **0.60 [0.22, 0.82]** | 160 (120–225) | 150 (90–210) | 0.476 |
| | | Weekly | 21 | **0.77 [0.51, 0.90]** | 137 (87–200) | 107 (75–193) | 0.473 |
| Night sleep (diary + ACC) | Night sleep | Weekday | 21 | **0.65 [0.31, 0.85]** | 597 (571–611) | 600 (570–600) | 0.695 |
| | | Weekend | 21 | 0.22 [−0.23, 0.60] | 587 (576–632) | 600 (600–630) | 0.257 |
| | | Weekly | 21 | **0.70 [0.39, 0.87]** | 591 (575–612) | 601 (580–609) | 0.374 |
| **SSF-PAQ 24-h version, electronic** | | | | | | | |
| Total PA (ACC) | Organized PA and AP | Weekday | 19 | 0.09 [−0.38, 0.52] | 373 (332–428) | 90 (46–147) | NA |
| | | Weekend | 19 | −0.14 [−0.56, 0.34] | 376 (340–431) | 95 (59–175) | NA |
| | | Weekly | 19 | 0.01 [−0.45, 0.46] | 374 (336–437) | 83 (56–155) | NA |
| MVPA (ACC) | MVPA | Weekday | 19 | −0.11 [−0.54, 0.36] | 72 (46–99) | 16 (8–24) | NA |
| | | Weekend | 19 | 0.44 [−0.01, 0.75] | 91 (84–110) | 20 (10–60) | NA |
| | | Weekly | 19 | 0.06 [−0.41, 0.50] | 73 (65–100) | 21 (10–30) | NA |
| Sedentary screen time (diary) | Total screen time | Weekday | 19 | **0.59 [0.19, 0.83]** | 124 (58–238) | 90 (45–150) | 0.025 |
| | | Weekend | 19 | **0.48 [0.04, 0.77]** | 145 (105–220) | 150 (90–210) | 0.334 |
| | | Weekly | 19 | **0.60 [0.20, 0.83]** | 134 (72–226) | 107 (58–167) | 0.055 |
| Night sleep (diary + ACC) | Night sleep | Weekday | 19 | **0.54 [0.11, 0.80]** | 578 (563–603) | 570 (562–600) | 0.469 |
| | | Weekend | 19 | **0.58 [0.17, 0.82]** | 602 (569–644) | 570 (570–622) | 0.352 |
| | | Weekly | 19 | **0.71 [0.38, 0.88]** | 596 (570–604) | 570 (569–606) | 0.312 |

Abbreviations: ACC, accelerometer-derived; AP, active play; CI, confidence interval; diary, time-use diary; Med, median; MVPA, moderate-to-vigorous physical activity; organized PA, organized physical activity; PA, physical activity; Q1–Q3, interquartile range; SSF-PAQ, Sasakawa Sports Foundation Preschoolers' Activity Questionnaire.

Values are min/day. Overall combines paper and electronic modes; the paper and electronic sections comprise the children who completed the Day 7 questionnaire in that mode. Bold ρ values indicate 95% CIs excluding zero. P, Wilcoxon signed-rank test for systematic differences between the paired criterion and SSF-PAQ values; NA, not examined because the accelerometer-derived criteria are not equivalent to the questionnaire estimates.

eTable 3. Continued.

**Part B. Within- and out-of-preschool validity (SSF-PAQ 24-h version, weekday)**

| Construct | Criterion measure | SSF-PAQ-24 item | n | ρ [95% CI] | Criterion Med (Q1–Q3) | SSF-PAQ Med (Q1–Q3) |
|---|---|---|---|---|---|---|
| Out-of-preschool organized PA and AP | Total PA outside of preschool (ACC) | Organized lessons + outdoor and indoor active play, outside of preschool | 40 | 0.19 [−0.13, 0.48] | 159 (128–205) | 33 (12–60) |
| Within-preschool active play (outdoor + indoor) | Total PA at preschool (ACC) | Outdoor + indoor active play, at preschool | 40 | 0.24 [−0.08, 0.51] | 224 (167–280) | 40 (14–98) |

SSF-PAQ 24-h version (Groups 4–6; weekday; n = 40). The out-of-preschool row is organized PA and AP: organized lessons plus outdoor and indoor active play outside of preschool, summed per day and averaged over weekdays, with the same 480 min/day cap as in Part A and Table 2 (not triggered in this subsample). Organized lessons are included because they take place after the child leaves preschool and therefore fall within the out-of-preschool window defined by the diary-recorded departure time, which the accelerometer criterion also covers. The within-preschool row is active play only (the preschool outdoor and indoor rows), because the questionnaire does not ask about organized lessons at preschool. The accelerometer criterion is total physical activity accumulated at preschool (whole-day minus out-of-preschool) or outside of preschool, defined from the diary-recorded arrival and departure times. The higher-intensity (breathing-harder) portion is collected only at the aggregate outdoor/indoor level, so within-preschool MVPA cannot be derived from the questionnaire. The out-of-preschool row matches the organized PA and AP construct used in Part A and Table 2, but the estimates are still not directly comparable: they come from the smaller 24-h subsample and cover weekdays only, whereas Part A and Table 2 use the larger Original-version sample. Systematic differences were not examined for this table and no P values are reported; P is shown for Part A only. Bold ρ indicates a 95% CI excluding zero. Other abbreviations as in Part A.

**Part C. Test–retest reliability (ICC), by version and mode**

| SSF-PAQ item | Period | n | ICC [95% CI] | Day 7 Med (Q1–Q3) | Day 10 Med (Q1–Q3) |
|---|---|---|---|---|---|
| **SSF-PAQ Original version, overall (paper + electronic)** | | | | | |
| Organized PA and AP | Weekday | 79 | **0.84 [0.76, 0.89]** | 57 (20–90) | 51 (21–77) |
| | Weekend | 79 | **0.82 [0.73, 0.88]** | 155 (109–265) | 120 (92–232) |
| | Weekly | 79 | **0.84 [0.75, 0.89]** | 86 (54–121) | 75 (50–105) |
| MVPA | Weekday | 79 | **0.91 [0.87, 0.94]** | 8 (0–20) | 9 (0–20) |
| | Weekend | 79 | **0.80 [0.71, 0.87]** | 38 (10–60) | 38 (20–60) |
| | Weekly | 79 | **0.85 [0.77, 0.90]** | 17 (4–34) | 19 (12–34) |
| Total screen time | Weekday | 78 | **0.76 [0.65, 0.84]** | 90 (45–150) | 90 (45–150) |
| | Weekend | 78 | **0.79 [0.69, 0.86]** | 120 (90–150) | 120 (90–150) |
| | Weekly | 77 | **0.78 [0.68, 0.86]** | 107 (58–150) | 107 (45–150) |
| Night sleep | Weekday | 79 | **0.97 [0.95, 0.98]** | 600 (570–630) | 600 (570–630) |
| | Weekend | 79 | **0.83 [0.75, 0.89]** | 600 (585–630) | 600 (597–630) |
| | Weekly | 79 | **0.96 [0.94, 0.97]** | 604 (579–621) | 600 (579–621) |
| **SSF-PAQ Original version, paper** | | | | | |
| Organized PA and AP | Weekday | 38 | **0.79 [0.59, 0.89]** | 58 (21–88) | 38 (18–74) |
| | Weekend | 38 | **0.86 [0.74, 0.93]** | 150 (120–221) | 120 (96–210) |
| | Weekly | 38 | **0.80 [0.60, 0.90]** | 90 (56–101) | 78 (39–101) |
| MVPA | Weekday | 38 | **0.92 [0.85, 0.96]** | 10 (0–20) | 6 (0–16) |
| | Weekend | 38 | **0.68 [0.47, 0.82]** | 42 (11–60) | 42 (21–67) |
| | Weekly | 38 | **0.86 [0.74, 0.92]** | 19 (6–34) | 21 (12–34) |
| Total screen time | Weekday | 37 | **0.83 [0.70, 0.91]** | 90 (90–150) | 90 (90–150) |
| | Weekend | 37 | **0.88 [0.77, 0.93]** | 150 (90–210) | 150 (90–210) |
| | Weekly | 36 | **0.87 [0.76, 0.93]** | 107 (90–154) | 107 (90–154) |
| Night sleep | Weekday | 38 | **0.98 [0.96, 0.99]** | 600 (570–630) | 600 (570–630) |
| | Weekend | 38 | **0.81 [0.66, 0.89]** | 600 (592–630) | 600 (586–630) |
| | Weekly | 38 | **0.98 [0.96, 0.99]** | 606 (580–621) | 602 (586–621) |
| **SSF-PAQ Original version, electronic** | | | | | |
| Organized PA and AP | Weekday | 41 | **0.88 [0.78, 0.93]** | 57 (20–90) | 54 (26–88) |
| | Weekend | 41 | **0.80 [0.65, 0.89]** | 180 (90–270) | 150 (90–240) |
| | Weekly | 41 | **0.85 [0.74, 0.92]** | 86 (54–135) | 73 (56–137) |
| MVPA | Weekday | 41 | **0.90 [0.83, 0.95]** | 7 (0–30) | 12 (0–23) |
| | Weekend | 41 | **0.89 [0.81, 0.94]** | 35 (10–60) | 35 (20–60) |
| | Weekly | 41 | **0.84 [0.72, 0.91]** | 17 (4–28) | 17 (13–34) |
| Total screen time | Weekday | 41 | **0.69 [0.49, 0.82]** | 45 (15–90) | 45 (15–90) |
| | Weekend | 41 | **0.70 [0.51, 0.83]** | 90 (90–150) | 90 (45–150) |
| | Weekly | 41 | **0.70 [0.50, 0.82]** | 58 (36–107) | 58 (24–107) |
| Night sleep | Weekday | 41 | **0.94 [0.90, 0.97]** | 600 (570–620) | 600 (570–630) |
| | Weekend | 41 | **0.85 [0.73, 0.92]** | 610 (580–630) | 600 (600–645) |
| | Weekly | 41 | **0.94 [0.90, 0.97]** | 600 (579–621) | 600 (579–630) |

eTable 3. Continued.

| SSF-PAQ item | Period | n | ICC [95% CI] | Day 7 Med (Q1–Q3) | Day 10 Med (Q1–Q3) |
|---|---|---|---|---|---|
| **SSF-PAQ 24-h version, overall (paper + electronic)** | | | | | |
| Organized PA and AP | Weekday | 39 | **0.79 [0.64, 0.89]** | 96 (40–140) | 90 (54–120) |
| | Weekend | 39 | **0.72 [0.52, 0.84]** | 95 (60–162) | 80 (56–135) |
| | Weekly | 39 | **0.86 [0.74, 0.92]** | 91 (47–150) | 87 (60–116) |
| MVPA | Weekday | 39 | **0.66 [0.44, 0.80]** | 12 (1–25) | 24 (4–39) |
| | Weekend | 39 | **0.75 [0.57, 0.86]** | 25 (8–52) | 25 (14–45) |
| | Weekly | 39 | **0.82 [0.68, 0.90]** | 18 (8–33) | 24 (9–37) |
| Total screen time | Weekday | 39 | **0.80 [0.66, 0.89]** | 90 (45–150) | 90 (45–150) |
| | Weekend | 39 | **0.84 [0.72, 0.91]** | 150 (90–210) | 150 (90–210) |
| | Weekly | 39 | **0.83 [0.70, 0.91]** | 107 (58–167) | 107 (58–159) |
| Night sleep | Weekday | 39 | **0.95 [0.91, 0.97]** | 600 (570–600) | 590 (570–600) |
| | Weekend | 39 | **0.60 [0.35, 0.77]** | 600 (570–630) | 600 (600–630) |
| | Weekly | 39 | **0.91 [0.84, 0.95]** | 591 (570–609) | 599 (575–609) |
| **SSF-PAQ 24-h version, paper** | | | | | |
| Organized PA and AP | Weekday | 20 | **0.83 [0.62, 0.93]** | 100 (36–134) | 89 (43–104) |
| | Weekend | 20 | 0.49 [0.07, 0.76] | 98 (71–135) | 90 (59–112) |
| | Weekly | 20 | **0.86 [0.67, 0.95]** | 99 (45–138) | 85 (44–97) |
| MVPA | Weekday | 20 | 0.42 [0.01, 0.71] | 8 (0–25) | 21 (2–38) |
| | Weekend | 20 | **0.63 [0.28, 0.83]** | 30 (9–45) | 28 (15–38) |
| | Weekly | 20 | **0.63 [0.28, 0.83]** | 15 (6–33) | 22 (10–35) |
| Total screen time | Weekday | 20 | **0.91 [0.78, 0.96]** | 90 (45–165) | 90 (45–150) |
| | Weekend | 20 | **0.89 [0.75, 0.96]** | 150 (90–210) | 150 (90–210) |
| | Weekly | 20 | **0.92 [0.82, 0.97]** | 107 (71–178) | 107 (58–154) |
| Night sleep | Weekday | 20 | **0.92 [0.81, 0.97]** | 600 (581–600) | 600 (570–600) |
| | Weekend | 20 | 0.40 [−0.00, 0.70] | 600 (592–630) | 615 (600–630) |
| | Weekly | 20 | **0.82 [0.60, 0.92]** | 604 (580–609) | 600 (589–609) |
| **SSF-PAQ 24-h version, electronic** | | | | | |
| Organized PA and AP | Weekday | 19 | **0.73 [0.42, 0.89]** | 90 (46–147) | 90 (70–132) |
| | Weekend | 19 | **0.94 [0.86, 0.98]** | 95 (59–175) | 75 (55–135) |
| | Weekly | 19 | **0.86 [0.67, 0.94]** | 83 (56–155) | 87 (71–146) |
| MVPA | Weekday | 19 | **0.80 [0.55, 0.92]** | 16 (8–24) | 25 (7–46) |
| | Weekend | 19 | **0.89 [0.74, 0.96]** | 20 (10–60) | 20 (11–54) |
| | Weekly | 19 | **0.93 [0.83, 0.97]** | 21 (10–30) | 29 (8–40) |
| Total screen time | Weekday | 19 | **0.70 [0.36, 0.87]** | 90 (45–150) | 90 (90–150) |
| | Weekend | 19 | **0.81 [0.57, 0.92]** | 150 (90–210) | 150 (90–210) |
| | Weekly | 19 | **0.75 [0.45, 0.89]** | 107 (58–167) | 107 (90–150) |
| Night sleep | Weekday | 19 | **0.96 [0.91, 0.99]** | 570 (562–600) | 570 (562–600) |
| | Weekend | 19 | **0.73 [0.41, 0.89]** | 570 (570–622) | 600 (570–622) |
| | Weekly | 19 | **0.95 [0.87, 0.98]** | 570 (569–606) | 579 (570–600) |

ICC, intraclass correlation coefficient (two-way mixed-effects, absolute agreement, single measurement); other abbreviations as in Part A. Values are in min/day (weighted weekly average where applicable). For reliability, Overall, Paper, and Electronic are restricted to children who completed the SSF-PAQ in the same mode on Day 7 and Day 10. Bold ICC values indicate ICC ≥ 0.50.

**eTable 4. Test–retest reliability of setting-specific physical activity time in the SSF-PAQ**

| SSF-PAQ item | Period | n | ICC [95% CI] | Day 7 Med, min/day (Q1–Q3) | Day 10 Med, min/day (Q1–Q3) | % > 0 (Day 7, Day 10) |
|---|---|---|---|---|---|---|
| **SSF-PAQ Original version, overall (paper + electronic)** | | | | | | |
| Organized PA (lessons) | Weekday | 79 | **0.88 [0.81, 0.92]** | 0 (0–11) | 0 (0–10) | 29, 28 |
| | Weekend | 79 | **0.88 [0.82, 0.92]** | 0 (0–0) | 0 (0–0) | 16, 16 |
| | Weekly | 79 | **0.86 [0.79, 0.91]** | 0 (0–9) | 0 (0–9) | 37, 35 |
| Organized MVPA | Weekday | 79 | **0.87 [0.81, 0.92]** | 0 (0–0) | 0 (0–2) | 24, 27 |
| | Weekend | 79 | **0.84 [0.76, 0.90]** | 0 (0–0) | 0 (0–0) | 14, 14 |
| | Weekly | 79 | **0.86 [0.79, 0.91]** | 0 (0–3) | 0 (0–3) | 32, 33 |
| Outdoor active play: park | Weekday | 79 | **0.78 [0.67, 0.85]** | 0 (0–25) | 0 (0–24) | 48, 42 |
| | Weekend | 79 | **0.55 [0.37, 0.68]** | 60 (30–120) | 60 (1–90) | 82, 75 |
| | Weekly | 79 | **0.66 [0.51, 0.77]** | 26 (11–44) | 24 (9–36) | 87, 82 |
| Outdoor active play: home premises | Weekday | 79 | **0.89 [0.84, 0.93]** | 0 (0–12) | 0 (0–15) | 43, 46 |
| | Weekend | 79 | 0.34 [0.13, 0.52] | 10 (0–30) | 8 (0–45) | 54, 52 |
| | Weekly | 79 | **0.76 [0.65, 0.84]** | 7 (0–22) | 7 (0–20) | 62, 62 |
| Outdoor active play: street | Weekday | 79 | **0.88 [0.82, 0.92]** | 0 (0–0) | 0 (0–0) | 24, 22 |
| | Weekend | 79 | **0.75 [0.64, 0.83]** | 0 (0–0) | 0 (0–0) | 18, 18 |
| | Weekly | 79 | **0.85 [0.78, 0.90]** | 0 (0–2) | 0 (0–1) | 27, 27 |
| Outdoor active play: other | Weekday | 79 | 0.12 [−0.10, 0.33] | 0 (0–0) | 0 (0–0) | 8, 8 |
| | Weekend | 79 | **0.54 [0.37, 0.68]** | 0 (0–0) | 0 (0–0) | 19, 16 |
| | Weekly | 79 | 0.27 [0.06, 0.46] | 0 (0–0) | 0 (0–0) | 22, 20 |
| Outdoor active play: MVPA (total) | Weekday | 79 | **0.90 [0.85, 0.93]** | 0 (0–12) | 1 (0–11) | 42, 51 |
| | Weekend | 79 | **0.64 [0.49, 0.76]** | 15 (0–30) | 20 (8–30) | 72, 81 |
| | Weekly | 79 | **0.78 [0.68, 0.85]** | 9 (1–18) | 9 (4–18) | 76, 85 |
| Indoor active play: active video games | Weekday | 79 | **0.91 [0.86, 0.94]** | 0 (0–0) | 0 (0–0) | 18, 14 |
| | Weekend | 79 | **0.84 [0.76, 0.90]** | 0 (0–0) | 0 (0–0) | 19, 20 |
| | Weekly | 79 | **0.89 [0.84, 0.93]** | 0 (0–0) | 0 (0–0) | 22, 23 |
| Indoor active play: exercise | Weekday | 79 | **0.83 [0.74, 0.89]** | 10 (0–30) | 8 (0–21) | 62, 61 |
| | Weekend | 79 | **0.86 [0.79, 0.91]** | 30 (0–34) | 20 (0–30) | 68, 72 |
| | Weekly | 79 | **0.88 [0.82, 0.92]** | 17 (1–30) | 11 (0–28) | 75, 73 |
| Indoor active play: facility | Weekday | 79 | **0.57 [0.40, 0.70]** | 0 (0–0) | 0 (0–0) | 14, 10 |
| | Weekend | 79 | **0.51 [0.33, 0.66]** | 0 (0–30) | 0 (0–30) | 34, 30 |
| | Weekly | 79 | **0.57 [0.40, 0.70]** | 0 (0–14) | 0 (0–9) | 35, 34 |
| Indoor active play: other | Weekday | 79 | **0.83 [0.75, 0.89]** | 0 (0–0) | 0 (0–0) | 11, 9 |
| | Weekend | 79 | **0.51 [0.32, 0.65]** | 0 (0–0) | 0 (0–0) | 16, 16 |
| | Weekly | 79 | **0.83 [0.74, 0.89]** | 0 (0–0) | 0 (0–0) | 16, 16 |
| Indoor active play: MVPA (total) | Weekday | 79 | **0.88 [0.82, 0.92]** | 0 (0–6) | 0 (0–7) | 35, 44 |
| | Weekend | 79 | **0.78 [0.68, 0.85]** | 5 (0–24) | 15 (0–30) | 56, 72 |
| | Weekly | 79 | **0.85 [0.78, 0.90]** | 3 (0–10) | 6 (0–12) | 59, 72 |
| **SSF-PAQ 24-h version, overall (paper + electronic)** | | | | | | |
| Organized PA (lessons) | Weekday | 39 | **0.90 [0.82, 0.95]** | 0 (0–10) | 0 (0–10) | 41, 41 |
| | Weekend | 39 | **0.94 [0.90, 0.97]** | 0 (0–0) | 0 (0–0) | 15, 21 |
| | Weekly | 39 | **0.89 [0.81, 0.94]** | 6 (0–9) | 6 (0–9) | 54, 56 |
| Organized MVPA | Weekday | 39 | **0.84 [0.71, 0.91]** | 0 (0–2) | 0 (0–4) | 31, 36 |
| | Weekend | 39 | **0.95 [0.90, 0.97]** | 0 (0–0) | 0 (0–0) | 15, 18 |
| | Weekly | 39 | **0.84 [0.71, 0.91]** | 0 (0–4) | 2 (0–4) | 44, 51 |
| Outdoor active play: park | Weekday | 39 | **0.87 [0.76, 0.93]** | 6 (0–30) | 10 (0–30) | 56, 54 |
| | Weekend | 39 | **0.65 [0.43, 0.80]** | 60 (30–90) | 50 (30–68) | 87, 92 |
| | Weekly | 39 | **0.82 [0.69, 0.90]** | 26 (15–42) | 26 (12–34) | 95, 92 |
| Outdoor active play: home premises | Weekday | 39 | **0.78 [0.63, 0.88]** | 0 (0–0) | 0 (0–0) | 8, 5 |
| | Weekend | 39 | 0.27 [−0.03, 0.53] | 0 (0–0) | 0 (0–2) | 18, 26 |
| | Weekly | 39 | **0.91 [0.83, 0.95]** | 0 (0–0) | 0 (0–1) | 23, 26 |
| Outdoor active play: street | Weekday | 39 | 0.12 [−0.20, 0.41] | 0 (0–0) | 0 (0–0) | 5, 5 |
| | Weekend | 39 | **0.74 [0.56, 0.85]** | 0 (0–0) | 0 (0–0) | 10, 5 |
| | Weekly | 39 | 0.19 [−0.12, 0.47] | 0 (0–0) | 0 (0–0) | 13, 10 |
| Outdoor active play: other | Weekday | 39 | — | 0 (0–0) | 0 (0–0) | 0, 0 |
| | Weekend | 39 | **0.65 [0.43, 0.80]** | 0 (0–0) | 0 (0–0) | 10, 3 |
| | Weekly | 39 | **0.65 [0.43, 0.80]** | 0 (0–0) | 0 (0–0) | 10, 3 |
| Outdoor active play: MVPA (total) | Weekday | 39 | 0.43 [0.15, 0.65] | 6 (0–15) | 8 (0–20) | 59, 72 |
| | Weekend | 39 | **0.61 [0.36, 0.77]** | 15 (1–30) | 15 (9–30) | 74, 87 |
| | Weekly | 39 | **0.63 [0.39, 0.79]** | 10 (2–17) | 14 (5–20) | 82, 92 |

eTable 4. Continued.

| SSF-PAQ item | Period | n | ICC [95% CI] | Day 7 Med, min/day (Q1–Q3) | Day 10 Med, min/day (Q1–Q3) | % > 0 (Day 7, Day 10) |
|---|---|---|---|---|---|---|
| Indoor active play: active video games | Weekday | 39 | **0.99 [0.99, 1.00]** | 0 (0–0) | 0 (0–0) | 10, 8 |
| | Weekend | 39 | **0.83 [0.70, 0.91]** | 0 (0–0) | 0 (0–0) | 13, 10 |
| | Weekly | 39 | **0.98 [0.96, 0.99]** | 0 (0–0) | 0 (0–0) | 15, 10 |
| Indoor active play: exercise | Weekday | 39 | 0.29 [−0.02, 0.55] | 0 (0–11) | 0 (0–11) | 33, 28 |
| | Weekend | 39 | **0.53 [0.26, 0.72]** | 10 (0–30) | 0 (0–22) | 51, 41 |
| | Weekly | 39 | **0.58 [0.32, 0.75]** | 3 (0–16) | 0 (0–15) | 59, 44 |
| Indoor active play: facility | Weekday | 39 | — | 0 (0–0) | 0 (0–0) | 0, 0 |
| | Weekend | 39 | **0.75 [0.57, 0.86]** | 0 (0–18) | 0 (0–2) | 28, 26 |
| | Weekly | 39 | **0.75 [0.57, 0.86]** | 0 (0–5) | 0 (0–1) | 28, 26 |
| Indoor active play: other | Weekday | 39 | — | 0 (0–0) | 0 (0–0) | 0, 0 |
| | Weekend | 39 | 0.30 [−0.01, 0.56] | 0 (0–0) | 0 (0–0) | 10, 13 |
| | Weekly | 39 | 0.30 [−0.01, 0.56] | 0 (0–0) | 0 (0–0) | 10, 13 |
| Indoor active play: MVPA (total) | Weekday | 39 | **0.76 [0.59, 0.87]** | 0 (0–10) | 0 (0–12) | 41, 49 |
| | Weekend | 39 | **0.73 [0.55, 0.85]** | 5 (0–15) | 0 (0–12) | 54, 41 |
| | Weekly | 39 | **0.81 [0.66, 0.89]** | 4 (0–11) | 4 (0–11) | 59, 59 |
| At preschool: outdoor | Weekday | 39 | **0.88 [0.78, 0.94]** | 16 (0–60) | 18 (0–60) | 56, 54 |
| | Weekend | 39 | — | 0 (0–0) | 0 (0–0) | 0, 8 |
| | Weekly | 39 | **0.90 [0.82, 0.95]** | 11 (0–43) | 17 (0–43) | 56, 54 |
| At preschool: indoor | Weekday | 39 | 0.48 [0.20, 0.69] | 15 (0–45) | 30 (0–39) | 67, 72 |
| | Weekend | 39 | 0.10 [−0.22, 0.40] | 0 (0–0) | 0 (0–0) | 3, 5 |
| | Weekly | 39 | **0.52 [0.25, 0.72]** | 11 (0–32) | 21 (0–28) | 67, 72 |

Values are the average time per day within the period (min/day): the weekly value is the total weekly time divided by 7, and the weekday and weekend values are the means over weekdays and over weekend/holidays. Activities done on only some days (e.g., organized lessons) therefore show low daily medians even when they last longer on the days they occur (for example, a 60-min weekly lesson equals about 9 min/day). The last column gives the percentage of children reporting more than 0 min/day; a median of 0 therefore means that fewer than half of the children reported the activity at all. For organized lessons this follows the questionnaire's filter question on whether the child takes part in any organized sports or exercise lesson, which 37% of the Original-version sample answered affirmatively at Day 7; the remaining children report 0 min on every day. Within indoor active play, 'active video games' is active play while using a screen (e.g., Wii Sports, Nintendo Switch games, follow-along videos), 'exercise' is active play indoors without games or videos (e.g., running around, active play with a parent), 'facility' is an indoor exercise facility (e.g., gymnasium, swimming pool, children's center), and 'other' is any other indoor setting. CI, confidence interval; ICC, intraclass correlation coefficient (two-way mixed-effects, absolute agreement, single measurement); Med, median; MVPA, moderate-to-vigorous physical activity; PA, physical activity; Q1–Q3, interquartile range; SSF-PAQ, Sasakawa Sports Foundation Preschoolers' Activity Questionnaire. Overall pools paper and electronic modes among children completing the same version and mode on Day 7 and Day 10 (Original, n = 79; 24-h, n = 39). The Original version asks about activity outside of preschool; the 24-h version additionally asks about activity at preschool. MVPA rows are the parent-reported time spent in activity intense enough to make the child breathe harder within each domain (organized lessons, outdoor play, and indoor play); the questionnaire collects this higher-intensity portion only at the domain level, not per location or setting.

Bold ICC values indicate ICC ≥ 0.50. ICC is shown as — where all children reported the same value (all zeros) on Day 7 or Day 10, leaving the ICC undefined; medians and quartiles are still shown. Settings reported by a minority of children (e.g., organized lessons, street play and 'other') have many zero values, which can inflate or destabilize the ICC; these should be interpreted together with the percentage reporting > 0 rather than from the ICC alone. Weekend rows for the at-preschool items are retained for completeness: almost no child attended preschool on weekends (0–3 children per administration reported nonzero time), so their weekend ICCs are uninformative.

#### eTable 5. Measurement error of setting-specific physical activity time in the SSF-PAQ

| SSF-PAQ item | Period | n | Day 7 Mean (SD) | Day 10 Mean (SD) | Difference Mean (SD) | CV (%) | SEM | SDCind |
|---|---|---|---|---|---|---|---|---|
| **SSF-PAQ Original version, overall (paper + electronic)** | | | | | | | | |
| Organized PA (lessons) | Weekday | 79 | 5 (11) | 4 (8) | 1 (5) | 9.1 | 3.4 | 9.4 |
| | Weekend | 79 | 8 (22) | 7 (16) | 1 (9) | 10.3 | 6.5 | 18.1 |
| | Weekly | 79 | 6 (13) | 5 (9) | 1 (6) | 8.5 | 4.1 | 11.4 |
| Organized MVPA | Weekday | 79 | 2 (4) | 2 (3) | 0 (2) | 23.9 | 1.3 | 3.5 |
| | Weekend | 79 | 4 (13) | 3 (9) | 1 (6) | 14.4 | 4.6 | 12.6 |
| | Weekly | 79 | 2 (5) | 2 (4) | 0 (2) | 16.3 | 1.7 | 4.8 |
| Outdoor active play: park | Weekday | 79 | 15 (22) | 15 (24) | 1 (16) | 41.9 | 10.9 | 30.3 |
| | Weekend | 79 | 68 (57) | 61 (67) | 7 (59) | 40.1 | 42.0 | 116.3 |
| | Weekly | 79 | 30 (25) | 28 (31) | 2 (23) | 39.0 | 16.4 | 45.5 |
| Outdoor active play: home premises | Weekday | 79 | 11 (20) | 10 (18) | 0 (9) | 56.8 | 6.4 | 17.6 |
| | Weekend | 79 | 21 (29) | 25 (41) | −4 (41) | 64.3 | 28.8 | 79.9 |
| | Weekly | 79 | 14 (19) | 14 (20) | −1 (14) | 54.4 | 9.6 | 26.7 |
| Outdoor active play: street | Weekday | 79 | 4 (9) | 3 (8) | 0 (4) | 69.8 | 2.9 | 8.0 |
| | Weekend | 79 | 7 (18) | 6 (16) | 1 (12) | 62.7 | 8.3 | 23.0 |
| | Weekly | 79 | 5 (11) | 4 (9) | 0 (5) | 63.2 | 3.8 | 10.4 |
| Outdoor active play: other | Weekday | 79 | 3 (16) | 1 (4) | 2 (15) | 99.0 | 11.0 | 30.4 |
| | Weekend | 79 | 15 (37) | 10 (28) | 5 (31) | 94.8 | 22.3 | 61.8 |
| | Weekly | 79 | 6 (18) | 3 (9) | 3 (17) | 86.8 | 12.0 | 33.2 |
| Outdoor active play: MVPA (total) | Weekday | 79 | 8 (14) | 9 (16) | −1 (7) | 51.7 | 4.7 | 13.1 |
| | Weekend | 79 | 25 (32) | 34 (63) | −9 (42) | 49.0 | 29.8 | 82.7 |
| | Weekly | 79 | 13 (17) | 16 (26) | −3 (14) | 50.2 | 10.3 | 28.5 |
| Indoor active play: active video games | Weekday | 79 | 4 (13) | 4 (15) | 0 (6) | 53.0 | 4.3 | 11.8 |
| | Weekend | 79 | 8 (22) | 7 (20) | 1 (12) | 73.7 | 8.2 | 22.8 |
| | Weekly | 79 | 5 (15) | 5 (16) | 0 (7) | 69.1 | 5.1 | 14.1 |
| Indoor active play: exercise | Weekday | 79 | 17 (22) | 16 (25) | 1 (14) | 49.1 | 9.7 | 26.9 |
| | Weekend | 79 | 32 (42) | 30 (43) | 1 (23) | 52.9 | 15.8 | 43.9 |
| | Weekly | 79 | 21 (26) | 20 (29) | 1 (13) | 47.7 | 9.5 | 26.2 |
| Indoor active play: facility | Weekday | 79 | 2 (5) | 1 (5) | 0 (5) | 86.3 | 3.4 | 9.4 |
| | Weekend | 79 | 20 (33) | 18 (33) | 2 (33) | 80.9 | 23.2 | 64.2 |
| | Weekly | 79 | 7 (11) | 6 (10) | 1 (10) | 79.7 | 7.1 | 19.6 |
| Indoor active play: other | Weekday | 79 | 3 (9) | 3 (9) | 0 (5) | 59.4 | 3.6 | 10.1 |
| | Weekend | 79 | 6 (16) | 7 (18) | −1 (17) | 90.9 | 11.9 | 32.9 |
| | Weekly | 79 | 4 (10) | 4 (10) | 0 (6) | 84.9 | 4.2 | 11.8 |
| Indoor active play: MVPA (total) | Weekday | 79 | 6 (12) | 6 (12) | 0 (6) | 66.3 | 4.1 | 11.3 |
| | Weekend | 79 | 18 (28) | 20 (27) | −2 (18) | 64.2 | 12.7 | 35.3 |
| | Weekly | 79 | 9 (15) | 10 (15) | 0 (8) | 58.6 | 5.7 | 15.8 |

eTable 5. Continued.

| SSF-PAQ item | Period | n | Day 7 Mean (SD) | Day 10 Mean (SD) | Difference Mean (SD) | CV (%) | SEM | SDCind |
|---|---|---|---|---|---|---|---|---|
| **SSF-PAQ 24-h version, overall (paper + electronic)** | | | | | | | | |
| Organized PA (lessons) | Weekday | 39 | 6 (8) | 6 (9) | 0 (4) | 5.9 | 2.7 | 7.6 |
| | Weekend | 39 | 5 (13) | 6 (13) | −1 (4) | 35.4 | 3.0 | 8.5 |
| | Weekly | 39 | 5 (6) | 6 (7) | −1 (3) | 11.8 | 2.1 | 6.0 |
| Organized MVPA | Weekday | 39 | 2 (5) | 3 (7) | −1 (3) | 40.4 | 2.5 | 6.8 |
| | Weekend | 39 | 3 (8) | 4 (10) | −1 (3) | 24.2 | 2.0 | 5.7 |
| | Weekly | 39 | 3 (4) | 3 (5) | −1 (3) | 35.4 | 1.8 | 5.1 |
| Outdoor active play: park | Weekday | 39 | 17 (21) | 18 (23) | 0 (11) | 55.3 | 8.0 | 22.1 |
| | Weekend | 39 | 63 (46) | 55 (37) | 7 (35) | 35.3 | 24.9 | 69.0 |
| | Weekly | 39 | 30 (22) | 28 (22) | 2 (13) | 32.7 | 9.3 | 25.8 |
| Outdoor active play: home premises | Weekday | 39 | 2 (7) | 1 (4) | 1 (4) | 75.9 | 2.8 | 7.8 |
| | Weekend | 39 | 2 (6) | 6 (15) | −4 (13) | 62.6 | 9.8 | 27.1 |
| | Weekly | 39 | 2 (6) | 2 (6) | −1 (3) | 58.3 | 1.9 | 5.3 |
| Outdoor active play: street | Weekday | 39 | 2 (9) | 0 (2) | 1 (9) | 94.3 | 6.2 | 17.3 |
| | Weekend | 39 | 3 (11) | 2 (7) | 2 (7) | 82.5 | 4.8 | 13.3 |
| | Weekly | 39 | 2 (8) | 1 (3) | 1 (8) | 78.6 | 5.5 | 15.3 |
| Outdoor active play: other | Weekday | 39 | 0 (0) | 0 (0) | 0 (0) | — | 0.0 | 0.0 |
| | Weekend | 39 | 5 (18) | 2 (10) | 3 (12) | 113.1 | 8.5 | 23.5 |
| | Weekly | 39 | 1 (5) | 0 (3) | 1 (3) | 113.1 | 2.4 | 6.7 |
| Outdoor active play: MVPA (total) | Weekday | 39 | 8 (10) | 13 (14) | −5 (13) | 41.1 | 9.5 | 26.5 |
| | Weekend | 39 | 22 (28) | 22 (20) | 0 (21) | 46.4 | 15.0 | 41.6 |
| | Weekly | 39 | 12 (12) | 16 (13) | −3 (11) | 41.5 | 7.8 | 21.6 |
| Indoor active play: active video games | Weekday | 39 | 2 (10) | 2 (10) | 0 (1) | 47.1 | 0.8 | 2.3 |
| | Weekend | 39 | 3 (11) | 2 (10) | 1 (6) | 101.0 | 4.3 | 12.1 |
| | Weekly | 39 | 3 (10) | 2 (10) | 0 (2) | 83.1 | 1.4 | 4.0 |
| Indoor active play: exercise | Weekday | 39 | 10 (18) | 7 (13) | 3 (19) | 84.0 | 13.4 | 37.2 |
| | Weekend | 39 | 22 (38) | 14 (23) | 8 (30) | 81.4 | 21.6 | 60.0 |
| | Weekly | 39 | 13 (20) | 9 (14) | 5 (16) | 73.6 | 11.5 | 32.0 |
| Indoor active play: facility | Weekday | 39 | 0 (0) | 0 (0) | 0 (0) | — | 0.0 | 0.0 |
| | Weekend | 39 | 12 (25) | 10 (24) | 2 (18) | 83.6 | 12.3 | 34.1 |
| | Weekly | 39 | 3 (7) | 3 (7) | 0 (5) | 83.6 | 3.5 | 9.8 |
| Indoor active play: other | Weekday | 39 | 0 (0) | 0 (0) | 0 (0) | — | 0.0 | 0.0 |
| | Weekend | 39 | 3 (11) | 7 (22) | −4 (21) | 111.1 | 14.8 | 41.0 |
| | Weekly | 39 | 1 (3) | 2 (6) | −1 (6) | 111.1 | 4.2 | 11.7 |
| Indoor active play: MVPA (total) | Weekday | 39 | 9 (23) | 10 (15) | 0 (13) | 71.0 | 9.3 | 25.9 |
| | Weekend | 39 | 14 (24) | 10 (19) | 4 (16) | 81.1 | 11.3 | 31.4 |
| | Weekly | 39 | 11 (22) | 10 (16) | 1 (12) | 60.1 | 8.3 | 23.0 |
| At preschool: outdoor | Weekday | 39 | 32 (37) | 31 (35) | 1 (18) | 40.0 | 12.4 | 34.5 |
| | Weekend | 39 | 0 (0) | 3 (11) | −3 (11) | 141.4 | 7.8 | 21.6 |
| | Weekly | 39 | 23 (26) | 23 (26) | 0 (12) | 38.3 | 8.1 | 22.6 |

eTable 5. Continued.

| SSF-PAQ item | Period | n | Day 7 Mean (SD) | Day 10 Mean (SD) | Difference Mean (SD) | CV (%) | SEM | SDCind |
|---|---|---|---|---|---|---|---|---|
| At preschool: indoor | Weekday | 39 | 31 (40) | 28 (28) | 3 (35) | 51.7 | 24.6 | 68.2 |
| | Weekend | 39 | 0 (1) | 1 (5) | −1 (5) | 70.7 | 3.4 | 9.4 |
| | Weekly | 39 | 22 (28) | 20 (20) | 2 (24) | 51.2 | 16.8 | 46.6 |

Values are the average time per day within the period (min/day; the weekly value is the total weekly time divided by 7), so activities done on only some days (e.g., organized lessons) show low daily means. Within indoor active play, ‘active video games’ is active play while using a screen (e.g., Wii Sports, Nintendo Switch games, follow-along videos), ‘exercise’ is active play indoors without games or videos (e.g., running around, active play with a parent), ‘facility’ is an indoor exercise facility (e.g., gymnasium, swimming pool, children’s center), and ‘other’ is any other indoor setting. Difference is Day 7 minus Day 10. CV, within-child coefficient of variation (mean of 100 × |Day 7 − Day 10|/√2 ÷ mean); SEM, standard error of measurement from a mixed model with random effects for participant and measurement occasion; SDCind = 1.96 × √2 × SEM is the individual-level smallest detectable change (also termed the minimal detectable change at the 95% level, MDC95). Overall pools paper and electronic modes (Original, n = 79; 24-h, n = 39). MVPA rows are the parent-reported ‘breathing harder’ time within each domain (organized lessons, outdoor play, indoor play), collected only at the domain level, not per location or setting. MVPA, moderate-to-vigorous physical activity; PA, physical activity; SD, standard deviation; SDC, smallest detectable change; SDCind, individual-level smallest detectable change; SEM, standard error of measurement; SSF-PAQ, Sasakawa Sports Foundation Preschoolers' Activity Questionnaire.

For settings reported by a minority of children (e.g., organized lessons, street play and ‘other’), the mean approaches zero and the CV is unstable or undefined; such values should be interpreted with caution, alongside the percentage of children reporting > 0 min/day given in eTable 4. CV is shown as — where every child reported 0 min on both days, leaving the CV undefined.

#### eTable 6. Concurrent validity of the main behaviours by accelerometer wear-time criterion

| SSF-PAQ item | n (360) | ρ [95% CI] (360) | n (600) | ρ [95% CI] (600) | n (960) | ρ [95% CI] (960) |
|---|---|---|---|---|---|---|
| **SSF-PAQ Original version, overall (paper + electronic)** | | | | | | |
| Organized PA and AP | 130 | **0.20 [0.03, 0.36]** | 127 | **0.24 [0.07, 0.39]** | 107 | **0.25 [0.06, 0.42]** |
| MVPA | 130 | **0.27 [0.10, 0.42]** | 127 | **0.29 [0.13, 0.45]** | 107 | **0.34 [0.16, 0.49]** |
| Total screen time | 129 | **0.84 [0.77, 0.88]** | 126 | **0.83 [0.77, 0.88]** | 106 | **0.83 [0.75, 0.88]** |
| Night sleep | 130 | **0.79 [0.71, 0.84]** | 127 | **0.78 [0.71, 0.84]** | 107 | **0.82 [0.75, 0.87]** |
| **SSF-PAQ Original version, paper** | | | | | | |
| Organized PA and AP | 74 | 0.15 [−0.08, 0.37] | 72 | 0.17 [−0.06, 0.39] | 61 | 0.18 [−0.07, 0.42] |
| MVPA | 74 | 0.14 [−0.09, 0.36] | 72 | 0.15 [−0.08, 0.37] | 61 | 0.21 [−0.05, 0.43] |
| Total screen time | 73 | **0.82 [0.72, 0.88]** | 71 | **0.82 [0.72, 0.88]** | 60 | **0.81 [0.70, 0.88]** |
| Night sleep | 74 | **0.73 [0.60, 0.82]** | 72 | **0.74 [0.61, 0.83]** | 61 | **0.77 [0.64, 0.85]** |
| **SSF-PAQ Original version, electronic** | | | | | | |
| Organized PA and AP | 56 | **0.27 [0.01, 0.50]** | 55 | **0.32 [0.06, 0.54]** | 46 | 0.28 [−0.01, 0.53] |
| MVPA | 56 | **0.42 [0.17, 0.61]** | 55 | **0.47 [0.24, 0.66]** | 46 | **0.48 [0.23, 0.68]** |
| Total screen time | 56 | **0.85 [0.76, 0.91]** | 55 | **0.84 [0.74, 0.91]** | 46 | **0.84 [0.73, 0.91]** |
| Night sleep | 56 | **0.86 [0.77, 0.92]** | 55 | **0.85 [0.76, 0.91]** | 46 | **0.90 [0.82, 0.94]** |
| **SSF-PAQ 24-h version, overall (paper + electronic)** | | | | | | |
| Organized PA and AP | 40 | 0.13 [−0.19, 0.42] | 40 | 0.17 [−0.15, 0.46] | 35 | 0.22 [−0.12, 0.52] |
| MVPA | 40 | 0.07 [−0.24, 0.38] | 40 | 0.06 [−0.26, 0.36] | 35 | 0.00 [−0.33, 0.34] |
| Total screen time | 40 | **0.68 [0.47, 0.82]** | 40 | **0.68 [0.47, 0.82]** | 35 | **0.66 [0.42, 0.82]** |
| Night sleep | 40 | **0.63 [0.40, 0.79]** | 40 | **0.63 [0.40, 0.79]** | 35 | **0.63 [0.38, 0.80]** |
| **SSF-PAQ 24-h version, paper** | | | | | | |
| Organized PA and AP | 21 | 0.30 [−0.15, 0.65] | 21 | 0.36 [−0.08, 0.69] | 20 | **0.46 [0.02, 0.75]** |
| MVPA | 21 | 0.17 [−0.29, 0.56] | 21 | 0.16 [−0.29, 0.56] | 20 | 0.16 [−0.31, 0.56] |
| Total screen time | 21 | **0.77 [0.51, 0.90]** | 21 | **0.77 [0.51, 0.90]** | 20 | **0.77 [0.50, 0.90]** |
| Night sleep | 21 | **0.67 [0.34, 0.86]** | 21 | **0.70 [0.39, 0.87]** | 20 | **0.73 [0.42, 0.88]** |
| **SSF-PAQ 24-h version, electronic** | | | | | | |
| Organized PA and AP | 19 | 0.01 [−0.45, 0.46] | 19 | 0.01 [−0.45, 0.46] | 15 | −0.06 [−0.56, 0.46] |
| MVPA | 19 | 0.04 [−0.42, 0.49] | 19 | 0.06 [−0.41, 0.50] | 15 | −0.06 [−0.56, 0.46] |
| Total screen time | 19 | **0.60 [0.20, 0.83]** | 19 | **0.60 [0.20, 0.83]** | 15 | **0.57 [0.08, 0.84]** |
| Night sleep | 19 | **0.71 [0.38, 0.88]** | 19 | **0.71 [0.38, 0.88]** | 15 | **0.77 [0.42, 0.92]** |

Values are weighted weekly averages (min/day); the main analysis used the 600 min/day wear-time criterion. The 360 and 600 min/day criteria refer to wear time within the 07:00–21:00 waking window; the 960 min/day criterion refers to all-day wear time including sleep. The criterion determines which days are valid for each child, so criterion values—and therefore ρ—can change even when n is unchanged. Overall combines paper and electronic modes; the paper and electronic sections comprise the children who completed the Day 7 questionnaire in that mode. Bold ρ values indicate 95% CIs that do not include zero.

Abbreviations: AP, active play; CI, confidence interval; MVPA, moderate-to-vigorous physical activity; organized PA, organized physical activity; ρ, Spearman rank correlation coefficient; SSF-PAQ, Sasakawa Sports Foundation Preschoolers' Activity Questionnaire.

#### eTable 7. Concurrent validity and test–retest reliability among children aged 3–5 years

##### A. Concurrent validity (weighted weekly average)

| SSF-PAQ item | n | ρ [95% CI] | Criterion Med (Q1–Q3) | SSF-PAQ Med (Q1–Q3) | P |
|---|---|---|---|---|---|
| **SSF-PAQ Original version, overall (paper + electronic)** | | | | | |
| Organized PA and AP | 122 | **0.24 [0.06, 0.40]** | 238 (200–281) | 73 (45–101) | NA |
| MVPA | 122 | **0.30 [0.13, 0.46]** | 45 (32–60) | 13 (4–26) | NA |
| Total screen time | 121 | **0.84 [0.78, 0.89]** | 86 (43–133) | 90 (58–150) | < 0.001 |
| Night sleep | 122 | **0.80 [0.72, 0.85]** | 590 (570–612) | 602 (579–627) | < 0.001 |
| **SSF-PAQ Original version, paper** | | | | | |
| Organized PA and AP | 69 | 0.16 [−0.08, 0.38] | 235 (208–278) | 63 (41–99) | NA |
| MVPA | 69 | 0.16 [−0.08, 0.39] | 46 (33–60) | 11 (4–26) | NA |
| Total screen time | 68 | **0.82 [0.72, 0.88]** | 88 (43–135) | 107 (58–150) | 0.002 |
| Night sleep | 69 | **0.75 [0.63, 0.84]** | 587 (570–619) | 600 (585–624) | < 0.001 |
| **SSF-PAQ Original version, electronic** | | | | | |
| Organized PA and AP | 53 | **0.33 [0.07, 0.55]** | 239 (191–288) | 81 (54–120) | NA |
| MVPA | 53 | **0.48 [0.24, 0.67]** | 45 (32–57) | 14 (4–24) | NA |
| Total screen time | 53 | **0.86 [0.76, 0.92]** | 78 (39–126) | 90 (45–133) | 0.183 |
| Night sleep | 53 | **0.86 [0.77, 0.92]** | 596 (575–609) | 604 (573–628) | 0.001 |
| **SSF-PAQ 24-h version, overall (paper + electronic)** | | | | | |
| Organized PA and AP | 27 | 0.35 [−0.03, 0.65] | 393 (342–429) | 93 (47–162) | NA |
| MVPA | 27 | 0.23 [−0.16, 0.56] | 72 (55–86) | 21 (10–40) | NA |
| Total screen time | 27 | **0.55 [0.21, 0.77]** | 132 (73–201) | 107 (66–167) | 0.313 |
| Night sleep | 27 | **0.53 [0.18, 0.76]** | 602 (584–615) | 600 (571–610) | 0.148 |
| **SSF-PAQ 24-h version, paper** | | | | | |
| Organized PA and AP | 14 | 0.35 [−0.22, 0.74] | 397 (355–429) | 99 (42–164) | NA |
| MVPA | 14 | 0.04 [−0.50, 0.56] | 82 (56–86) | 19 (4–41) | NA |
| Total screen time | 14 | **0.74 [0.35, 0.91]** | 129 (81–170) | 107 (79–186) | 0.761 |
| Night sleep | 14 | **0.82 [0.52, 0.94]** | 605 (576–616) | 604 (574–609) | 0.670 |
| **SSF-PAQ 24-h version, electronic** | | | | | |
| Organized PA and AP | 13 | 0.50 [−0.07, 0.82] | 346 (341–420) | 83 (64–144) | NA |
| MVPA | 13 | 0.53 [−0.03, 0.84] | 70 (55–85) | 21 (14–35) | NA |
| Total screen time | 13 | 0.37 [−0.22, 0.77] | 132 (68–212) | 107 (58–150) | 0.273 |
| Night sleep | 13 | 0.39 [−0.20, 0.77] | 602 (596–611) | 591 (570–617) | 0.146 |

##### B. Test–retest reliability (weighted weekly average)

| SSF-PAQ item | n | ICC [95% CI] | Day 7 Med (Q1–Q3) | Day 10 Med (Q1–Q3) |
|---|---|---|---|---|
| **SSF-PAQ Original version, overall (paper + electronic)** | | | | |
| Organized PA and AP | 77 | **0.84 [0.76, 0.90]** | 86 (54–116) | 74 (50–106) |
| MVPA | 77 | **0.85 [0.77, 0.90]** | 17 (4–33) | 17 (11–34) |
| Total screen time | 75 | **0.79 [0.68, 0.86]** | 107 (51–150) | 107 (45–150) |
| Night sleep | 77 | **0.96 [0.94, 0.98]** | 604 (579–621) | 600 (579–621) |
| **SSF-PAQ Original version, paper** | | | | |
| Organized PA and AP | 37 | **0.81 [0.62, 0.90]** | 90 (54–100) | 76 (39–102) |
| MVPA | 37 | **0.86 [0.74, 0.92]** | 17 (6–34) | 19 (11–34) |
| Total screen time | 35 | **0.87 [0.76, 0.93]** | 107 (90–159) | 107 (90–159) |
| Night sleep | 37 | **0.98 [0.96, 0.99]** | 604 (579–621) | 600 (585–621) |
| **SSF-PAQ Original version, electronic** | | | | |
| Organized PA and AP | 40 | **0.86 [0.75, 0.92]** | 85 (53–140) | 73 (54–141) |
| MVPA | 40 | **0.84 [0.72, 0.91]** | 17 (4–26) | 17 (12–34) |
| Total screen time | 40 | **0.70 [0.50, 0.83]** | 58 (36–114) | 58 (24–107) |
| Night sleep | 40 | **0.94 [0.90, 0.97]** | 602 (579–624) | 600 (579–630) |
| **SSF-PAQ 24-h version, overall (paper + electronic)** | | | | |
| Organized PA and AP | 26 | **0.86 [0.72, 0.94]** | 88 (46–153) | 86 (47–125) |
| MVPA | 26 | **0.90 [0.80, 0.96]** | 21 (9–42) | 23 (10–31) |
| Total screen time | 26 | **0.75 [0.52, 0.88]** | 107 (62–163) | 107 (58–146) |
| Night sleep | 26 | **0.92 [0.84, 0.97]** | 602 (571–610) | 600 (577–610) |

eTable 7. Continued.

| SSF-PAQ item | n | ICC [95% CI] | Day 7 Med (Q1–Q3) | Day 10 Med (Q1–Q3) |
|---|---|---|---|---|
| **SSF-PAQ 24-h version, paper** | | | | |
| Organized PA and AP | 13 | **0.88 [0.67, 0.96]** | 93 (41–156) | 93 (44–96) |
| MVPA | 13 | **0.85 [0.58, 0.95]** | 17 (3–45) | 20 (10–27) |
| Total screen time | 13 | **0.87 [0.65, 0.96]** | 107 (75–167) | 90 (58–150) |
| Night sleep | 13 | **0.88 [0.66, 0.96]** | 607 (572–609) | 600 (587–609) |
| **SSF-PAQ 24-h version, electronic** | | | | |
| Organized PA and AP | 13 | **0.83 [0.55, 0.95]** | 83 (64–144) | 84 (70–146) |
| MVPA | 13 | **0.94 [0.81, 0.98]** | 21 (14–35) | 29 (11–31) |
| Total screen time | 13 | **0.63 [0.14, 0.87]** | 107 (58–150) | 107 (90–133) |
| Night sleep | 13 | **0.96 [0.88, 0.99]** | 591 (570–617) | 600 (573–616) |

Restricted to children aged 3–5 years (< 72 months at measurement; 174 of the 208 children whose parents consented). The n for each analysis is shown in the table. Values are weighted weekly averages (min/day). Overall combines paper and electronic modes; the paper and electronic sections comprise the children who completed the Day 7 questionnaire in that mode. Bold ρ values indicate 95% CIs that do not include zero; bold ICC values indicate ICC ≥ 0.50. P, Wilcoxon signed-rank test; NA, not examined for the physical-activity items, as in Table 2. Abbreviations and formatting as in eTable 3 and Tables 2–3.

#### eTable 8. Concurrent validity of the main behaviours under usual-day and diary-quality sensitivity analyses

| SSF-PAQ item | Main analysis | (1) Usual days | (2) Accurate diary days | (3) Usual & accurate days |
|---|---|---|---|---|
| **SSF-PAQ Original version, overall (paper + electronic)** | | | | |
| Organized PA and AP | **0.24 [0.07, 0.39] (n = 127)** | **0.28 [0.10, 0.44] (n = 117)** | **0.25 [0.08, 0.41] (n = 123)** | **0.28 [0.10, 0.44] (n = 114)** |
| MVPA | **0.29 [0.13, 0.45] (n = 127)** | **0.30 [0.13, 0.46] (n = 117)** | **0.30 [0.13, 0.45] (n = 123)** | **0.30 [0.12, 0.46] (n = 114)** |
| Total screen time | **0.83 [0.77, 0.88] (n = 126)** | **0.85 [0.79, 0.89] (n = 118)** | **0.83 [0.77, 0.88] (n = 123)** | **0.86 [0.80, 0.90] (n = 115)** |
| Night sleep | **0.78 [0.71, 0.84] (n = 127)** | **0.78 [0.70, 0.84] (n = 121)** | **0.77 [0.69, 0.84] (n = 124)** | **0.77 [0.68, 0.83] (n = 118)** |
| **SSF-PAQ 24-h version, overall (paper + electronic)** | | | | |
| Organized PA and AP | 0.17 [−0.15, 0.46] (n = 40) | 0.17 [−0.21, 0.50] (n = 30) | −0.02 [−0.35, 0.31] (n = 36) | 0.02 [−0.38, 0.41] (n = 25) |
| MVPA | 0.06 [−0.26, 0.36] (n = 40) | 0.12 [−0.26, 0.46] (n = 30) | 0.06 [−0.27, 0.38] (n = 36) | 0.24 [−0.17, 0.58] (n = 25) |
| Total screen time | **0.68 [0.47, 0.82] (n = 40)** | **0.65 [0.38, 0.82] (n = 31)** | **0.58 [0.31, 0.76] (n = 36)** | **0.60 [0.26, 0.80] (n = 25)** |
| Night sleep | **0.63 [0.40, 0.79] (n = 40)** | **0.68 [0.44, 0.83] (n = 34)** | **0.70 [0.50, 0.84] (n = 38)** | **0.70 [0.45, 0.85] (n = 29)** |

Values are Spearman ρ [95% CI] between weekly SSF-PAQ estimates and the accelerometer/diary criteria; Overall combines paper and electronic modes. The Main analysis column corresponds to Table 2. Each sensitivity analysis recomputed the weighted weekly criterion means using only the specified day subset, leaving all other aspects unchanged: (1) usual days, days the parent reported the child had spent in the usual way; (2) accurate diary days, days rated as accurately recorded (the two most accurate of four categories); (3) usual & accurate days, days meeting both criteria. Because the criterion means are recomputed from a restricted day set, ρ can differ from the main analysis even when n is unchanged; n can be smaller because the weekly mean requires at least one weekday and one weekend/holiday within the subset. Bold ρ values indicate 95% CIs that do not include zero.

Abbreviations: AP, active play; CI, confidence interval; MVPA, moderate-to-vigorous physical activity; organized PA, organized physical activity; ρ, Spearman rank correlation coefficient; SSF-PAQ, Sasakawa Sports Foundation Preschoolers' Activity Questionnaire.

#### eTable 9. Sensitivity of night sleep validity to the definition of weekend (free-day) nights

| Period | n | ρ [95% CI],<br>next-day-free definition (main analysis) | ρ [95% CI],<br>calendar-weekend definition |
|---|---|---|---|
| Weekday | 167 | **0.68 [0.58, 0.75]** | **0.72 [0.64, 0.79]** |
| Weekend | 167 | **0.49 [0.36, 0.59]** | **0.33 [0.18, 0.45]** |
| Weekly average | 167 | **0.74 [0.67, 0.80]** | **0.75 [0.67, 0.81]** |

Each criterion night-sleep duration (diary integrated with accelerometer) was classified as weekday or weekend in two ways: ‘next-day-free’, by whether the following day is a day off (the night before a free day; the definition used in the main analysis, Table 2), which corresponds to the nights on which children can sleep in, and ‘calendar-weekend’, by the calendar day on which the night begins (Saturday and Sunday nights). The SSF-PAQ estimates and the analytic sample are identical in the two columns; only the night-classification rule differs. Because the bedtime and wake-up time items are identical in the two SSF-PAQ versions, the two versions are pooled (n = 167). Bold ρ values indicate 95% CIs that do not include zero.

Abbreviations: CI, confidence interval; ρ, Spearman rank correlation coefficient; SSF-PAQ, Sasakawa Sports Foundation Preschoolers' Activity Questionnaire.

#### eTable 10. Validity, systematic differences, test–retest reliability, and measurement error of parent-reported sleep timing in the SSF-PAQ

| Sleep-timing measure | n | ρ [95% CI] | SSF-PAQ median (Q1–Q3) | Systematic difference (min); P | ICC [95% CI] | SEM (min) | SDCind (min) | SDCgroup (SDCind/√N) |
|---|---|---|---|---|---|---|---|---|
| Bedtime (Weekday) | 167 | **0.78 [0.71, 0.83]** | 21:00 (20:00–21:30) | −16; < 0.001 | **0.99 [0.98, 0.99]** | 5.9 | 16.4 | 16.4/√N |
| Bedtime (Weekend) | 167 | **0.81 [0.76, 0.86]** | 21:00 (20:30–21:45) | −9; < 0.001 | **0.95 [0.93, 0.97]** | 12.0 | 33.2 | 33.2/√N |
| Wake-up time (Weekday) | 167 | **0.91 [0.87, 0.93]** | 06:45 (06:15–07:00) | −4; 0.002 | **0.99 [0.98, 0.99]** | 4.4 | 12.2 | 12.2/√N |
| Wake-up time (Weekend) | 167 | **0.84 [0.79, 0.88]** | 07:00 (06:30–08:00) | −2; 0.400 | **0.92 [0.89, 0.95]** | 15.3 | 42.4 | 42.4/√N |
| Midsleep (Weekday) | 167 | **0.81 [0.75, 0.86]** | 01:45 (01:15–02:15) | −10; < 0.001 | **0.99 [0.99, 1.00]** | 3.4 | 9.5 | 9.5/√N |
| Midsleep (Weekend) | 167 | **0.86 [0.81, 0.90]** | 02:00 (01:30–02:45) | −4; 0.003 | **0.96 [0.95, 0.97]** | 10.1 | 28.0 | 28.0/√N |
| Social jetlag (weekend − weekday midsleep) | 167 | **0.36 [0.22, 0.48]** | 15 (0–38) | 7; 0.003 | **0.83 [0.77, 0.88]** | 10.5 | 29.1 | 29.1/√N |

Both SSF-PAQ versions are pooled because the bedtime and wake-up time items are identical in the two versions: validity compares the Day 7 questionnaire with accelerometer-estimated sleep timing (Tracy algorithm; n = 167), and the ICC and measurement error compare Day 7 and Day 10 among children who answered in the same mode on both days (n = 118). Weekday and weekend nights were classified by whether the following day was a day off, consistent with the main sleep analysis. Clock times are shown as hh:mm; midsleep is the midpoint between sleep onset and offset, and social jetlag is the weekend-minus-weekday difference in midsleep (positive = later midsleep on weekends). The systematic difference is the median within-child difference (SSF-PAQ − criterion, min; negative = earlier reported time or smaller reported social jetlag), with P from a Wilcoxon signed-rank test. SDCind = 1.96 × √2 × SEM; SDCgroup = SDCind/√N is the smallest change detectable in the mean of N participants. Because clock times have an arbitrary origin, the coefficient of variation is not reported. ρ is shown in bold where the 95% CI excludes 0, and ICC where ≥ 0.50.

Abbreviations: CI, confidence interval; ICC, intraclass correlation coefficient; Q1–Q3, interquartile range; SDCind, individual-level smallest detectable change; SEM, standard error of measurement; ρ, Spearman rank correlation coefficient; SSF-PAQ, Sasakawa Sports Foundation Preschoolers' Activity Questionnaire.

#### eTable 11. Concurrent validity and test–retest reliability of day-based indicators of exercise and active play in the SSF-PAQ

| Indicator (days per week) | Day 7 median (Q1–Q3) | 0 days, n (%) | 7 days, n (%) | ρ [95% CI] versus total PA | ρ [95% CI] versus MVPA | ρ [95% CI] versus mean ENMO | ICC [95% CI] |
|---|---|---|---|---|---|---|---|
| Activity days (any organized lessons, outdoor active play, or indoor active play) | 7 (5–7) | 2 (1.6%) | 76 (60%) | 0.12 [−0.06, 0.29] | 0.09 [−0.09, 0.26] | 0.11 [−0.07, 0.28] | **0.65 [0.50, 0.76]** |
| Days with organized lessons | 0 (0–1) | 85 (67%) | 0 (0%) | **0.24 [0.07, 0.40]** | 0.11 [−0.07, 0.28] | **0.19 [0.02, 0.36]** | **0.99 [0.99, 0.99]** |
| Outdoor-play days | 4 (2–7) | 4 (3.1%) | 35 (28%) | **0.32 [0.16, 0.47]** | 0.16 [−0.01, 0.33] | **0.25 [0.07, 0.40]** | **0.90 [0.85, 0.93]** |
| Days with indoor active play | 6 (2–7) | 14 (11%) | 58 (46%) | −0.02 [−0.20, 0.15] | 0.06 [−0.12, 0.23] | 0.03 [−0.15, 0.20] | **0.68 [0.54, 0.78]** |

SSF-PAQ Original version. Each indicator is the number of days of the week (0–7) on which the parent reported at least 1 minute of the activity in a usual week over the past month. The first three columns of results describe the Day 7 responses of the concurrent-validity sample (n = 127). Concurrent validity compares the Day 7 questionnaire with accelerometer-derived out-of-preschool total PA, MVPA, and mean ENMO (weekly averages; n = 127); ρ is shown in bold where the 95% CI excludes 0. Test–retest reliability compares Days 7 and 10 among participants who completed the questionnaire in the same mode at both administrations (n = 79); ICC is shown in bold where ≥ 0.50. No child had seven days with organized lessons. Because an accelerometer cannot identify the days on which a child played, the accuracy of the reported number of days itself was not examined.

Abbreviations: CI, confidence interval; ENMO, Euclidean Norm Minus One; ICC, intraclass correlation coefficient; MVPA, moderate-to-vigorous physical activity; PA, physical activity; Q1–Q3, interquartile range; ρ, Spearman rank correlation coefficient; SSF-PAQ, Sasakawa Sports Foundation Preschoolers' Activity Questionnaire.
